# Prolonged SARS-CoV-2 RNA virus shedding and lymphopenia are hallmarks of COVID-19 in cancer patients with poor prognosis

**DOI:** 10.1101/2021.04.26.21250357

**Authors:** Anne-Gaëlle Goubet, Agathe Dubuisson, Arthur Geraud, François-Xavier Danlos, Safae Terrisse, Carolina Alves Costa Silva, Damien Drubay, Lea Touri, Marion Picard, Marine Mazzenga, Aymeric Silvin, Garett Dunsmore, Yacine Haddad, Eugenie Pizzato, Pierre Ly, Caroline Flament, Cléa Melenotte, Eric Solary, Michaela Fontenay, Gabriel Garcia, Corinne Balleyguier, Nathalie Lassau, Markus Maeurer, Claudia Grajeda-Iglesias, Nitharsshini Nirmalathasan, Fanny Aprahamian, Sylvère Durand, Oliver Kepp, Gladys Ferrere, Cassandra Thelemaque, Imran Lahmar, Jean-Eudes Fahrner, Lydia Meziani, Abdelhakim Ahmed-Belkacem, Nadia Saïdani, Bernard La Scola, Didier Raoult, Stéphanie Gentile, Sébastien Cortaredona, Giuseppe Ippolito, Benjamin Lelouvier, Alain Roulet, Fabrice Andre, Fabrice Barlesi, Jean-Charles Soria, Caroline Pradon, Emmanuelle Gallois, Fanny Pommeret, Emeline Colomba, Florent Ginhoux, Suzanne Kazandjian, Arielle Elkrief, Bertrand Routy, Makoto Miyara, Guy Gorochov, Eric Deutsch, Laurence Albiges, Annabelle Stoclin, Bertrand Gachot, Anne Florin, Mansouria Merad, Florian Scotte, Souad Assaad, Guido Kroemer, Jean-Yves Blay, Aurélien Marabelle, Frank Griscelli, Laurence Zitvogel, Lisa Derosa

## Abstract

Patients with cancer are at higher risk of severe coronavirus infectious disease 2019 (COVID-19), but the mechanisms underlying virus-host interactions during cancer therapies remain elusive. When comparing nasopharyngeal swabs from cancer and non-cancer patients for RT-qPCR cycle thresholds measuring acute respiratory syndrome coronavirus-2 (SARS-CoV-2) in 1063 patients (58% with cancer, 89% COVID-19^+^), we found that malignant disease favors the magnitude and duration of viral RNA shedding concomitant with prolonged serum elevations of type 1 IFN that anticorrelated with anti-RBD IgG antibodies. Chronic viral RNA carriers exhibited the typical immunopathology of severe COVID-19 at the early phase of infection including circulation of immature neutrophils, depletion of non-conventional monocytes and a general lymphopenia that, however, was accompanied by a rise in plasmablasts, activated follicular T helper cells, and non-naive Granzyme B^+^ FasL^+^, Eome^high^TCF-1^high^, PD-1^+^CD8^+^ Tc1 cells. Virus-induced lymphopenia worsened cancer-associated lymphocyte loss, and low lymphocyte counts correlated with chronic SARS-CoV-2 RNA shedding, COVID-19 severity and a higher risk of cancer-related death in the first and second surge of the pandemic. Lymphocyte loss correlated with significant changes in metabolites from the polyamine and biliary salt pathways as well as increased blood DNA from Enterobacteriaceae and Micrococcaceae gut family members in long term viral carriers. We surmise that cancer therapies may exacerbate the paradoxical association between lymphopenia and COVID-19-related immunopathology, and that the prevention of COVID-19-induced lymphocyte loss may reduce cancer-associated death.

## Introduction

Severe acute respiratory syndrome coronavirus-2 (SARS-CoV-2) is a novel beta-coronavirus that has caused a worldwide pandemic of the human respiratory illness COVID-19, resulting in a severe threat to public health and safety worldwide. Because of age, gender, cancer-associated risk factors, metabolic syndrome and side effects induced by their specific therapies (such as cardiomyopathy, systemic immunosuppression and cellular senescence), cancer patients appear more vulnerable to severe infection than individuals without cancer (Derosa et al., 2020). Indeed, a high hospitalization and mortality rates of SARS-CoV-2 infection were heralded in patients with malignancy in several studies across distinct geographical sites (Albiges et al., 2020; Assaad et al., 2020; Luo et al., 2020; Rugge et al., 2020). Cancer types, performance status and stage are additional risk factors for severe COVID-19 in this patient population. Patients with hematological, lung and breast cancer have been reported to be more susceptible to hospitalization or death due to COVID-19 as compared to patients with other malignancies (Garassino et al., 2020; Luo et al., 2020; Martín-Moro et al., 2020; Passamonti et al., 2020; Rugge et al., 2020). Patients diagnosed with metastatic cancers are more vulnerable to severe forms of COVID-19 than individuals with localized malignancies (Robilotti et al., 2020). Recent (<3 months) cancer treatments including surgery, chemotherapy and immunotherapy independently contribute to worsen the prognosis of COVID-19 among patients with malignant disease (Albiges et al., 2020; Dai et al., 2020; Luo et al., 2020; Passamonti et al., 2020; Q et al., 2020; Robilotti et al., 2020).

Here we explored several independent cohorts of cancer patients diagnosed with COVID-19 (1063 patients, 58% with cancer) during the first surge of the pandemic to analyze the dynamics between host (blood immunology, metabolism, metagenomics) and viral parameters and validated the most clinically relevant findings in the second surge of the pandemic. We concluded that virus-induced or -associated lymphopenia that coincided with T cell exhaustion, abnormalities in polyamine and biliary salt pathways and circulation of Enterobacteriaceae and Micrococcaceae bacterial DNA, is a dismal prognosis factor in cancer patients, likely participating in the vicious circle of immunosuppression-associated chronic virus shedding.

## Results

### Prolonged viral shedding and higher viral loads in cancer patients compared with cancer free COVID-19^+^ patients

To explore the clinical significance of viral and/or immunological parameters in cancer patients, we gathered the data from electronic clinical files from various cancer centers or general hospitals across France and Canada, in order to monitor the magnitude and duration of virus RNA shedding in nasopharyngeal swabs according to cancer (*versus* health care workers (HCW) or COVID-19^+^ cancer free individuals), tumor types (hematological *versus* solid malignancies) and staging (localized, locally advanced, metastatic diseases) (Figure 1A).

**Figure 1.**
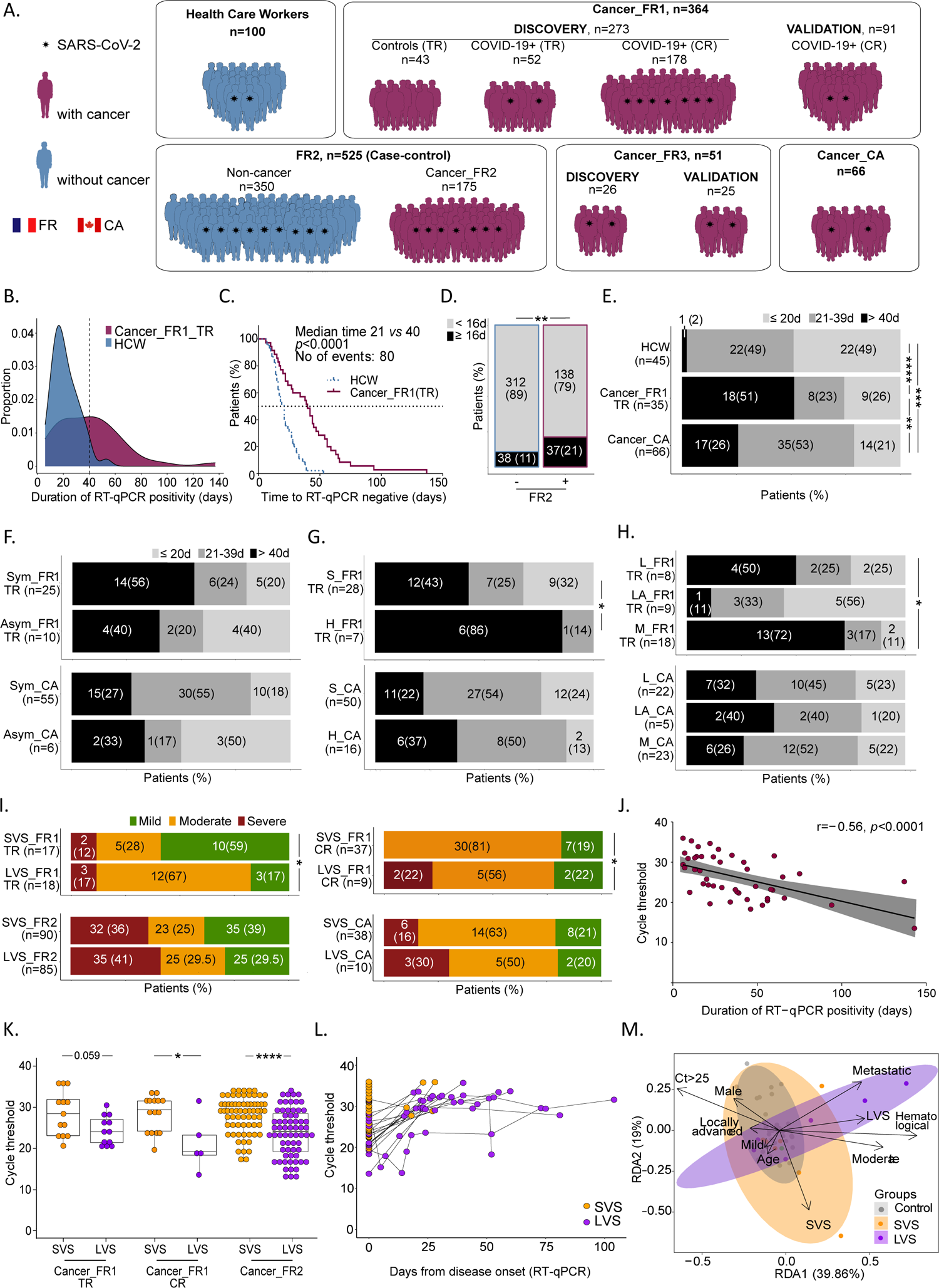
Prolonged duration of SARS-CoV-2 RNA shedding correlated with high viral load and COVID-19 severity in patients with cancer. **A.** Graphical schema of cohorts and patients’ accrual. **B.** Proportion of patients with cancer from translational research (TR) (Cancer_FR1_TR, n=35, magenta area) or healthcare workers (HCW, n=45, blue area) by days of RT-qPCR positivity. Vertical dashed line at 40 days represents the 95^th^ percentile of HCW and the median of positivity of patients with cancer. **C.** Kaplan Meier curves of time to negative RT-qPCR in HCW (n=45, blue dotted lines) and patients with cancer (Cancer_FR1_TR, n=35, magenta continuous lines). **D.** COVID-19+ cancer bearing or history of cancer (+) and cancer-free (-) individuals from FR2 treated with hydroxychloroquine +/-azithromycine: number (percentages) of patients with RT-qPCR positivity beyond 16 days (90^th^ percentile of the cancer-free population of FR2). **E.** Number (percentages) of HCW, Cancer_FR1 patients (Cancer_FR1_TR) or Canadian patients with cancer (Cancer_CA) with short, intermediate (grouped in short term viral RNA shedding, SVS) and prolonged (long term viral RNA shedding, LVS) viral RNA shedding (**E**), according to the presence/absence of viral symptoms (symptomatic, Sym, *vs* asymptomatic, Asym) (**F**), diagnosis of hematological (H) versus solid (S) malignancy (**G**), and cancer staging (localized (L), locally advanced (LA), metastatic (M)) (**H**). **I.** Number (percentages) of Cancer_FR1 patients (from translational research and clinical routine), Cancer_FR2 patients (Cancer_FR2) or Canadian patients with cancer (Cancer_CA) divided in SVS and LVS and regarding their respective COVID-19 severity. **J.** Spearman correlation between Cycle threshold (Ct) for the RT-qPCR amplification of genes encoding proteins of SARS-CoV-2 replication-transcription complex at diagnosis and duration of RT-qPCR positivity for Cancer_FR1 (from translational research and clinical routine), each dot representing one sample/patient. **K-L**. Ct values for the RT-qPCR amplification of genes encoding proteins of SARS-CoV-2 replication-transcription complex in nasopharyngeal swabs performed at diagnosis in SVS *versus* LVS in Cancer_FR1_TR and CR and Cancer_FR2 (**K**), and dynamics over time from day 0 up to day 80 after inclusion in SVS (n=33 samples, n=28 patients, orange dots) *versus* LVS (57 samples, n=17 patients, purple dots) in Cancer_FR1 (from translational research and clinical routine) (**L**). Each line represents one patient. **M.** Redundancy statistical analysis (RDA) of the cancer and viral related-clinical factors accounting for the variance of SARS-CoV-2 viral shedding status. Clinical components were influenced by the virus shedding (SVS *versus* COVID-19 negative, *p*=0.037; LVS *versus* COVID-19 negative, *p*=0.0010), COVID severity (mild *versus* COVID-19 negative, *p*=0.0030; moderate *versus* COVID-19 negative, *p*=0.0574; severe *versus* COVID-19 negative, *p* not computable), age (*p*=0.0514), hematological rather than solid malignancy (hematological *versus* solid, *p*=0.001), metastatic status (*p*=0.0059), and Ct values at diagnosis ( 25 *versus* < 25, *p*=0.0738). Chi-square tests with \**p*<0.05, \*\**p*<0.01, \*\*\**p*<0.001, \*\*\*\**p*<0.0001.

First, we conducted a prospective epidemiological study named Cancer_FR1_Translational Research (TR) at Gustave Roussy, Villejuif, France, during the first surge of the COVID-19 pandemic (from April 10, 2020 to May 11, 2020, NCT04341207) to evaluate the prevalence and severity of COVID-19 in all adult patients under treatment or recently treated for solid tumors or hematological malignancies (Figure S1, Table S1). Our secondary endpoint was the identification of viral, immunological, metabolic and metagenomics blood predictors of severe complications among cancer patients. Clinical characteristics were collected from electronic medical records (Table S1). Nasopharyngeal samples were serially collected at every hospital visit motivated by the cancer management or any symptomatology related to seasonal flu or COVID-19 and transferred to the virology laboratory for SARS-CoV-2-specific quantitative reverse transcription-PCR (RT-qPCR) testing. Out of 473 patients enrolled in Cancer_FR1_TR, 53 (11%) were diagnosed with COVID-19 by RT-qPCR, and this diagnosis was corroborated by a specific serology in 87% cases (Figure S1A). Among the 52 patients evaluable for translational research, 37% were males, 60% suffered from at least one of the co-morbidities associated with coronavirus pandemic, such as hypertension (58%) or obesity (21%) (Table S1). Seventy seven percent had an ECOG performance status of 0-1 at the time of nasopharyngeal sampling. Twenty one percent of COVID-19 positive cancer patients did not report any symptom of infection, 61% required hospital admission (for any cause or because of COVID-19 aggravation within 28 days after diagnosis) and 11% a transfer to intensive care unit (ICU), culminating with cancer death in 7% of the cases (from undetermined cause, no systematic necropsy) (Figure S1B-G, Table S1). Among patients with cancer diagnosed with COVID-19, 20% were followed up for hematological (as opposed to solid) malignancies and developed more severe symptoms of infection (Figure S1B-G, Table S1). In the Cancer_FR1_TR study, 33%, 21% and 46% presented with localized, locally advanced and metastatic disease, respectively that were equally susceptible to severe COVID-19 (Figure S1F-G).

Given that cycle threshold (Ct) values of the first RT-qPCR test may be correlated with the clinical characteristics of the patients (Shlomai et al., 2020; Westblade et al., 2020), we performed a longitudinal follow-up of Ct values by RT-qPCR. We targeted several genes coding for the envelope, the nucleocapsid and/or the replication-transcription complex (RdRP, Orf1a, subgenomic RNA of the SARS-CoV-2 (Corman et al., 2020)(Wölfel et al., 2020)) to assess the duration of the nasopharyngeal SARS-CoV-2 RNA shedding, starting at COVID-19 diagnosis for up to 6 months as per-protocol indications (Figure S2A). The duration of viral shedding was defined as the number of days from the first positive to the first negative RT-qPCR, after longitudinal monitoring with an interval inferior to 40 days, to reduce bias in viral shedding estimation. This time lapse of 40 days corresponded to the median of SARS-CoV-2 virus carriage in the cancer population (Figure 1B-C, Table S1). In parallel, a similar and systematic COVID-19 protocol with longitudinal RT-qPCR testing was applied to health care workers (HCW) at Gustave Roussy. Health care workers had a mean age of 35 years (range: 19-61), were mostly females (male *versus* female: 13% *versus* 87%), and presented with one or two co-morbidities in 27% and 4%, respectively, thereby significantly diverging from the cancer population diagnosed with COVID-19. Starting from 50 COVID-19 positive cancer patients and 100 HCW, we conducted RT-qPCR in 210 and 200 nasopharyngeal swabs, respectively (Figure S2). However, applying the exclusion criteria detailed in Figure S2, we could compare the median length of SARS-CoV-2 RNA detection in 35 cancer patients (Cancer_FR1_TR) and 45 HCW using 168 and 118 samples, respectively. Patients with cancer exhibited prolonged nasopharyngeal RNA virus shedding (Figure 1B, median of 40 days (range: 6-137) for patients with cancer compared to 21 days (range: 7-53) for HCW, Figure 1C, log-rank test *p*-value <0.0001). This difference persisted after adjusting for age, gender and co-morbidities (Cox multivariate analysis, adjusted hazard ratio [95% confidence interval] =2.88 [1.42;5/85], *p*=0.00291, Figure 1C). To further validate the differences observed in the duration of viral RNA shedding between Cancer_FR1_TR and HCW, we analyzed another cohort of patients diagnosed with COVID-19 in a general hospital from Southern France and paired - in a case-control study - 175 cancer patients (with a history of cancer or currently treated with cancer (Table S1)) with 350 cancer free individuals based on age, gender, comorbidities and COVID-19 severity (FR2_Case-Control, Cancer and Non-Cancer) (Figure 1A,Table S1). Here again, there was a prolonged length of RT-qPCR positivity in cancer individuals compared with cancer free COVID-19 patients (8 days *versus* 6 days, log-rank test *p*-value, *p*=0.03), taking into account that >70% were treated with hydroxychloroquine and azithromycine, a combination regimen reducing viral shedding (Lagier et al., 2020). Moreover, the proportion of patients with a viral shedding above 16 days (corresponding to the 90^th^ percentile of the viral shedding in cancer-free patients) was higher in cancer patients (Figure 1D, *p*<0.0015). A second independent validation was achieved in a third series of 66 patients with cancer extracted from a cohort of 252 cancer individuals living in Canada and diagnosed with COVID-19 (Cancer_CA), for whom a longitudinal SARS-CoV-2-specific RT-qPCR (using *Orf1* and *E* gene probe sets (Boutin et al., 2020)) follow-up had been carried out (Elkrief et al., 2020) (Figure 1A, Table S1). Here again, we observed that 26% of cancer patients were still PCR positive after 40 days from diagnosis by RT-qPCR (Figure 1E). Such a long term PCR detection of viral RNA could indicate stable subgenomic RNA contained within double membrane vesicles or presence of a replicative mucosal viral strain. Hence, we confirmed in three independent series of cancer patients a prolongation of RNA virus shedding previously described in case reports in hematological or immuno-compromised patients (Avanzato et al., 2020; Aydillo et al., 2020; Choi et al., 2020; Helleberg et al., 2020).

Hence, we focused on the differential characteristics of cancer patients presenting with Long term Viral RNA Shedding (LVS), defined by a positive RT-qPCR duration ≥ 40 days (median of RT-qPCR duration in Cancer_FR1_TR (Figure 1C)), compared to those experiencing Short term Viral RNA Shedding (SVS), defined by a positive RT-qPCR duration < 40 days henceforth (Table S1). The increased susceptibility to develop a LVS was independent of initial symptomatology, observed in 33% of Canadian (CA) to 40% of French (FR1_TR) asymptomatic and 27% (CA) to 56% (FR1_TR) of symptomatic cancer patients (Figure 1F). There was a higher propensity to LVS in hematological malignancies compared to solid cancers (86% *versus* 43%, respectively (*p*=0.04, Figure 1G, Table S1) and in advanced disease (*p*=0.011) in FR1_TR cohort (Figure 1H, Table S1) but less so, in the CA cohort. Importantly, the LVS phenotype was associated with COVID-19 severity, notably an increased risk to develop a moderate form (defined by thoracic CT scan, hospitalization and oxygen requirement < 9 L/min) in Cancer_FR1_TR (*p*=0.032) (Figure 1I). This trend was confirmed in a third series of French patients from the clinical routine (CR) managed outside the translational ancillary study at Gustave Roussy (called henceforth “Cancer_FR1_CR” Table S1, Figure S3), where 20% cancer patients were diagnosed with LVS and exhibited more severe COVID-19 infections (Figure 1I, *p*=0.011). Again, the hospitalization rates and transfer to intensive care units were increased in LVS compared with SVS patients in Cancer_FR1_TR (*p*=0.0018, Table S1) and Cancer_FR2, respectively (*p*=0.02, Table S1). Finally, the FR2 and Canadian series of LVS cancer patients also tended to exhibit more severe manifestations of COVID-19 compared with SVS Canadian cancer patients (Figure 1I, bottom).

Of note, the duration of viral RNA shedding correlated with “viral load”, *i.e* Ct values at diagnosis, in that cancer patients with LVS experienced lower Ct values at diagnosis than SVS cancer patients in most cohorts for which the data were available (Figure 1J). Importantly, cancer patients doomed to develop LVS presented with lower Ct values at diagnosis than those prone to become SVS in Cancer_FR1 and Cancer_FR2 cohorts (Figure 1K). The dynamic course of the infection was significantly different in SVS and LVS, as indicated by the Ct values that remained high for a prolonged period of time in LVS patients compared with SVS (Figure 1L). Of note, Ct values at disease onset were significantly anticorrelated with duration of viral RNA shedding in cancer patients using either N or *Orf1ab*/*RdRP* gene-specific probe sets.

The redundancy analysis (RDA) is an extension of the principal component analysis (PCA) aimed at identifying viral components which depend on other known covariates such as clinical parameters. RDA revealed that, within 30 days from diagnosis, 18% of the variance of the biological parameters are explained by 10 components adjusted for the major clinical parameters for COVID-19 in Cancer_FR1_TR (Figure 1M). These components were mainly influenced by the virus shedding (SVS *versus* COVID-19 negative, *p*=0.037; LVS *versus* COVID-19 negative, *p*=0.0010), COVID severity (mild *versus* COVID-19 negative, *p*=0.0030; moderate *versus* COVID-19 negative, *p*=0.0574; severe *versus* COVID-19 negative, *p* not computable), age (*p*=0.0514), hematological rather than solid malignancy (hematological *versus* solid, *p*=0.001), metastatic status (*p*=0.0059), and Ct values at diagnosis (> 25 *versus* < 25, *p*=0.0738). As outlined in Table S1, LVS patients tended to be older (66 *versus* 56 years old, *p*=0.08), more metastatic (72% *versus* 29%, *p*=0.01), and experienced increased hospitalization rates (83% *versus* 23%, *p*<0.001) than SVS cancer patients in the Cancer_FR1_TR cohort.

### Immunological hallmarks of long-term virus carriers at diagnosis

Intrigued by these findings, we addressed the question as to whether and how prolonged viral RNA shedding would impact on Cancer_FR1_TR patients with respect to COVID-19-related immunological alterations previously reported for cancer-free infected individuals (Arunachalam et al., 2020; Chua et al., 2020; Kaneko et al., 2020; Laing et al., 2020; Mathew et al., 2020; Silvin et al., 2020; Takahashi et al., 2020). More than 80 phenotypic markers were quantified on circulating leukocytes by means of high dimensional spectral flow cytometry, complemented by multiplex ELISAs to detect serum chemokines, cytokines and growth factors. These parameters were recorded within or after the first 20 days of inclusion in the Cancer_FR1_TR protocol, for 25 COVID-19^+^ cancer patients that were divided into LVS *versus* SVS subgroups, in comparison to 43 COVID-19 negative cancer patients (“controls” or “Ctls”) matched for age, gender, co-morbidities, cancer types and tumor extension (Table S2). Asymptomatic individuals and cancer patients enrolled at the recovery phase of COVID-19 (meaning that they became PCR negative) were analyzed separately. Within the first 20 days from diagnosis, LVS presented increased proportions of monocytes among circulating leukocytes (Figure S4A, left panel), and a parallel drop in CD169^-^ A-DR^+^ within conventional monocytes (Figure S4A, middle panel) and in non-conventional monocytes (CD1 ^+^CD1 ^low/-^ Figure S3A right panel) compared to SVS, cancer controls, asymptomatic or recovered patients, as reported (Carvelli et al., 2020; Silvin et al., 2020). Polymorphonuclear cells (PMN) tended to increase in LVS, specifically immature CD10 ^+/-^ 10^+/-^ 16^-^ neutrophils, compared with SVS, convalescent and controls (Figure 2A-B, upper and lower panels, Figure S4B).

**Figure 2.**
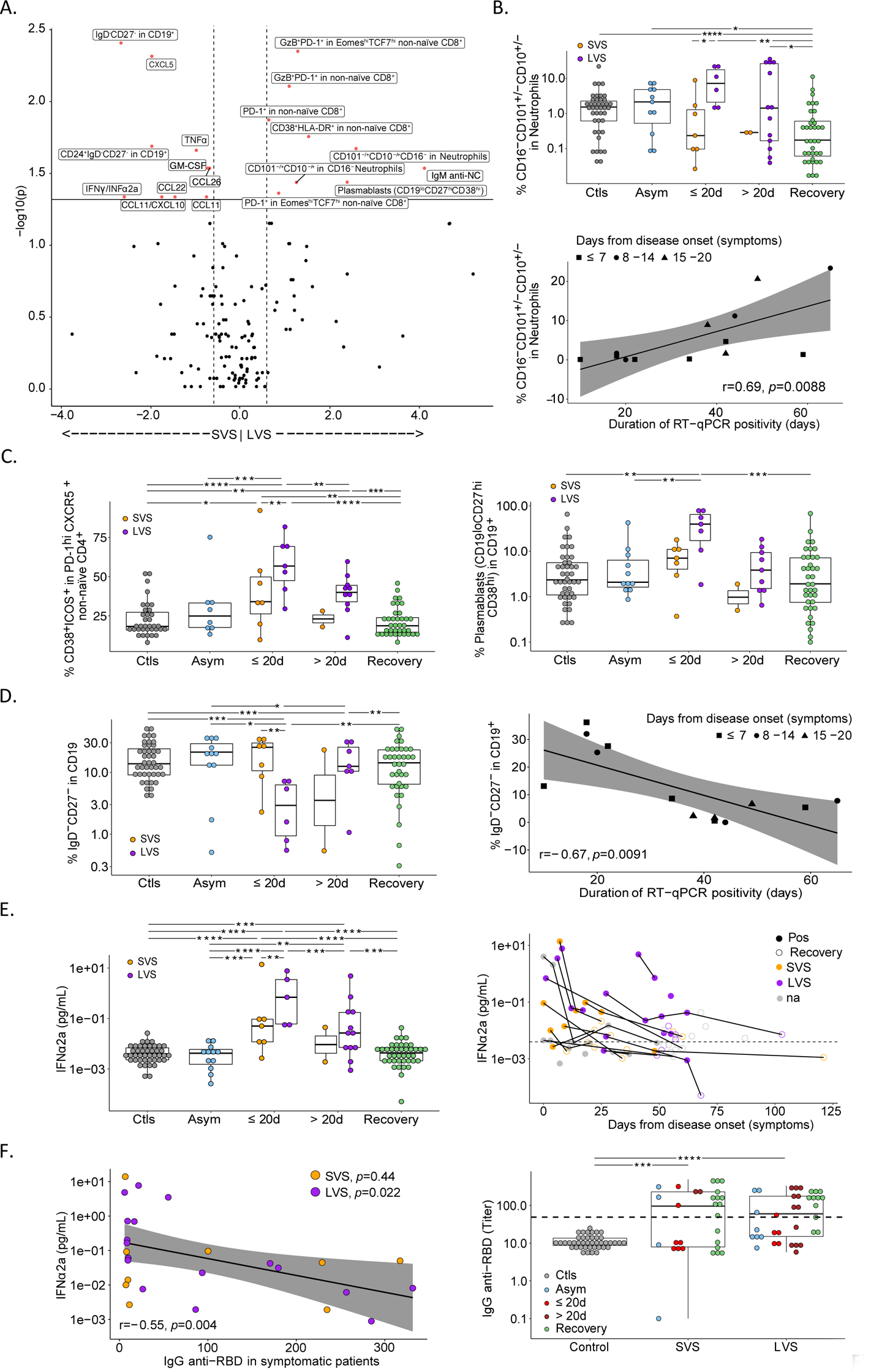
Immunotypes associated with prolonged viral RNA shedding in patients with cancer. **A.** Volcano plot of the differential cellular and soluble immune parameters contrasting short term viral RNA shedding (SVS) *versus* long term viral RNA shedding (LVS) during the first 20 days of symptoms. Volcano plot was generated computing for each immune factor: i) the log2 of fold change among the mean relative percentages after normalization in SVS *versus* LVS (x axis); ii) the log10 of *p* values deriving from Wilcoxon test calculated on relative percentages in absolute values (y axis). Black and red dots are considered non-significant (*p*<0.05) or significant (*p*>0.05), respectively. **B-F.** Temporal changes and correlation of blood leukocyte parameters measured by high dimensional spectral flow cytometry (**B-D**) and soluble factors IFNα2a and anti-SARS-CoV-2 IgG (**E, F**) in various phases of COVID-19 presentation (no virus infection (Ctls, grey dots), asymptomatic viral infection (Asym, light blue dots), symptomatic viral infection examined in the first 20 days (≤20d) or after 20 days (>20d) of symptoms with those experiencing Short term Viral RNA Shedding (SVS, orange dots) or Long term Viral RNA Shedding (LVS, purple dots) and RT-qPCR negative COVID-19 patients in the convalescent phase (Recovery, green dots or circled dots) (supplementary material, Figure 1). Box plots display a group of numerical data through their 3^rd^ and 1^st^ quartiles (box), mean (central band), minimum and maximum (whiskers). Each dot represents one sample, each patient being drawn 1-3 times. Statistical analyses used one-way ANOVA with Kenward-Roger method to take into account the number of specimen/patient: \**p*<0.05, \*\**p*<0.01, \*\*\**p*<0.001, \*\*\*\**p*<0.0001. **B-D.** Percentages of neutrophils that do not express either CD101 and/or CD10 and lost CD16 within the gate of CD45^+^CD56^-^CD3^-^CD19^-^CD15^+^ cells (**B, upper panel**). Spearman correlation between the percentage of immature neutrophils (CD10^+/-^CD101^+/-^CD16^-^) measured within the first 20 days of symptoms with the duration of SARS-CoV-2 RT-qPCR positivity (**B, lower panel**). **C-D**. Percentages of CD38^+^ICOS^+^ among CXCR5^+^PD-1^+^ non-naive CD4^+^ (**C, left panel**), plasmablasts defined as CD19^low^CD38^high^CD27^+^ within the CD19^+^ gate (**C, right panel**), double negative IgD^-^CD27^-^ among CD19^+^ cells (**D, left panel**) and their Spearman correlation when measured within the first 20 days of symptoms with the duration of SARS-CoV-2 RT-qPCR positivity (**D, right panel**). **E.** Ultrasensitive electrochemiluminescence assay to monitor the serum concentrations of IFNα2a (**E, left panel**) in a kinetic fashion (**E, right panel**). Each line and dot represent one patient and one sample, respectively and dashed line represents the median value of controls. **F.** Spearman correlation between the serum IFNα2a values measured in symptomatic patients with IgG titers against SARS-CoV-2 S1-RBD considered as continuous variables (**F, left panel**). The raw data are represented in the **right panel** at both time points for each group of patients.

Importantly, the most significant phenotypic traits distinguishing LVS from SVS featured among the reported hallmarks of severe COVID-19 in cancer-free subjects (Arunachalam et al., 2020; Chua et al., 2020; Kaneko et al., 2020; Laing et al., 2020; Mathew et al., 2020; Silvin et al., 2020; Takahashi et al., 2020) (Figure 2A). In accordance with the reported defects in germinal center formation in secondary lymphoid organs of severe COVID-19 (Kaneko et al., 2020), LVS cancer patients exhibited increased recirculation of activated CXCR5^+^PD-1^high^ CD4^+^ follicular T helper cells (TFH) expressing ICOS and CD38 (Figure 2C, left panel), as well as a marked rise in plasmablasts (defined as CD19^low^ CD27^hi^CD38^hi^) at the expense of transitional B (CD24^+^ 38^hi^CD19^+^) and double negative B cells (IgD^-^CD27^-^CD19^+^) (Figure 2C right panel, Figure S4C, Figure 2D). As indicated in the Volcano plot in Figure 2A, immature PMN and double negative B cells were among the most significant immunological features, positively and negatively predicting LVS, respectively (Figure 2B bottom panel and Figure 2D right panel). LVS coincided with prolonged systemic release of, and exposure to, type 1 IFN above levels measured in SVS, controls and recovered individuals (Figure 2E). Type 1 IFN levels anticorrelated with titers of neutralizing anti-S1 RBD antibodies (Figure 2F). This landscape of immune profiling was corroborated by non-supervised hierarchical clustering of innate and cognate immunotypes and serum cytokine concentrations analyzed within 30 days from diagnosis. This method allowed to segregate a small cluster of individuals characterized by low Ct values (< 25), and moderate/severe complications of COVID-19, that included metastatic cancer carriers with LVS or SVS (Figure S5). This cluster was separated from the others by typical signs of viral infection, including abundant circulating CD38^+^HLA-DR^+^CD8^+^T cells, plasmablasts, activated TFH cells and high serum IFNα2a levels (Figure S5). Likewise, while many inflammatory cytokines, chemokines or alarmins (such as IFNγ, CXCL10, IL-4, IL-6 and calprotectin) were elevated in symptomatic COVID-19 individuals compared with controls, asymptomatic and recovered patients, none of them could predict LVS, except a drop in the IFNγ/IFNα2a and CCL11/CXCL10 ratios whose significance remains unclear (*p*=0.016 and *p*=0.0019, respectively) (Figure S4D-I). Interestingly, innate and cognate immunotypes performed in convalescent patients and controls segregated at random in the non-supervised hierarchical clustering (Figure S6).

Altogether, the high dimensional flow cytometry of blood immune subsets indicated that LVS cancer patients harbored the immunological hallmarks of severe COVID-19 at diagnosis.

### Virus-associated lymphopenia predicted shorter overall survival in the first and second surge of the pandemic

Lymphocyte loss is a feature of severe COVID-19 in non-cancer patients (Laing et al., 2020; Mathew et al., 2020). Not surprisingly, blood absolute lymphocyte counts (ALC) at diagnosis anticorrelated with the duration of PCR positivity in Cancer_FR1_TR and Cancer_FR1_CR cohorts (Figure 3A). However, although the ALC before the COVID-19 pandemic (blood drawn from December 2019 to mid-March 2020) were already somewhat lower in LVS than in SVS cancer patients, the ALC during the outbreak dramatically dropped in cancer patients doomed to develop LVS (in both Cancer_FR1_TR and Cancer_FR1_CR cohorts), more so than in individuals prone to SVS (Figure 3B, left panel). The extent in ALC reduction was more severe in patients presenting LVS than SVS (Figure 3C). Of note, ALC recovered in both patient groups regardless of the LVS/SVS status, supporting the contention that reduced ALC at COVID-19 diagnosis is induced by the virus rather than by the cancer (Figure 3B, left panel). In accord with the finding that LVS correlates with high viral load at symptom onset (Figure 1J-K), higher viral loads at diagnosis were associated with a pronounced COVID19-associated lymphopenia (Figure 3B, right panel).

**Figure 3.**
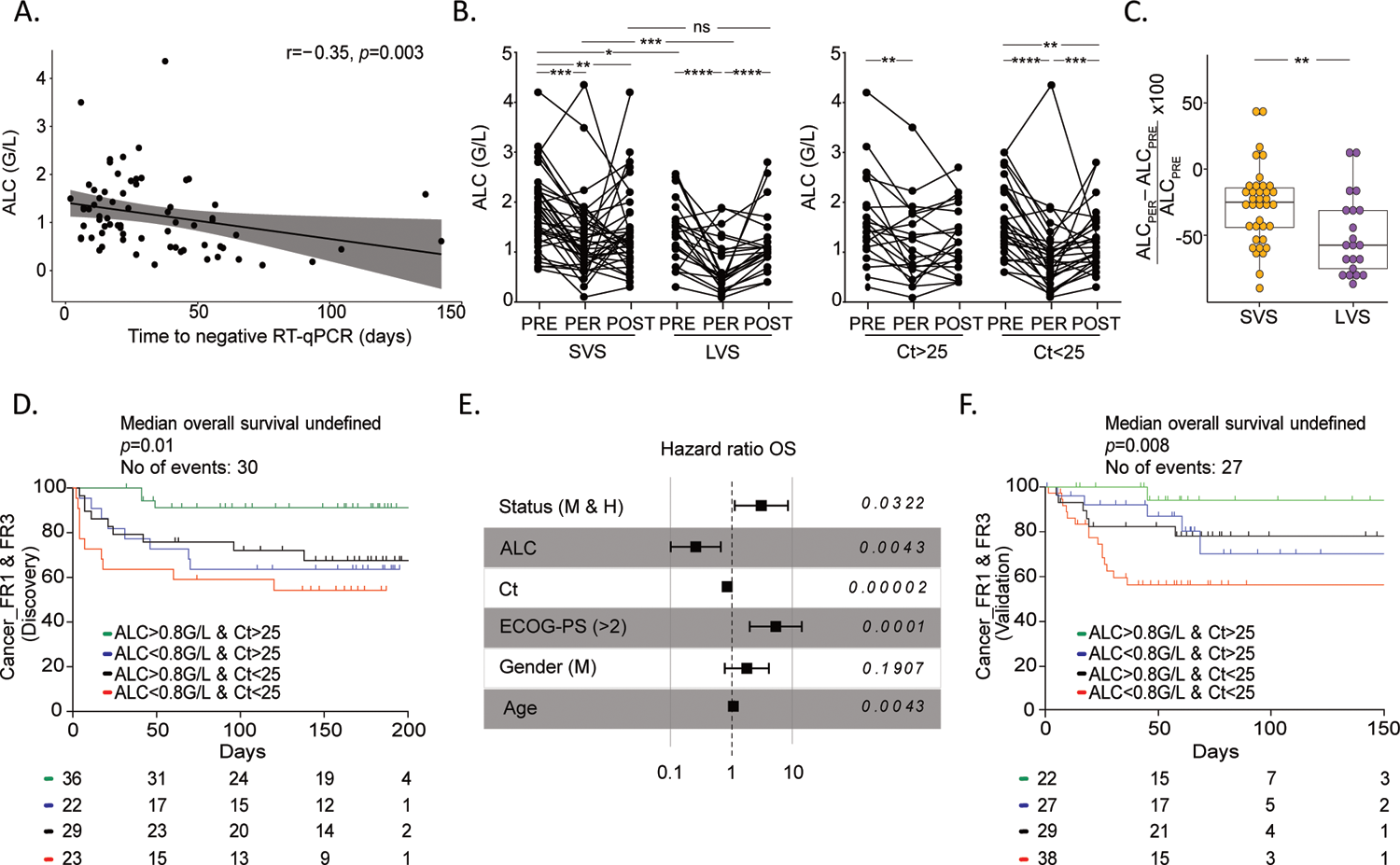
Lymphopenia and high viral load are dismal prognosis factors for overall survival in cancer patients in the first and second surge of the pandemic. **A.** Spearman correlation between the absolute lymphocyte counts (ALC) of Cancer_FR1 (from translational research and clinical routine), with the duration of SARS-CoV-2 RT-qPCR positivity (only evaluable patients for both factors, n=69 patients). **B-C.** ALC of Cancer_FR1 (from translational research and clinical routine) in SVS (n=37 patients) *versus* LVS (n=22 patients) subsets (**B, left panel**) or SARS-CoV-2-cycle threshold (Ct) > 25 (n=21 patients) *versus* Ct < 25 (n=29 patients) (**B, right panel)** monitored during the COVID-19 pandemic (“PER”, between −4 and +7 days of the disease diagnosis by RT-qPCR), between 210 and 12 days before the symptom onset of COVID-19 (“PRE”) or within the recovery period (between 0 and 123 days after negative RT-qPCR) (“POST”) at Gustave Roussy, with calculation of the reduction between “PRE” and during COVID-19 (**C**). One patient defined as outlier (at 215%) by ROUT method was excluded from the LVS group for the analysis. Each line and dot represents one patient and one sample. Statistical analyses used one-way ANOVA (paired and unpaired) with Kenward-Roger method taking into account the number of specimen/patient (B): \**p*<0.05, \*\**p*<0.01, \*\*\**p*<0.001, \*\*\*\**p*<0.000 and Mann-Whitney (C): \*\**p*<0.01. **D**. Kaplan Meier curve and Cox regression analysis of overall survival of cancer patients from the Discovery (1st surge) cohort (Cancer_FR1+Cancer_FR3), all stages included, according to ALC and Ct value at diagnosis. Refer to Table 1 for patient characteristics. **E.** Multivariate Cox regression analysis stratified for the cohort and adjusted for age, ECOG status, gender and metastatic and/or hematological status of cancer patients from the Discovery (1st surge) cohort (Cancer_FR1+Cancer_FR3). **F**. Kaplan Meier curve and Cox regression analysis of overall survival of cancer patients from Validation (2nd surge) cohort (Cancer_FR1+Cancer_FR3), all stages included, according to ALC and Ct value at diagnosis. Refer to Table 1 for patient characteristics.

We next assessed the clinical significance of the interaction between Ct values, ALC and cancer patient survival in 110 cancer patients with COVID-19 (Discovery cohort (first surge of the pandemic) including 84 patients from Cancer_FR1 treated at Gustave Roussy and 26 patients from Cancer_ FR3 treated at Léon Bérard Cancer Center in Lyon, France) (Figure 3D, Table 1). Cox logistic regression analyses and Kaplan Meier survival curves were performed after stratification of the patients according to both, Ct and ALC values at diagnosis. The cut-off for the Ct value was 25 and corresponded to the median of the whole cohort FR1+FR3, which coincided with the threshold at which live virus particles can be isolated in 70% of the cases (Jaafar et al., 2020). The cut-off value for ALC was the median found in patients with high viral load (Ct < 25) at diagnosis (ALC=800/mm^3^). ALC combined with Ct values predicted cancer-related overall survival in univariate analyses across all cancer stages (local, locally advanced or metastatic) (Figure 3D, Table 1). While patients presenting with ALC > 800/mm^3^ and low viral load (Ct > 25) exhibited prolonged survival, a dismal prognosis affected 21% of them (23/110) who presented both deep lymphopenia (ALC < 800/mm^3^) and high viral loads (Ct < 25) at diagnosis (Figure 3D) culminating in 40% deaths at 3 months. All 4 groups were comparable in terms of age, gender, co-morbidities, cancer type or staging (Table 1). Multivariate Cox analysis stratified for the cohort origin and adjusted for age (hazard ratio [95% confidence interval]=1.042 [1.013; 1.072], *p=*0.0043), ECOG performance status (4.547 [1.845; 11.206], *p=*0.0001), gender (1.668 [0.775; 3.588], *p=*0.1907) and metastatic status and hematological malignancies (2.747 [1.090; 6.923], *p=*0.0322) confirmed a continuous decrease of risk with the increase of the Ct value (0.841 [0.776; 0.911], *p=*0.00002) and the increase of the ALC (0.282 [0.119; 0.672], *p=*0.0043) (Figure 3E). Of note, treatment retardation could not explain the high mortality of patients presenting with a high viral load and low ALC (Table 1).

We confirmed these predictors (ALC < 800 & Ct < 25) of poor survival during the second surge of the pandemic (between May 5^th^, 2020 to November 25^th^, 2020) in 116 new COVID-19 cancer patients (“Validation”, Cancer_FR1 and Cancer_FR3, Figure 1A). Here again, the subset of patients with ALC < 800 & Ct < 25 (n=38/116, 32.7%) exhibited the most reduced overall survival compared to the other groups with > 40% deaths at 50 days (Figure 3F). Of note, the reduced survival rate in the subset of patients defined by ALC < 800 & Ct < 25 was not a peculiarity of hematological malignancies (characterized by therapy-induced B cell depletion) since it was also observed in patients with solid neoplasia (Figure S7).

In conclusion, it appears that uncontrolled viral infection capable of compromising the number and function of circulating lymphocytes predicts lethal outcome of patients with a malignant disease.

### Immunological, metabolic and metagenomic parameters associated with virus induced-lymphocyte loss

Multiple and non-exclusive mechanisms could account for virus-associated lymphopenia (Arunachalam et al., 2020; Campbell et al., 2020; Mathew et al., 2020; Moole and Papireddypalli, 2020; Sheng et al., 2017; Song et al., 2020). To further investigate this deleterious virus-induced lymphocyte loss, we searched for the most robust correlates between ALC and immunological, metabolic or pathogenic cues in the Cancer_FR_TR cohort as well as non-cancer COVID-19 patients that we previously reported (Silvin et al., 2020).

Firstly, the Spearman correlation matrix of the main immunological and serum markers monitored at the peak of disease (within the first 20 days of disease onset) indicated close interconnections between lymphocyte proportions and their subsets within leukocytes (Figure 4A). Lymphopenia, which is a prominent feature of COVID-19 and a hallmark of severe infection, distinguished LVS from SVS or asymptomatic individuals (Figure S8A-B), as exemplified for the proportion of B lymphocytes among total CD45^+^ leukocytes after 20 days of symptoms. As reported (Mathew et al., 2020), the transitional differentiation of naive into effector/memory T cells co-expressing CD38^+^HLA-DR^+^ among CD8^+^T cells is a hallmark of COVID-19 that persisted in LVS compared to controls and SVS *(p*=0.002 and *p*=0.012) (Figure S7C, right panel). In particular, the most compelling LVS-associated T cell subpopulation that expanded in the context of lymphopenia was the non-naive (non-CD45RA^+^CD27^+^) CD8^+^ T subset expressing an activation/exhaustion phenotype characterized by early and sustained expression of PD-1 (Figure 4B), Eomes, Granzyme B, TCF-1 including the pro-apoptotic marker CD95-L (FasL) (Figure 4C-D left panel). The abundance of these non-naïve exhausted PD-1^+^CD8^+^ Tc1 cells positively correlated with the duration of SARS-CoV-2 specific RT-qPCR positivity (Figure 4B, bottom panel and Figure 4D, right panel) and may explain, at least partly, the reduced fitness and half-life of peripheral lymphocytes.

**Figure 4.**
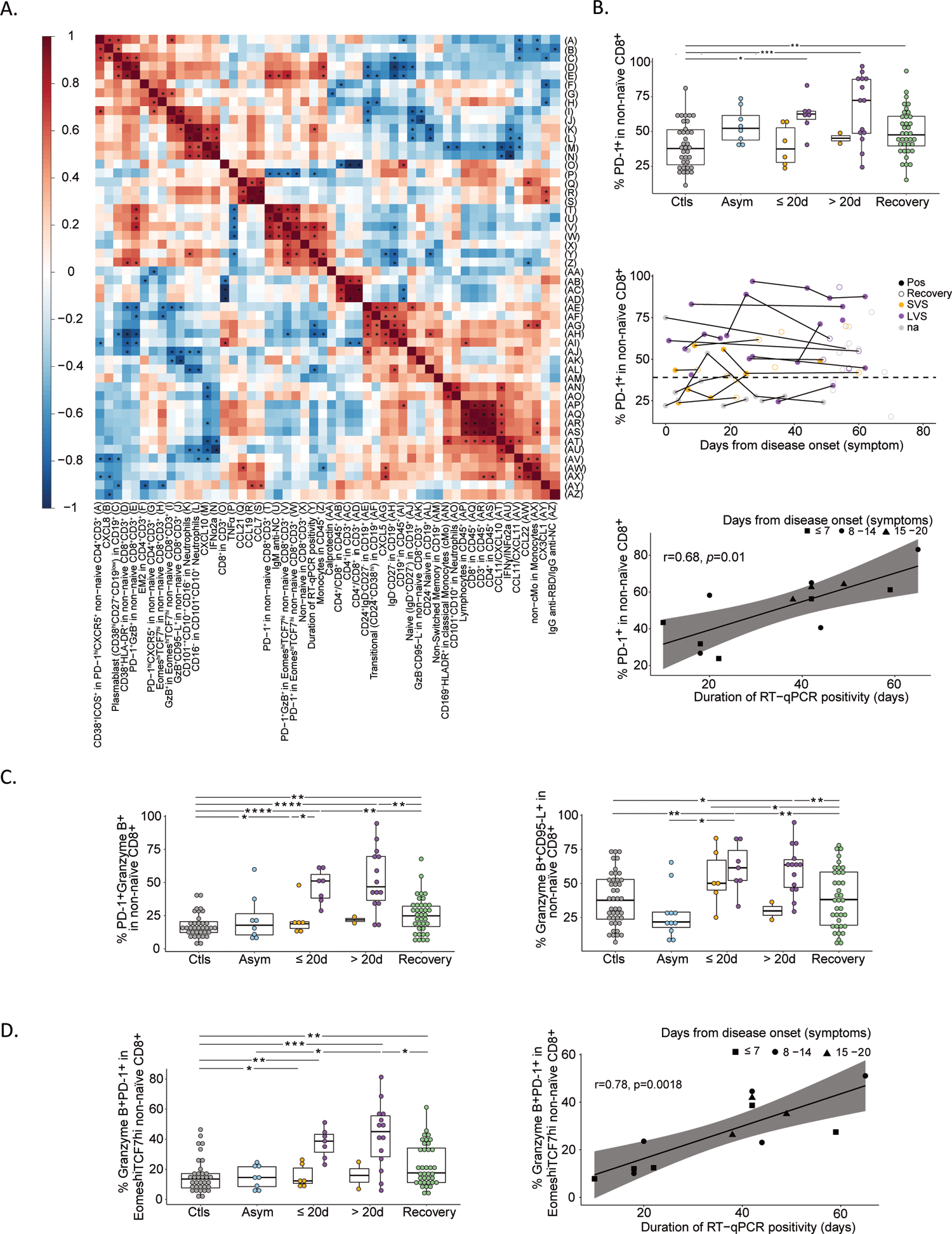
Prolonged viral shedding is associated with T cell exhaustion. **A.** Spearman correlation matrix focusing on the most significant immune variables and serum analytes monitored within the first 20 days of symptoms in patients diagnosed with COVID-19 in the Cancer_FR1_TR cohort. Stars indicate significant values (*p*<0.05) for positive (red) or negative (blue) correlations. **B.** Percentages of PD-1 expressing cells within the non-naive CD8^+^CD3^+^ population (**B, upper panel**), monitoring in various phases of COVID-19 presentation (no virus infection (Ctls, grey dots), asymptomatic viral infection (Asym, light blue dots), symptomatic viral infection examined in the first 20 days (≤ d) or after 20 days (>20d) of symptoms with those experiencing Short term Viral RNA Shedding (SVS, orange dots) or Long term Viral RNA Shedding (LVS, purple dots) and RT-qPCR negative COVID-19 patients in the convalescent phase (Recovery, green dots or circled dots) among Cancer_FR1_TR (**B, middle panel**) and Spearman correlation with the duration of SARS-CoV-2 RT-qPCR positivity measured within the first 20 days of symptoms (**B, lower panel**). **C.** Percentages of subsets co-expressing PD-1 and Granzyme B (**C, left panel**) or Granzyme B and FasL (**C, right panel**) in non-naïve CD8^+^. **D.** Percentage of PD-1^+^ and Granzyme B^+^ within the non-naïve CD8^+^ expressing Eomes^high^TCF-1^high^ gate (**D, left panel**) and Spearman correlation between this ratio measured within the first 20 days of symptoms with the duration of SARS-CoV-2 RT-qPCR positivity (**D, right panel**). Box plots display a group of numerical data through their 3^rd^ and 1^st^ quartiles (box), mean (central band), minimum and maximum (whiskers). Each dot represents one sample, each patient being drawn 1-3 times. Statistical analyses used one-way ANOVA with Kenward-Roger method to take into account the number of specimen/patients: \**p*<0.05, \*\**p*<0.01, \*\*\**p*<0.001. Each line and dot represents one patient and one sample, respectively (B, middle panel).

Secondly, we performed the serum metabolome determined by untargeted and targeted mass spectrometry-based metabolomics analyzing more than 221 metabolites in 31 cancer patients from Cancer_FR1_TR, as well as in a previously described cohort of 66 cancer-free COVID-19^+^ patients for validation (Silvin et al., 2020). The non-supervised hierarchical clustering of the serum metabolome clearly contrasted LVS from LVS patients (Figure S9). The Volcano plot aimed at identifying significant differences between LVS and SVS patients pointed out the biliary salt metabolic pathway segregating SVS from LVS serum (Figure 5A), previously described to have biological significance for lymphocyte fitness and maintenance (Campbell et al., 2020; Moole and Papireddypalli, 2020; Sheng et al., 2017; Song et al., 2020). Secondary biliary acids (such as the murideoxycholic acid (muri-DOC) (Figure 5B, left panel) and the DOC (Figure 5C)) were decreased in LVS compared with SVS and controls and correlated with lower ALC in cancer patients (Figure 5B, right panels) or severe COVID-19 (Figure 5D). Similarly, two other derivatives of DOC (hyo-DOC, urso-DOC) were decreased in LVS (compared to controls and SVS, Figure S10A-B, left panels) and were associated with lymphocyte loss (Figure S10A and S10B, right panels).

**Figure 5.**
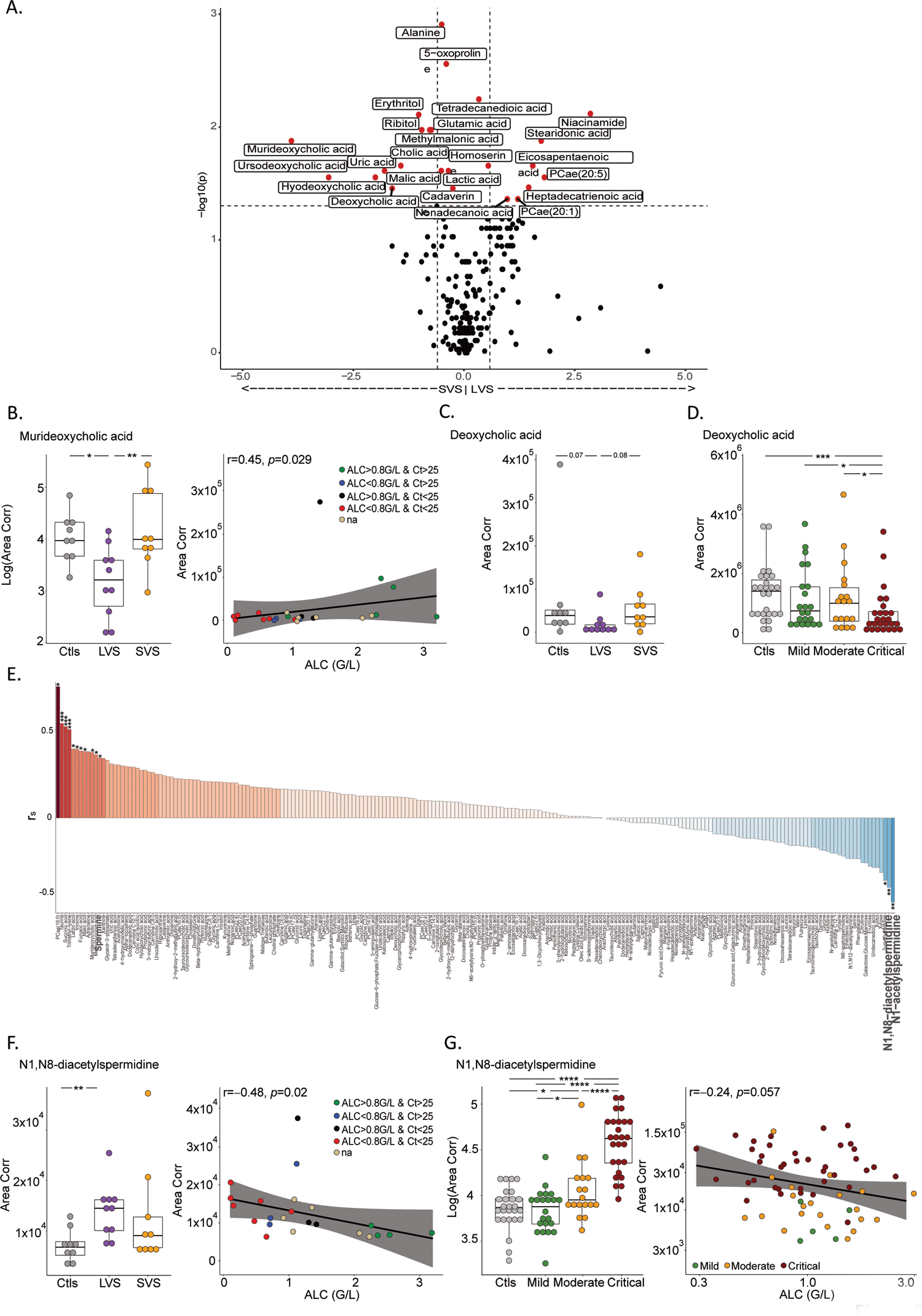
Lymphopenia and prolonged viral shedding are associated with perturbations of the polyamine and biliary acid pathways. **A.** Volcano plot identifying statistically different serum metabolites between patients experiencing Short term Viral RNA Shedding (SVS) and those experiencing Long term Viral RNA Shedding (LVS) in Cancer_FR1_TR cohort. Metabolites significantly different between both groups are in red and annotated (*p*<0.05, FC>0.5). **B.** Levels of murideoxycholic acid according to the duration of viral shedding in Cancer_FR1_TR (**left panel**) and Spearman correlation with absolute lymphocyte count (ALC) (**right panel**). The color code corresponds to the category of cycle threshold (Ct) and ALC at diagnosis. **C-D**. Serum concentrations of deoxycholic acid according to the duration of viral shedding in Cancer_FR1_TR (**C**) and the severity of COVID-19 infection in cancer free individuals (**D**). **E.** Waterfall plot of Spearman’s correlation coefficient (r_s_) between ALC and 221 metabolites in the serum of patients diagnosed positive for COVID-19. **F.** N1, N8-diacetylspermidine relative abundance in controls, SVS and LVS patients in the Cancer_FR1 cohort, that is negatively correlated with the ALC. The color code corresponds to the category of cycle threshold (Ct) and ALC at diagnosis. **G.** Levels of N1, N8-diacetylspermidine in non-cancer COVID-19 patients according the clinical severity compared to COVID19 negative controls (Ctls) (*p*<0.0001) (**G**, **left panel**), that are negatively correlated with the absolute lymphocyte count (ALC) (**G**, **right panel**). Box plots display a group of numerical data through their 3^rd^ and 1^st^ quartiles (box), mean (central band), minimum and maximum (whiskers). Each dot represents one sample, each patient being drawn once for cancer free individuals and 1-2 times for cancer patients. Statistical analyses used one-way ANOVA with Kenward-Roger method to take into account the number of specimen/patient (B, left panel, C-E, left panel): \**p*<0.05, \*\**p*<0.01), non-parametric unpaired Wilcoxon test (Mann-Whitney) for each two-group comparison: \**p*<0.05, \*\**p*< 0.01, \*\*\**p*< 0.001, \*\*\*\**p*<0.0001.

Another metabolic pathway pertaining to polyamines with high biological significance for age-related immunosenescence (Alsaleh et al., 2020; Puleston et al., 2014; Zhang et al., 2019) was also strongly associated with the duration of RT-qPCR positivity, ALC and disease severity (Figure 5E-G, Figure S9). In particular, the N1, N8-diacetylspermidine that anticorrelated with ALC (Figure 5F, right panel) increased in the serum of LVS patients (but not SVS, Figure 5F, left panel), in accordance with its marked rise in severe COVID-19 in cancer-free individuals (Figure 5G, left panel) where high levels coincided with the lymphocyte drop (Figure 5G, right panel). Of note, the tryptophane/kynurenine or lactic acid metabolites were not relevant in our study (Figure 5A, Figure S9).

Thirdly, endotoxemia was shown to correlate with the cytokine storm during COVID-19 (Arunachalam et al., 2020) and might cause activation-induced lymphocyte cell death. Assuming that the gut permeability could be altered during COVID-19-associated intestinal dysbiosis (Yeoh et al., 2021), we studied the circulating microbial populations associated with whole leukocytes by sequencing blood rDNA using next-generation sequencing of V3-V4 variable regions of the 16S rRNA bacterial gene as previously described (Païssé et al., 2016). Although we failed to observe significant quantitative differences in blood bacterial load between SVS (n=14) and LVS (n=15) patients, the linear discriminant analysis effect size indicated significant taxonomic differences in the bacteria family members between the two groups (Figure 6A-B). The DNA from Enterobacteriaceae (mainly composed of *Escherichia Shigella* genus) was overrepresented in leukocytes of LVS compared with SVS patients (Figure 6A-B, C left panel). The circulating Enterobacteriaceae-related DNA markedly anticorrelated with CCL22 (a hallmark of SVS, Figure 2A), but was strongly associated with the increase of exhausted CD8^+^ T lymphocytes (Figure 6D-E). There was a trend for an increase in the relative abundance of Micrococcaceae in the blood leukocytes of LVS that was confirmed in cancer patients with dismal prognosis (ALC < 800 & Ct < 25) (Figure 6F-G-H).

**Figure 6.**
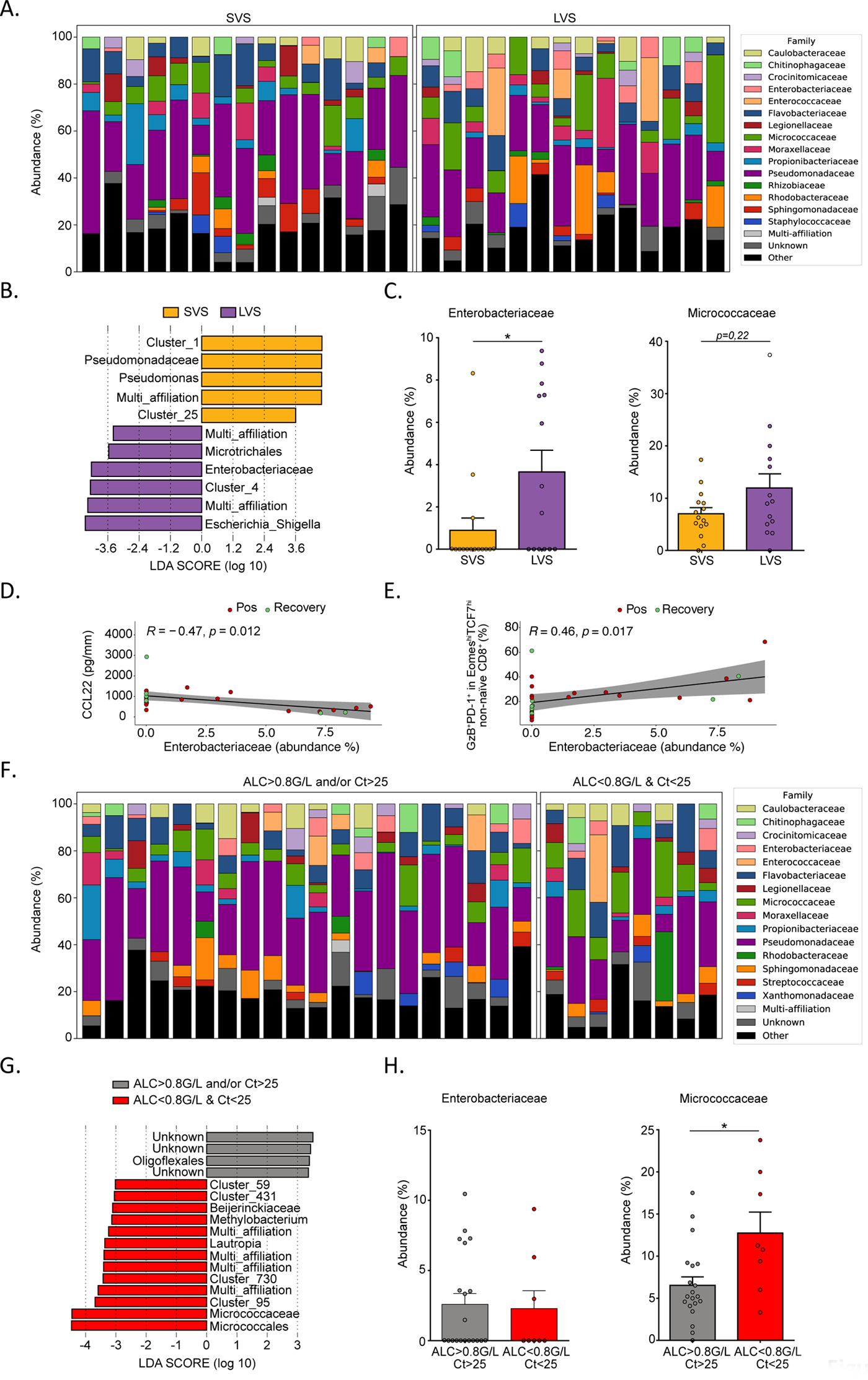
Lymphopenia and prolonged viral shedding are associated with blood recirculation of Enterobacteriaceae and Micrococcaceae DNA. **A.** Stacked bar charts showing the relative abundance of bacterial families obtained by 16S sequencing of the whole blood samples in patients experiencing Short term Viral RNA Shedding (SVS) and Long-term Viral RNA Shedding (LVS) among Cancer_FR1_TR. Only the top 15 most abundant bacterial families are represented (the others are in the category “Other”). **B.** Linear discriminant analysis effect size (LEfSe) analysis displaying linear discriminant analysis score (LDA) of the blood bacterial taxa differentially recovered from SVS (orange) versus LVS (purple) patients (\**p*<0.05 with Mann–Whitney test between the two groups of patients). **C.** Mean (bar plots, +/- SEM) and individual values (dot plots) of relative proportions of Enterobacteriaceae (**C**, **left panel**) and Micrococcaceae (**C**, **right panel**) family members in SARS-CoV-2 positive and Recovered patients. Significance between SVS and LVS patients was evaluated using Mann-Whitney test (\**p*<0.05). **D-E.** Spearman correlations between the relative proportions of Enterobacteriaceae with paired concentrations of CCL22 in serum (**D**) and with paired percentages of Granzyme B (GzB)^+^PD-1^+^ in Eomes^hi^TCF-1^hi^ non-naïve CD8^+^ measured in blood (**E**). **F.** Idem as in A. considering segregating the cohort in two groups; ALC > 0.8G/L and/or Ct > 25 patients versus ALC < 0.8G/L & Ct < 25 patients. **G.** LEfSe analysis displaying LDA score of the blood bacterial taxa significantly increased in ALC > 0.8G/L and/or Ct > 25 patients (grey) and ALC < 0.8G/L & Ct < 25 patients (red). The displayed bacterial taxa are significantly different (\**p*<0.05 with Mann–Whitney test) between the two groups of patients. **H.** Idem as in C segregating the cohort into the same two groups as in F. Significance between ALC > 0.8G/L and/or Ct > 25 patients and ALC < 0.8G/L & Ct < 25 patients was evaluated using the Mann-Whitney test (\**p*<0.05).

Overall, we conclude that virus-associated lymphopenia may result for complementary or coordinated orthogonal disorders.

## Discussion

To interrogate viral-host interactions during the COVID-19 pandemic in cancer patients, we studied 1106 patients, among them 59% were cancer bearers (FR1+FR2+FR3+CA), and 1063 COVID-19 positive (Figure 1A). We used high dimensional flow cytometry to perform deep immune profiling of innate, B and T cells and measurements of 51 soluble markers, with temporal analysis of immune changes during infection in one cohort that was further explored by blood metabolomics and metagenomics. This longitudinal immune analysis was linked to virologic and oncological data (Figure S5-S6). Using this approach, we made several intriguing observations.

First, 51%, 20% and 26% of cancer patients in FR1_TR, FR1_CR and CA, respectively, still shed SARS-CoV-2 RNA after day 40 from symptoms onset (*versus* 2% in HCW), correlating with high viral loads (Ct values < 25) at diagnosis. Indeed, isolation of replication-competent viral strains between 10 and 20 days after symptom onset has been documented in some persons with severe COVID-19, mostly in immunocompromised cases (Kampen et al., 2020). However, about 90% of their specimens no longer yielded replication-competent viruses after day 15 from symptom onset (Ge et al., 2020; Li et al., 2020). Prolonged shedding of influenza, parainfluenza, rhinovirus, seasonal coronavirus and respiratory syncytial virus has previously been detected in immunosuppressed patients (El Ramahi and Freifeld, 2019; Geis et al., 2013; Lehners et al., 2016, 2013; Milano et al., 2010). Cancer dissemination, cancer therapies and virus-induced lymphopenia might cause an immunodeficiency that eventually jeopardizes virus clearance. The proposed mechanisms by which lymphopenia occurs in COVID-19 (often shared with cancer dissemination) (Péron et al., 2013) include virus-induced atrophy of secondary lymphoid organs (Buja et al., 2020; Lax et al., 2020; Xu et al., 2020), disappearance of germinal centers (Kaneko et al., 2020), direct pro-apoptotic activity of the virus related to ACE2-dependent or ACE2-independent entry into lymphocytes (Pontelli et al., 2020), T cell demise consecutive to activation and exhaustion (Channappanavar and Perlman, 2017; Park, 2020), senescence (Derosa et al., 2020; Westmeier et al., 2020), and antiproliferative effects of lactic acid (Fischer et al., 2007). However, in our study, we found that lymphocyte loss was correlated with decrease of secondary biliary salts in LVS patients, most likely associated with increased gut permeability that lead to bacterial translocation, as we observed increased circulating DNA for Micrococcaceae and Enterobacteriaceae family members. Moreover, the transformation of spermidine into N1, N8 diacetylspermidine was linked to decreased ALC, in accordance with the role of spermidine in preventing ageing-related loss of lymphocyte fitness (Alsaleh et al., 2020; Puleston et al., 2014; Zhang et al., 2019).

Second, prolonged viral RNA carriage was associated with signs of immunopathology (exacerbated T cell responses, extrafollicular TFH, and plasmablast recirculation, exhausted PD-1^+^ Tc1 cells, sustained serum type 1 IFN levels), likely maintaining a positive feed-back loop for the expression of the interferon-signaling genes product ACE2 (Ziegler et al., 2020) and pro-inflammatory interactions between airway epithelia and immune cells (Chua et al., 2020).

Third, prolonged SARS-CoV-2 RNA shedding after day 40 might precede the aggravation of both COVID-19 and malignant disease. Indeed, virus and/or cancer-induced lymphopenia and T cell exhaustion may jointly enfeeble tumor immunosurveillance (Rd et al., 2011). Interestingly, SARS-CoV-2 virus-induced immunopathology was accompanied by increased blood levels of IL-8 (Figure S4G) and VEGF (Takahashi et al., 2020), which are well known pro-angiogenic and pro-tumorigenic growth factors, predicting failure to cancer immunotherapy (Sanmamed et al., 2017). Of note, patients with high initial viral loads or LVS tended to accumulate poor prognosis-related parameters than SVS or patients with higher Ct values in both cohorts (Table 1 & Table S1), being older (66 *versus* 56 years old, *p*=0.08), more metastatic at diagnosis of infection (72% *versus* 29%, *p*=0.011), and increased hospitalization rates (83% *versus* 23%, *p*=0.001). As a result, virus-induced lymphopenia markedly predicted early death of patients, within the first 2-3 months post-COVID-19 diagnosis in the first and second surge of the pandemic (in more than 200 patients) and call for caution to administer chemotherapy or steroids at the acute phase of the viral infection that exacerbate immunosuppression.

These observations call for a careful follow up of cancer patients, in particular those bearing hematological and metastatic malignancies, during the second wave of COVID-19. Given the non-consensual efficacy of vaccines against influenza virus in vulnerable individuals suffering from cancer-, virus- and age-associated lymphopenia (Bitterman et al., 2018; Péron et al., 2013), passive immunization of high affinity neutralizing monoclonal antibodies against SARS-CoV-2 at COVID-19 onset might be envisaged. This could be combined with therapeutic stimulation of lymphopoiesis (for instance with rIL-7, G-CSF, inhibitors of indole amine 2,3 deoxygenase), to achieve an immunological tonus that is compatible with anticancer treatments (Cheng et al., 2020; Francois et al., 2018; Laterre et al., 2020). Clinical trials are underway to evaluate rIL-7 against COVID-19, but may benefit from patients stratification based on Ct values, duration of viral RNA shedding and ALC (NCT04407689; NCT04426201; NCT04379076).

## Data Availability

Data and materials availability: This study includes no data deposited in external repositories.

## Acknowledgments

We are thankful to the Spectral flow cytometry facility team of Gustave Roussy. We thank the ET-EXTRA team (Biological Resource Center (NF 96-600) and the microbiology team for technical help. We thank Dr Aude Jary for helping to set up the subgenomic RNA analysis. We thank the staff from health and safety of Gustave Roussy Cancer Campus for helping to set up the translational research studies. We are thankful to the Genalyte for their supportive help.

## Contributions

L.D., L.Z., A.M. and F.B. conceived and designed the clinical trial. A-G.G., L.D. and L.Z. conceived and designed the translational research trial. A.G., F-X.D., S.T., C.M., L.A., B.B., A.St., B.G., M.Mer., F.S., A.M., L.D. included patients in the clinical trial. A-G.G., A.D., F-X.D., M.Pic., M.Maz., A.S., G.D. carried out all the experiments. A-G.G., A.D., F-X.D., A.S., and G.D. performed all the biological analysis. D.D. performed the non-supervised analysis and gave advices for statistical analyses. L.D. and C.A-C-S. collected and analyzed the clinical data. L.T. and A.F. provided the information from the HCW series. A.E., S.K. and B.R. analyzed and provided the clinical data from the Canadian series. B. L.S., D.R., S.G. and S.C. analyzed and provided the clinical data from the IHU series. G.G. and C.B. analyzed CT scan. E.C. and F.G. performed the RT-qPCR. A-G.G., C. A-C-S. and M.Maz. collected the biological data (ALC and Ct values). C.P. collected the samples. M.Maz., Y.H., E.P., C.F., G.F., C.T., I.L. and J-E. F. prepared the biological samples and provided some help for the experiments. A-G.G., A.D. L.Z, and L.D. interpreted data. performed the viral cultures. M.My. and G.G. performed the dosages of antibodies. F-X.D., C.G-D., S.D., N.N., F.A. and A-G.G performed the metagenomic experiments and analyses. B.L., A.R. and A.D. performed metagenomic analyses of blood microbiota. All the authors advised for the interpretation of the data. O.K. gave advices for the design of the figures. L.Z. and L.D. wrote the manuscript, with all authors contributing to writing and providing feedback.

## Funding

A-G.G. was supported by Fondation pour la Recherche Médicale (FRM). L.D. has received support by the Philanthropia Fondation Gustave Roussy. The Gustave Roussy sponsored clinical study on COVID-19 (ONCOVID; NCT NCT04341207 has been supported by the Fondation Gustave Roussy, the Dassault family, Malakoff Humanis, Agnès b., Izipizi, Ralph Lauren and Sanofi). L.Z. and G.K. were supported by RHU Torino Lumière (ANR-16-RHUS-0008), ONCOBIOME H2020 network, the Seerave Foundation, the Ligue contre le Cancer (équipe labelisée); Agence Nationale de la Recherche (ANR) – Projets blancs; ANR under the frame of E-Rare-2, the ERA-Net for Research on Rare Diseases; Association pour la recherche sur le cancer (ARC); Cancéropôle Ile-de-France; FRM; a donation by Elior; the European Research Council (ERC); Fondation Carrefour; High-end Foreign Expert Program in China (GDW20171100085 and GDW20181100051), Institut National du Cancer (INCa); Inserm (HTE); Institut Universitaire de France; LeDucq Foundation; the LabEx Immuno-Oncology; the SIRIC Stratified Oncology Cell DNA Repair and Tumor Immune Elimination (SOCRATE); CARE network (directed by Prof. Mariette, Kremlin Bicêtre AP-HP), and the SIRIC Cancer Research and Personalized Medicine (CARPEM). G.I. and M.P. were supported by Italian Ministry of Health (grants Ricerca CorrenteLinea 1, 1 ‘Infezioni Emergenti e Riemergenti’, projects COVID-2020-12371675 and COVID 2020 12371817). M.My and G.G. were supported by ANR Flash COVID19 program and ARS-CoV-2 Program of the Faculty of Medicine from Sorbonne University ICOViD programs (PI: G.G.).

### The authors declare the following competing interests

L.Z. and G.K. are cofounders of everImmune, a biotech company devoted to the use of commensal microbes for the treatment of cancers. A.G. and A.M. as part of the Drug Development Department (DITEP) are Principal/sub-Investigator of Clinical Trials for Abbvie, Adaptimmune, Aduro Biotech, Agios Pharmaceuticals, Amgen, Argen-X Bvba, Arno Therapeutics, Astex Pharmaceuticals, Astra Zeneca, Astra Zeneca Ab, Aveo, Bayer Healthcare Ag, Bbb Technologies Bv, Beigene, Bioalliance Pharma, Biontech Ag, Blueprint Medicines, Boehringer Ingelheim, Boston Pharmaceuticals, Bristol Myers Squibb, Bristol-Myers Squibb International Corporation, Ca, Celgene Corporation, Cephalon, Chugai Pharmaceutical Co., Clovis Oncology, Cullinan-Apollo, Daiichi Sankyo, Debiopharm S.A., Eisai, Eisai Limited, Eli Lilly, Exelixis, Forma Tharapeutics, Gamamabs, Genentech, Gilead Sciences, Glaxosmithkline, Glenmark Pharmaceuticals, H3 Biomedicine, Hoffmann La Roche Ag, Incyte Corporation, Innate Pharma, Institut De Recherche Pierre Fabre, Iris Servier, Janssen Cilag, Janssen Research Foundation, Kura Oncology, Kyowa Kirin Pharm. Dev., Lilly France, Loxo Oncology, Lytix Biopharma As, Medimmune, Menarini Ricerche, Merck Kgaa, Merck Sharp & Dohme Chibret, Merrimack Pharmaceuticals, Merus, Millennium Pharmaceuticals, Molecular Partners Ag, Nanobiotix, Nektar Therapeutics, Nerviano Medical Sciences, Novartis Pharma, Octimet Oncology Nv, Oncoethix, Oncomed, Oncopeptides, Onyx Therapeutics, Orion Pharma, Oryzon Genomics, Ose Pharma, Pfizer, Pharma Mar, Philogen S.P.A., Pierre Fabre Medicament, Plexxikon, Rigontec Gmbh, Roche, Sanofi Aventis, Sierra Oncology, Sotio A.S, Syros Pharmaceuticals, Taiho Pharma, Tesaro, Tioma Therapeutics, Wyeth Pharmaceuticals France, Xencor, Y’s Therapeutics, Research Grants from Astrazeneca, BMS, Boehringer Ingelheim, Janssen Cilag, Merck, Novartis, Pfizer, Roche, Sanofi. Non-financial support (drug supplied) from Astrazeneca, Bayer, BMS, Boringher Ingelheim, Johnson & Johnson, Lilly, Medimmune, Merck, NH TherAGuiX, Pfizer, Roche. N.L. reports to be a Speaker at Jazz Pharmaceutical E.D. reports grants and personal fees from ROCHE GENENTECH, grants from SERVIER, grants from ASTRAZENECA, grants and personal fees from MERCK SERONO, grants from BMS, grants from MSD, outside the submitted work. O.K. is a cofounder of Samsara Therapeutics. F.B. reports personal fees from Astra-Zeneca, Bayer, Bristol-Myers Squibb, Boehringer–Ingelheim, Eli Lilly Oncology,ß. Hoffmann–La Roche Ltd, Novartis, Merck, MSD, Pierre Fabre, Pfizer and Takeda, outside the submitted work. J-C. S. was a full time employee of AstraZeneca between September 2017 and December 2019, he reports consultancy: Relay Therapeutics, Gritstone Oncology and shares: Gritstone, AstraZeneca, Daiichi-Sankyo, outside the submitted work. L.A. reports consulting fees compensated to institution for Pfizer, Novartis, Bristol Myer Squibb, Ipsen, Roche, MSD, Astra Zeneca, Merck, Amgen, Astellas, Exelixis, Corvus Pharmaceuticals, Peloton Therapeutics, outside the submitted work. F.S. reports consulting fees from AMGEN, Roche, Chugai, Mylan, Mundi Pharma, Leo Pharma, Pierre Fabre Oncology, Helsinn, MSD, Pfizer, BMS, outside the submitted work.

## Data and materials availability

This study includes no data deposited in external repositories.

## Table

**Table 1.** Clinical characteristics of Cancer_FR1 and Cancer_FR3 patients from discovery and validation cohorts presenting cycle threshold below (Ct<25) or above 25 (Ct>25) and with (< 800/mm^3^) or without (> 800/mm^3^) lymphopenia at diagnosis (refer to Figure 3D-F). Abbreviations: BMI: Body Mass Index; COPD: Chronic Obstructive Pulmonary Disease; CR: Clinical Routine; Ct: Cycle threshold; DM: Diabetes mellitus; H: Hematological malignancies; ICU: Intensive Care Unit; n: number; NED: no evidence of disease; no: number; PD: Progressive disease; PS: Performance Status; S: Solid tumors; SD/PR: Stable disease/Partial response; TR: Translational Research; *in the 4 weeks before inclusion. Statistical analyses: ANOVA (Kruskal-Wallis)(#), Chi-Square or Fisher’s exact tests. °unknown for Cancer_FR3_discovery (n=26 patients), calculations with Cancer_FR1_discovery, n=84. °*unknown for Cancer_FR3_validation (n=25 patients), calculations with Cancer_FR1_validation, n=91.

## Materials and methods

All cohorts (refer to **Supplementary Material,** Table 1)

**Supplementary Material, Table 1.**
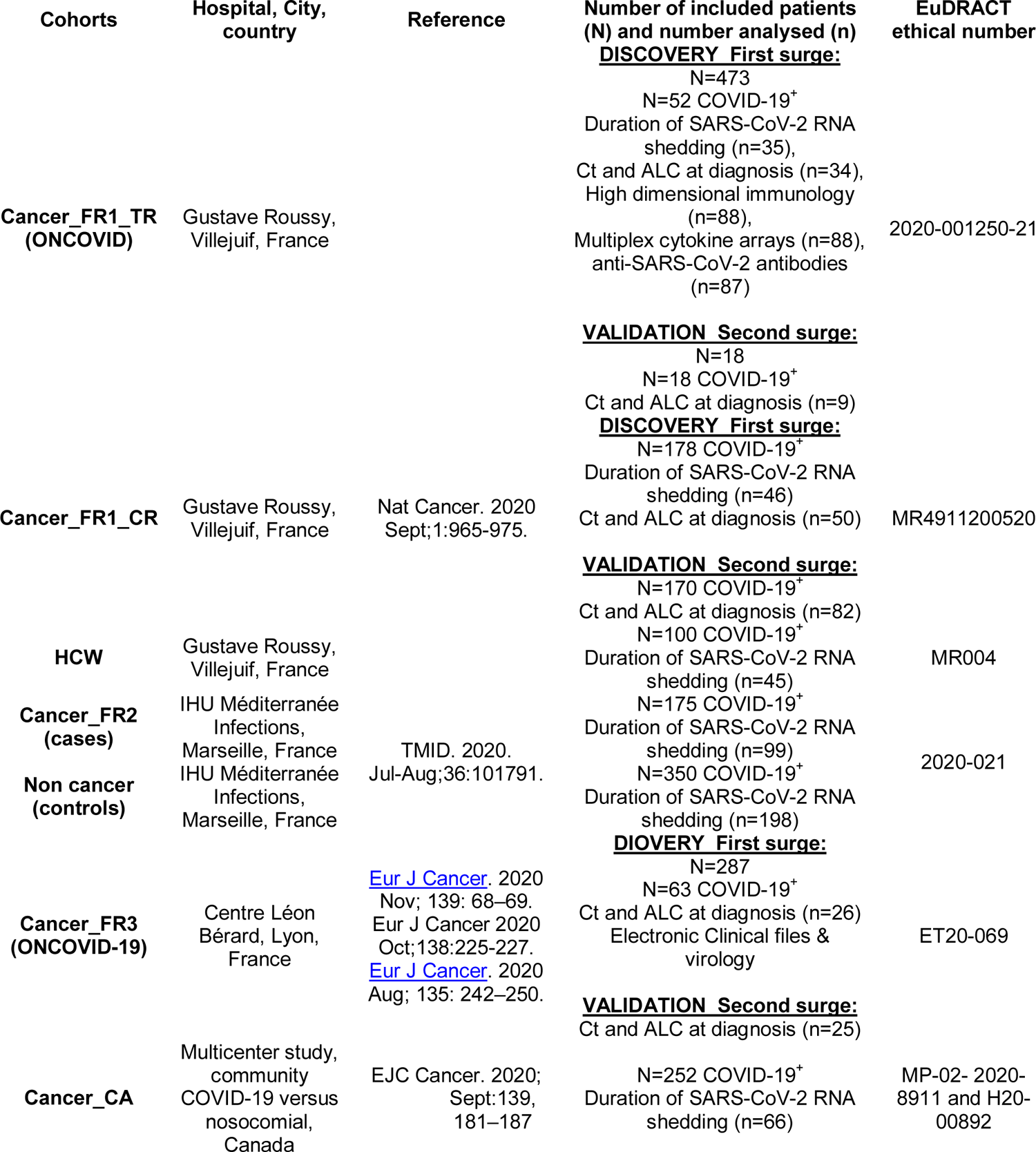
List of all cohorts analysed in the study.

### Cohorts for the duration of viral PCR positivity

#### 1/ Cancer_FR1_Translational Research (TR) (ONCOVID) clinical trial and regulatory approvals for translational research. Principles

Gustave Roussy Cancer Center sponsored the “ONCOVID” trial and collaborated with the academic authors on the design of the trial and on the collection, analysis, and interpretation of the data. Sanofi provided trial drugs. The trial was conducted in accordance with Good Clinical Practice guidelines and the provisions of the Declaration of Helsinki. All patients provided written informed consent. Protocol approval was obtained from an independent ethics committee (ethics protocol number EudraCT No: 2020-001250-21). The protocol is available with the full text of this article at https://clinicaltrials.gov/ct2/show/NCT04341207.

#### Patients

ONCOVID eligible patients were adults fitted for, or under, or recently treated by chemotherapy and/or immune-checkpoint blockade for the treatment of solid tumors or hematological malignancies (Refer to Table 1 and Table S1). Patients diagnosed for COVID-19 from April 10^th^, 2020 to May 4^th^, 2020 were included in the Discovery cohort and patients from May 5^th^, 2020 to November 25^th^, 2020 were included in the Validation cohort.

#### Trial design

Cancer patients were screened for SARS-CoV-2 virus carriage by nasopharyngeal sampling at every hospital visit. The presence of SARS-CoV-2 RNA was detected by RT-qPCR assay in a BSL-2 laboratory. Asymptomatic and symptomatic patients (*i.e* presenting with fever (t°>38°C) and/or cough and/or shortness of breath and/or headache and/or fatigue and/or runny nose and/or sore throat, anosmy/agueusia) with a positive SARS-CoV-2 RT-qPCR test, shifted to the interventional phase (tailored experimental approach with Hydroxychloroquine and Azithromycin therapy in symptomatic SARS-CoV-2 positive subjects). Asymptomatic or symptomatic patients with negative SARS-CoV-2 RT-qPCR test continued their standard of care anti-cancer treatments. Repeated RT-qPCR for SARS-CoV-2 on nasopharyngeal swabs and blood samples were performed to monitor the status for SARS-CoV-2 and the immune response, respectively, in COVID-19 positive and negative patients. The COVID-19 severity was defined based on oxygen, imaging and hospitalization criteria. Patients with a mild COVID-19 disease had limited clinical symptoms not requiring scan or hospitalization; patients with a moderate COVID-19 disease were symptomatic with a dyspnea and radiological findings of pneumonia on thoracic scan requiring hospitalization and a maximum of 9L/min of oxygen; severe patients had respiratory distress requiring intensive care and/or more than 9L/min of oxygen.

#### Samples for translational research

Whole blood was used for high dimensional spectral flow cytometry analyses. Serum samples were used to monitor the concentrations of cytokines and chemokines release and to titer anti-SARS-CoV-2 IgG, M and A antibodies (see blood analysis section) (Supplementary Material Figure 1).

#### 2/ Health Care Workers (HCW) of Cancer_FR1_TR

The part of the research including health care workers was conducted in compliance with General Data Protection Regulation (GDPR) and the French Data Protection Authority’s recommendation about Data Protection in clinical researches. Gustave Roussy Data Protection Officer (DPO) has evaluated this project and sent to the principal investigator a formalized-operational action plan about data protection compliance: patient’s information, security measures, good practices about pseudonymization, etc. All of the DPO’s recommendations has been applied by the research team. Health care workers diagnosed for COVID-19 between 24^th^ March and 24^th^ April 2020 were included. Results of RT-qPCR, cycle threshold, age, gender, and number of comorbidities were collected. Data from health care workers who refused to participate and/or with a cancer were excluded. In agreement with MR004 in France, we reported the series to the national information science and liberties commission.

#### 3/ Second series of patients with cancer (Cancer_FR2)

*CASE-CONTROL study.* All comers spontaneously presenting at a general hospital for infectious diseases (IHU Méditerranée Infections, Marseille, FR) (Table S1) from February 27^th^, 2020 to December 15^th^, 2020 composed of 996 COVID-19 patients. We performed a case-control study at a 1:2 paired ratio where the 175 cancer patients (with a currently treated cancer or history of cancer) were matched with 350 cancer free individuals on age, gender, comorbidities relevant for COVID-19. Of note, >75% received hydroxychloroquine and >96% received azithromycine (Table S1) (Amrane et al., 2020; Lagier et al., 2020). This study was approved by the IHU Méditerranée Infections review board committee (Méditerranée Infection N°: 2020-021).

#### 4/ Third series of cancer patients from Canada (Cancer_CA)

We used 66 individuals from the clinical cohort previously reported(Elkrief et al., 2020) for whom data were available (Table S1). This study was conducted across eight Canadian institutions in Quebec and British Columbia and was approved by the institutional ethics committee at each site (Ethics number: MP-02-2020-8911 and H20-00892).

#### 5/Fifth series of cancer patients, Cancer_FR1_Clinical Routine (CR)

We used the clinical cohort previously reported(Albiges et al., 2020) (Table S1). In accordance with the French regulations, there was no requirement for ethical approval to be sought for this observational study, based on medical files. Patients diagnosed for COVID-19 from March 14^th^, 2020 to April 29^th^, 2020 were included in the Discovery cohort and from April 29^th^, 20 to November 25^th^, 2020 in the Validation cohort. This study was also declared to the Gustave Roussy Cancer Centre’s data protection officer and registered on the website of the French Healthcare Data Institute (declaration number: MR4911200520).

### Cohorts for the ALC and Ct value predictors: first surge and second surge of the pandemic

#### 1/ Cancer_FR1_Translational Research (TR) (ONCOVID) clinical trial and regulatory approvals for translational research

Among the 52 patients diagnosed for COVID-19 during the first surge (from April 10^th^, 2020 to May 4^th^, 2020), absolute lymphocyte count (ALC) and cycle threshold (Ct) were available for 34 patients whom were included in this cohort. Then, among the 18 patients included in ONCOVID during the second surge (from May 5^th^, 2020 to November 25^th^, 2020), absolute lymphocyte count (ALC) and cycle threshold (Ct) were available for 9 patients whom were included in this cohort

#### 2/ Cancer patients referred to the clinical routine (Cancer_FR1_CR)

In accordance with the French regulations, there was no requirement for ethical approval to be sought for this observational study, based on medical files. Among the 178 patients diagnosed for COVID-19 during the first surge (March 14^th^, 2020 to April 29^th^, 2020), ALC and Ct were available for 50 patients whom were included in this cohort. Then, among 170 patients with cancer diagnosed for COVID-19 during the second surge (from May 5^th^, 2020 to November 25^th^, 2020), ALC and cycle threshold Ct were available for 82 patients whom were included in this cohort.

#### 3/ Cancer patients referred to the Centre Léon Bérard, Lyon, France (Cancer_FR3)

The PRE-ONCOVID-19 study was approved by the Institutional review board of the Centre Leon Bérard on March 12^th^, 2020 (ET20-069). We used a subset of 25 patients included during the first surge from March 5^th^, 2020 to May 4^th^, 2020 with available ALC and Ct values. We used 26 patients included during the second surge from October 1^th^, 2020 to December 5^th^, 2020 with available data. Patients from each cohort were classified using the same criteria.

### RT-qPCR analysis

SARS-CoV-2 diagnostic testing of clinical nasopharyngeal swabs or other samples by RT-qPCR was conducted from March 14^th^, 2020 to March 23^th^, 2020 at an outside facility using the Charité protocol. From March 23^th^, 2020 testing was performed internally at the Gustave Roussy. The cycle thresholds were collected only for assays performed at Gustave Roussy. Nasopharyngeal swab samples were collected using flocked swabs (Sigma Virocult) and placed in viral transport media. SARS-CoV-2 RNA was detected using one of two available technics at Gustave Roussy: the GeneFinder COVID-19 Plus Real*Amp* kit (ELITech Group) targeting three regions (*RdRp* gene, nucleocapsid and envelope genes) on the ELITe InGenius (ELITech Group) or the multiplex real-time RT-PCR diagnostic kit (the Applied Biosystems TaqPath COVID-19 CE-IVD RT-PCR Kit) targeting three regions (*ORF1ab*, nucleocapsid and spike genes) with the following modifications. Nucleic acids were extracted from specimens using automated Maxwell instruments following the manufacturer’s instructions (Maxwell RSC simplyRNA Blood Kit; AS1380; Promega). Real-time RT-PCR was performed on the QuantiStudio 5 Dx Real-Time PCR System (Thermo Fisher Scientific) in a final reaction volume of 20[μ including 5 l of extracted nucleic acids according to the manufacturer instruction.

The cut-off value of 25 for the cycle threshold was based on the median calculated on Cancer_FR1_TR and the mean calculated on Cancer_FR1_TR+CR.

### RT-PCR for subgenomic RNA (sgRNA) for SARS-CoV-2

We used the protocol previously described by Wölfel et al. in *Nature*, 2020. Briefly, the oligonucleotide sequence of the leader specific primer was as follows: sgLeadSARSCoV2-F; CGATCTCTTGTAGATCTGTTCTC, and the oligonucleotide sequence of the E primer was as follows: E_Sarbeco_R; ATATTGCAGCAGTACGCACACA. Briefly, 5 uL of RNA (>21 ng) were used for the sgRNA RT-PCR assay with Superscript III one-step RT-PCR system with Platinum Taq Polymerase (Invitrogen, Darmstadt, Germany) with 400 nM concentration of each primer. Thermal cycling was set up as described. Finally, RT-PCR products for sgRNAs were analyzed on agarose gel 2%.

### Evaluation of SARS-CoV-2 RNA shedding

The duration of viral shedding was defined as the number of days from the first positive to the first negative RT-qPCR, after longitudinal monitoring. In order to prevent an overvaluation of this duration, we considered in this analysis only patients with an interval below 40 days between the last positive RT-qPCR and the first negative RT-qPCR. Six patients had one negative RT-qPCR followed by positive RT-qPCR. We extend the duration to the second negative RT-qPCR for 3 patients with a cycle threshold below 35 for the gene coding replication-transcription complex and within 6 days after the first negative result.

### Absolute Lymphocyte Count (ALC)

The absolute lymphocyte count was measured for the clinical routine using the Sysmex XN (Sysmex, Belgium). Values “PRE” were collected between 210 and 12 days before the symptom onset of COVID-19, values at diagnosis of the infection were collected between −4 and +7 days of the disease diagnosis by RT-qPCR, values “POST” were collected at the recovery time or later, meaning between 0 and 123 days after the first negative RT-qPCR. For the interpretation, the cut-off value for ALC was the median found in patients with high viral load at diagnosis (ALC=800/m^3^). In parallel, we considered this value as relevant according to the common terminology criteria for adverse events where grades of lymphopenia were assigned as following: grade 1 ALC < lower limit of normal to 800/mm3, grade 2 ALC < 800-500/mm3, and grade 3 ALC < 500-200/mm3.

### Blood tests. Sampling

Blood samples were drawn from patients enrolled in ONCOVID at Gustave Roussy Cancer Campus (Villejuif, France). Whole human peripheral blood was collected into sterile vacutainer tubes.

### Spectral flow cytometry

One hundred and twenty-one whole blood samples from 88 patients (Supplementary Material Figure 1) was mixed at a 1:1 ratio with Whole Blood Cell Stabilizer (Cytodelics), incubated at room temperature for 10 min and transferred to −80°C freezer to await analysis. These samples were secondarily thawed in a water bath set to +37°C. Cells were fixed at a ratio 1:1 with Fixation Buffer (Cytodelics, ratio 1:1) and incubated for 10 min at room temperature. Red blood cells were lysed by addition of 2 mL of Lysis Buffer (Cytodelics, ratio 1:4) at room temperature for 10 min. White blood cells were washed with 2 mL of Wash Buffer (Cytodelics, ratio 1:5). Cells were resuspended in 100 µL extra-cellular antibody cocktail and incubated at room temperature for 15 min, then washed in Flow Cytometry Buffer (PBS containing 2% of fetal bovine serum and 2 mM EDTA). For intra-cellular labelling, a step of permeabilization was performed using 200 µL of eBioscience Foxp3 kit (ThermoFischer); cells were then incubated for 40 min at +4°C, washed in Perm Buffer (ThermoFischer) and resuspended in intra-cellular antibody cocktail. After incubation, cells were washed in Flow Cytometry Buffer and resuspended to proceed to the acquisition. All antibodies used are listed in Supplemental Material Table 2. Samples were acquired on CyTEK Aurora flow cytometer (Cytek Biosciences).

### 16S rDNA metagenomic profiling

DNA from blood was isolated and amplified in a strictly controlled environment at Vaiomer SAS (Labège, France) using a stringent contamination-aware approach as discussed previously (Anhê et al., 2020; Lluch et al., 2015; Païssé et al., 2016; Schierwagen et al., 2020). The microbial populations based on rDNA present in blood were determined using next-generation sequencing of V3-V4 variable regions of the 16S rRNA bacterial gene as previously described (Lluch et al., 2015). For each sample, a sequencing library was generated by addition of sequencing adapters. The joint pair length was set to encompass a 467 base pairs amplicon (using Escherichia coli 16S as a reference) with a 2 × 300 paired end MiSeq kit V3 (Illumina, San Diego, CA, USA). The detection of the sequencing fragments was performed using the MiSeq Illumina® technology. Targeted metagenomic sequences from microbiota were analysed using the bioinformatic pipeline from the FROGS guideline (Escudié et al., 2018). Briefly, the cleaning was done by removing amplicons without the two PCR primers (10% of mismatches were authorised), amplicons with at least one ambiguous nucleotide (‘N’), amplicons identified as chimera (with vsearch v1.9.5), and amplicons with a strong similarity (coverage and identity ≥ 80%) with the phiX (library used as a control for Illumina sequencing runs). Clustering was produced in two passes of the swarm algorithm v2.1.6. The first pass was a clustering with an aggregation distance equal to 1. The second pass was a clustering with an aggregation distance equal to 3. Taxonomic assignment of amplicons into operational taxonomic units (OTUs) was produced by Blast+ v2.2.30+ with the Silva 134 Parc databank. To assess if the richness of microbiota was adequately captured by metagenomic sequencing, a rarefaction analysis was performed. To ensure a low background signal from bacterial contamination of reagents and consumables, two types of negative controls consisting of molecular grade water were added in an empty tube separately at the DNA extraction step and at the PCR steps and amplified and sequenced at the same time as the extracted DNA of the blood samples. The controls confirm that bacterial contamination was well contained in our pipeline and had a negligible impact on the taxonomic profiles of the samples of this study as published before (Anhê et al., 2020; Lluch et al., 2015; Païssé et al., 2016; Schierwagen et al., 2020). One sample has been excluded of the analyses for aberrant profile.

### Serum tests

Serums from 120 samples corresponding to 88 patients (Supplementary Material Figure 1) were collected from whole blood after centrifugation at 600 g for 10 min at room temperature and transferred to −80°C freezer to await analysis.

### Multiplex cytokine and chemokine measurements

Serum samples were centrifuged for 15 min at 1,000 g, diluted 1:4, then monitored using the Bio-Plex ProTM Human Chemokine Panel 40-plex Assay (Bio-rad, ref: 171AK99MR2) according to the manufacturer’s instructions. 40-plex cytokines and chemokines provided are: CCL1, CCL11, CCL13, CCL15, CCL17, CCL19, CCL2, CCL20, CCL21, CCL22, CCL23, CCL24, CCL25, CCL26, CCL27, CCL3, CCL7, CCL8, CX3CL1, CXCL1, CXCL10, CXCL11, CXCL12, CXCL13, CXCL16, CXCL2, CXCL5, CXCL6, CXCL8, CXCL9, GM-CSF, IFN-γ, IL-10, IL-16, IL-1, IL-2, IL-4, IL-6, MIF, TNF-α. Acquisitions and analyses were performed on a Bio-Plex 200 system (Bio-rad) and a Bio-Plex Manager 6.1 Software (Bio-rad), respectively. Soluble Calprotectin (diluted 1:100) and IFN-α2a were analyzed using a R-plex Human Calprotectin Antibody Set (Meso Scale Discovery, ref: F21YB-3) and the ultra-sensitive assay S-plex Human IFN-α2a kit (Meso Scale Discovery, ref: K151P3S-1), respectively, following manufacturer’s instructions. Acquisitions and analyses of soluble Calprotectin and IFN-α2a were performed on a MESO™ QuickPlex SQ120 reader and the MSD’s Discovery Workbench 4.0. Each serum sample was assayed twice with the average value taken as the final result.

### Serology: Anti-SARS-CoV-2 immunoglobulins

Serum was collected from whole blood after centrifugation at 600 g for 10 min at room temperature and transferred to −80°C freezer to await analysis. Serological analysis SARS-CoV-2 specific IgA, IgM and IgG antibodies were measured in 119 serum samples from 87 patients (Supplementary Material Figure 1) with The Maverick ™ SARS-CoV-2 Multi-Antigen Serology Panel (Genalyte Inc. USA) according to the manufacturer’s instructions. The Maverick ™ SARS-CoV-2 Multi-Antigen Serology Panel (Genalyte Inc) is designed to detect antibodies to five SARS-CoV-2 antigens: nucleocapsid, Spike S1 RBD, Spike S1S2, Spike S2 and Spike S1 with in a multiplex format based on photonic ring resonance technology(Sterlin et al., 2020). This system detects and measure with good reproducibility changes in resonance when antibodies bind to their respective antigens in the chip. The instrument automates the assay. Briefly, 10µl of each serum samples were added in a sample well plate array containing required diluents and buffers. The plate and chip are loaded in the instrument. First the chip is equilibrated with the diluent buffer to get baseline resonance. Serum sample is then charged over the chip to bind specific antibodies to antigens present on the chip. Next, chip is washed to remove low affinity binders. Finally, specific antibodies of patients are detected with anti-IgG or -IgA or - IgM secondary antibodies.

### Metabolomics analysis

Samples were prepared as previously described (Danlos et al., In Press). Briefly, serum samples were mixed with ice-cold extraction mixture (methanol/water, 9/1, v/v, with a mixture of internal standards), then centrifugated. Supernatants were collected for widely-targeted analysis of intracellular metabolites. ***GC/MS analysis***. GC-MS/MS method was performed on a 7890B gas chromatography (Agilent Technologies, Waldbronn, Germany) coupled to a triple quadrupole 7000C (Agilent Technologies, Waldbronn, Germany) equipped with a high sensitivity electronic impact source (EI) operating in positive mode. ***Targeted analysis of bile acids***. Targeted analysis was performed on a RRLC 1260 system (Agilent Technologies, Waldbronn, Germany) coupled to a QTRAP 6500+ (Sciex) equipped with an electrospray source operating in negative mode. Gas temperature was set to 450°C, with ion source gas 1 and 2 set to 30 and 70, respectively. ***Targeted analysis of polyamines***. Targeted analysis was performed on a RRLC 1260 system (Agilent Technologies, Waldbronn, Germany) coupled to a QQQ 6410 (Agilent Technologies) equipped with an electrospray source operating in positive mode. The gas temperature was set to 350°C with a gas flow of 12 l/min. The capillary voltage was set to 3.5 kV. ***Targeted analysis of SCFA***. Targeted analysis was performed on a RRLC 1260 system (Agilent Technologies, Waldbronn, Germany) coupled to a QQQ 6410 (Agilent Technologies) equipped with an electrospray source operating in negative mode. Gas temperature was set to 350°C with a gas flow of 12 L/min. Capillary voltage was set to 4.0 kV. ***Pseudo-targeted analysis of intracellular metabolites***. The profiling experiment was performed with a Dionex Ultimate 3000 UHPLC system (Thermo Scientific) coupled to a Q-Exactive (Thermo Scientific) equipped with an electrospray source operating in both positive and negative mode and full scan mode from 100 to 1200 m/z. The Q-Exactive parameters were: sheath gas flow rate 55 au, auxiliary gas flow rate 15 au, spray voltage 3.3 kV, capillary temperature 300°C, S-Lens RF level 55 V. The mass spectrometer was calibrated with sodium acetate solution dedicated to low mass calibration.

### Data analysis. Spectral flow cytometry

Fcs files were exported and analyzed using FlowJo software using the gating strategy showed in Supplementary material, Figure 2. Briefly, gates on CD45^+^, CD3^+^ or CD19^+^ from the myeloid, T cell and B panels, respectively, were exported in an fcs file. All exported gate**s** from one panel were used to generate an UMAP(Becht et al., 2019). As shown on Supplementary material Figures 3 and 4, we used relative expression and manual gating strategy. For patients treated by anti-PD-1 monoclonal antibody, the gates including PD-1 were excluded of the analysis. For patients treated by anti-CD38 monoclonal antibody, the gates including CD38 were excluded of the analysis. ***Representation of the results.*** Data representation was performed with software R v3.3.3 using tidyverse, dplyr, ggplot2, ggpubr, pheatmap, corrplot or Hmisc packages or GraphPad Prism 7.

### Statistical analyses

Calculations and statistical tests were performed either with R v3.3.3 or Prism 7 (GraphPad, San Diego, CA, USA). Unless stated, p-values are two-sided with 95% confidence intervals for the reported statistic of interest. Individual data points representing the measurement from one patient are systematically calculated from the corresponding distribution. Biological parameters associated to statistically significant differences between groups were considered for the data visualization described below. Group comparison was performed using one-way ANOVA with the lmer function of the lme4 R package. The p-values were computed with the Kenward-Roger method, available in the lmertest R package. Spearman correlations were computed using Hmisc and Pheatmap R package. Hierarchical clustering of the patient’s factors was performed using using the hclust R package. The redundancy analysis (RDA) was performed using the vegan R package to explore the association between the clinical variables and the biological parameters correlation latent structure. The RDA performs variance decomposition such as principal component analysis, but including additional supervised components depending on the explanatory variables (e.g. clinical factors). The association of the clinical factors with the biological parameter correlation latent structure was tested using permutation test. Kaplan–Meier methodology was used to estimate the probability of overall survival as well as to visualize the median time of SARS-CoV-2 RNA shedding for each group (HCW and Cancer). One-way ANOVA (paired and unpaired) with Kenward-Roger method was used to calculate p-value between ALC among groups of viral RNA shedding and Ct values. Chi-Square, Fischer test were used to calculate the differences in proportion between groups. Comparing two groups, Mann-Whitney test was used. Univariate analyses were performed with the Cox regression model. *p*<0.05 was considered as significant. Multivariate Cox analysis was performed using the survival R package stratified for the cohort and adjusted for the age, ECOG performance status, gender and metastatic status and hematological malignancy.

## Supplemental figure legends

**Figure S1. Cancer_FR1_Translationnal Research patient clinical characteristics**. **A.** Consort flow chart diagram of ONCOVID (Cancer_FR1_Translationnal Research (TR)) patients at trial inclusion, delineating COVID-19 positive cases retained for the evaluation of duration of SARS-CoV-2 RNA shedding and cycle threshold (Ct) with absolute lymphocyte count (ALC) at diagnosis, and their controls for the translational research ancillary study (Cancer_FR1). **B-G.** Number (percentages) of patients with cancer diagnosed with COVID-19 and with hematological (H) *versus* solid (S) malignancy (**B-C**), presenting symptomatic (Sym) *vs* asymptomatic (Asym) infection (**D-E**), and at different stages of disease (localized (L), locally advanced (LA), metastatic (M)) of their cancer (**F-G**).

**Figure S2.** Reliability of the monitoring of viral shedding. Consort flow chart diagram of patients with cancer (**A**) and healthcare workers (HCW) (**B)** diagnosed with COVID-19 retained for the evaluation of duration of SARS-CoV-2 RNA shedding in the ancillary study presented in Figure 1.

Figure S3. Patient clinical characteristics in a second series of patients with cancer at Gustave Roussy, Cancer_FR1_Clinical Routine. **A.** Consort flow chart diagram of second series patients with cancer (Cancer_FR1_Clinical Routine (CR)) diagnosed with COVID-19 at Gustave Roussy and monitored in clinical routine during the pandemic (March 14^th^ - April 29^th^) retained for the evaluation of duration of SARS-CoV-2 RNA shedding and cycle threshold (Ct) with absolute lymphocyte count (ALC) at diagnosis. **B-G.** Proportions of COVID-19 cancer patients with hematological (H) *vs* solid malignancy (S) (**B-C**), presenting symptomatic (Sym) *vs* asymptomatic (Asym) infection (**D-E**), and at different stages of disease (localized (L), locally advanced (LA), metastatic (M)) of their cancer (**F-G**).

Figure S4. Immunotypes associated with prolonged viral RNA shedding in patients with cancer. Temporal changes and correlation of blood leukocyte parameters measured by high dimensional spectral flow cytometry and multiple soluble factors in various phases of COVID-19 presentation (no virus infection (Ctls, grey dots), asymptomatic viral infection (Asym, light blue dots), symptomatic viral infection examined in the first 20 days (≤20d) or after 20 days (>20d) of symptoms with those experiencing short term viral RNA shedding (SVS, orange dots) or long term viral RNA shedding (LVS, purple dots) and RT-qPCR negative COVID-19 patients in the convalescent phase (Recovery, green dots)). Box plots display a group of numerical data through their 3^rd^ and 1^st^ quartiles (boxe), mean (central band), minimum and maximum (whiskers). Each dot represents one sample, each patient being drawn 1-3 times. Statistical analyses used one-way ANOVA with Kenward-Roger method to take into account the number of specimen/patient: \**p*<0.05, \*\**p*<0.01, \*\*\**p*<0.001, \*\*\*\**p*<0.0001. **A.** Monocyte subset (**A, left panel**) defined within the CD45^+^CD3^-^CD56^-^ CD19^-^CD15^-^ gate, focusing on CD169^-^HLA-DR^+^ in CD16^low^CD14^+^ classical monocytes (cMo) (**A, middle panel**) and CD16^+^CD14^low^ non-classical monocytes (non-cMo) (**A, right panel**). **B.** Neutrophil subset (**B, left panel**) defined within the CD45^+^CD15^+^ gate, focusing on CD101, CD10 simple positive or double negative subset (**B, right panel**). **C.** Transitional B cell subset (CD24^+^CD38^hi^) in B cells. **D.** Serum titers of anti-S1 RBD IgA (**D, left panel**) and IgM (**D, right panel**) in various patient groups at different kinetics. **E-I.** Temporal changes of serum concentrations of various soluble factors and their ratios measured in various phases of COVID-19 presentation.

Figure S5. Non-supervised hierarchical clustering of 33 cancer patients negative for COVID-19 and 21 patients with cancer diagnosed positive for COVID-19 investigated for immunophenotyping and viral parameters within the first 30 days from symptoms onset (or positive RT-qPCR for asymptomatic patients) or COVID-19 negative controls. The heatmap shows z score-normalized concentration of parameters. Each column represents a patient and each row a parameter. The color gradient from blue up to red indicates increasing gradients of concentrations.

Figure S6. Non supervised hierarchical clustering of 29 cancer patients negative for COVID-19 and 35 patients with cancer recovered from COVID-19 investigated for immunophenotyping and viral parameters. The heatmap shows z score-normalized concentration of parameters. Each column represents a patient and each row a parameter. The color gradient from blue up to red indicates increasing gradients of concentrations. In order to assess the immune characteristics after COVID-19 events in convalescent *vs* controls, we compared only alive patients and we excluded all the dead ones in this analysis.

Figure S7. Lymphoid parameters associated with prolonged viral RNA shedding in patients with cancer. Temporal changes and correlation of blood leukocyte parameters measured by high dimensional spectral flow cytometry in various phases of COVID-19 presentation (no virus infection (Ctls, grey dots), asymptomatic viral infection (Asym, light blue dots), symptomatic viral infection examined in the first 20 days (≤20d) or after 20 days (>20d) of symptoms with those experiencing short term viral RNA shedding (SVS, orange dots) or long term viral RNA shedding (LVS, purple dots) and RT-qPCR negative COVID-19 patients in the convalescent phase (Recovery, green dots)). Box plots display a group of numerical data through their 3^rd^ and 1^st^ quartiles (box), mean (central band), minimum and maximum (whiskers). Each dot represents one sample, each patient being drawn 1-3 times. Statistical analyses used one-way ANOVA with Kenward-Roger method to take into account the number of specimen/patient: \**p*<0.05, \*\**p*<0.01, \*\*\**p*<0.001, \*\*\*\**p*<0.0001. **A.** Lymphocyte relative proportion within the CD45^+^ gate, encompassing CD3^+^, CD56^+^ and CD19^+^ cell subsets (**A, left panel**) with (**A, middle**) corresponding to CD19^+^ B cells and (**A, right**) corresponding to NK cells such as CD45^+^CD3^-^CD56^+^CD19^-^. **B.** CD8^+^ T cells (**B, left panel**) or CD4^+^ T cells (**B, right panel**) within the CD45^+^CD3^+^gate. **C.** Percentages of EM2 defined as CD45RA^-^CD27^-^CD62L^+^ in CD8^+^ T cells (**C, left panel**) and subset co-expressing HLA-DR and CD38 in non-naïve CD8^+^ (C, right panel).

Figure S8. Impact of ALC and Ct values in cancer patients survival according to their cancer type (hematological *versus* solid). Kaplan Meier curve and Cox regression analysis of cancer-related overall survival in patients pooled in the two institutes (Gustave Roussy and Centre Léon Bérard: Cancer_FR1_TR, Cancer_FR1_CR and Cancer_FR3) and in the two surges of the pandemic to analyze the impact of the Absolute Lymphocyte Count (ALC) and Cycle threshold (Ct) at diagnosis in solid tumors only (**A**) (n=177 patients) and hematological tumors only (**B**) (n=49 patients).

Figure S9. Serum metabolome variations in COVID-19 positive cancer patients, according to viral shedding duration, cycle threshold and absolute lymphocyte counts at diagnosis. Targeted metabolomics analysis was performed on serum samples from confirmed COVID-19 patients, allowing the identification of significant changes in the relative abundance of 221 metabolites, illustrated by a heatmap. Hierarchical clustering (Euclidean distance, ward linkage method) of the metabolite abundance is shown, with each row representing a metabolite and each column corresponding to a different patient. Polyamine derived metabolites, and bile acids and their conjugates are marked in red.

Figure S10. Lymphopenia and viral shedding are associated with perturbations in polyamine and secondary biliary acid pathways. **A-B. Biliary salt pathway related-metabolites.** Mass-spectrometry based abundance of serum Hyo-(**A**) and Urso-(**B**) deoxycholic acid in LVS (purple dots), SVS (orange dots) (**left panel**) compared to controls (Ctls) and Spearman correlations with ALC (**right panel**), color code corresponds to status for COVID-19 and the category of viral shedding. **C-D. Polyamine pathway-related metabolites.** N1-acetylspermidine relative abundance significantly increased in long term viral RNA shedding (LVS, purple dots) regarding controls (Ctls) (**C**, **left panel**) and anticorrelated with ALC (**C**, **right panel**), color code corresponds to the category of Ct and ALC at diagnosis. N1, N12-diacetylspermine relative abundance significantly increased in critical non-cancer patients (**D**, **left panel**) and anticorrelated with Absolute Lymphocyte Count (ALC) (**D**, **right panel**). Box plots display a group of numerical data through their 3^rd^ and 1^st^ quartiles (box), mean (central band), minimum and maximum (whiskers). Each dot represents one sample, each patient being drawn 1 time for cancer free individuals and 1-2 times for cancer patients. Statistical analyses used one-way ANOVA with Kenward-Roger method to take into account the number of specimen/patient (A-B-C left panel): \**p*<0.05, \*\**p*<0.01), non-parametric unpaired Wilcoxon test (Mann-Whitney) for each two-group comparison (D left panel): \**p*<0.05, \*\**p*< 0.01, \*\*\**p*< 0.001, \*\*\*\**p*<0.0001.

## Supplemental tables

**Table S1.** Clinical and viral characteristics of patients COVID-19^+^ enrolling in ONCOVID. Clinical and viral characteristics of patients COVID-19^+^ enrolling in three French COVID-19^+^ and one Canadian cohort according to the duration of viral shedding (SVS *vs* LVS).

**Table S2.** Clinical characteristics of Cancer_FR1 available for translational research

### Supplementary Material, Figures and Tables

**Supplementary Material, Figure 1.** Strategy of flow cytometry analysis. Briefly, manual gating was performed to extract fcs files of CD45^+^, CD3^+^ T cells and CD19^+^ B cells using the myeloid, T cell and B cell panels, respectively. An example of gating strategy was shown on one sample. Fcs files were compilated in uniform manifold approximation and projection (UMAP) representation. Concatenation of all the samples was shown on the UMAP representation. Relative expression of markers was used to gate the population of interest.

**Supplementary Material, Figure 2.** Relative expression of principal markers used to gate the populations explored with the myeloid panel. An example of gating strategy of monocytes, as well as CD169 and HLA-DR, was shown in one sample.

**Supplementary Material, Figure 3. A.** Relative expression of principal markers used to gate the populations explored with the T cell panel (**A, top panel**). An example of gating strategy of follicular helper T cells and the activated subset was shown on one sample (**A, bottom panel**). **B.** Relative expression of principal markers used to gate the populations explored with the B cell panel.

**Supplementary Material, Table 2.**
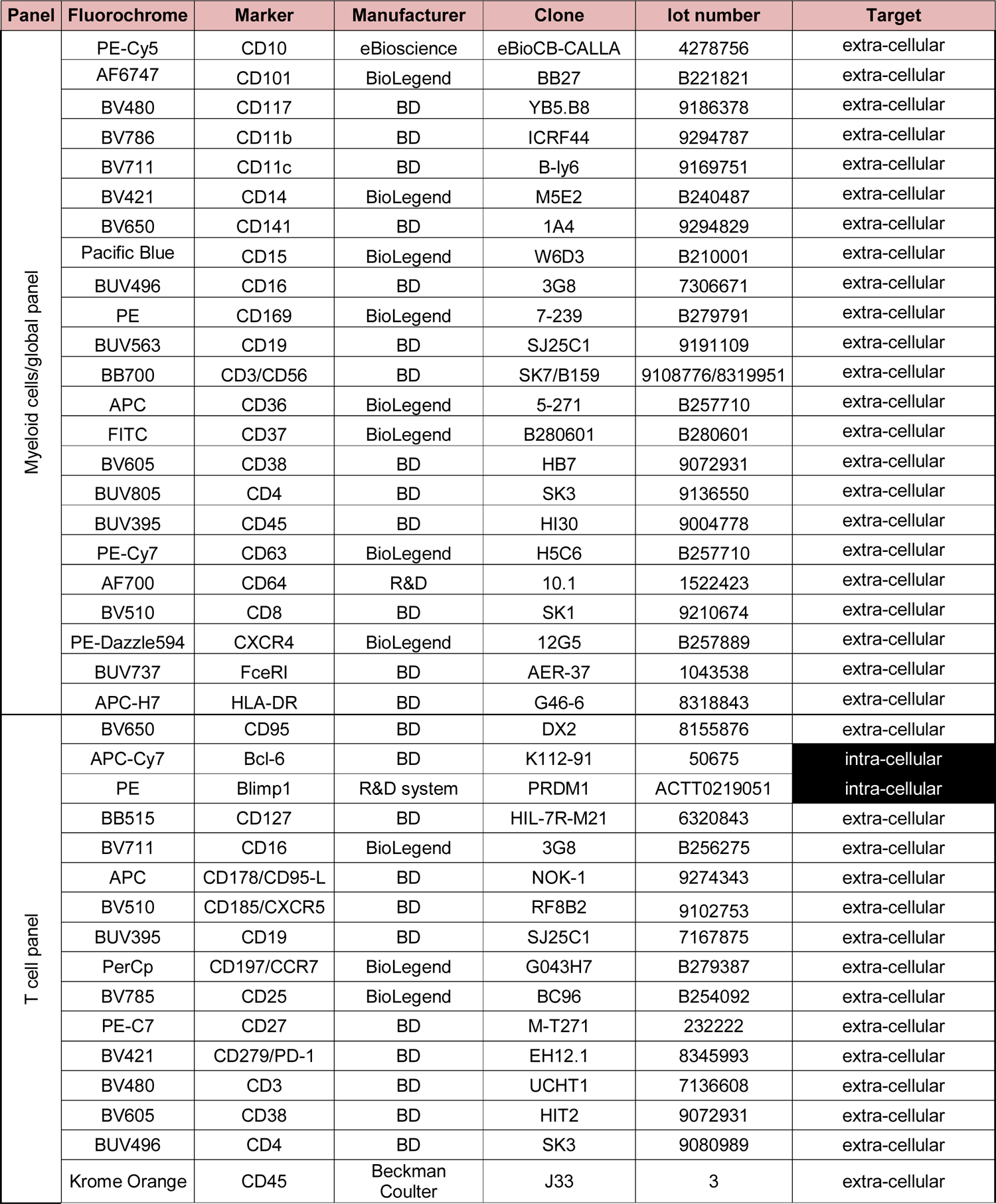
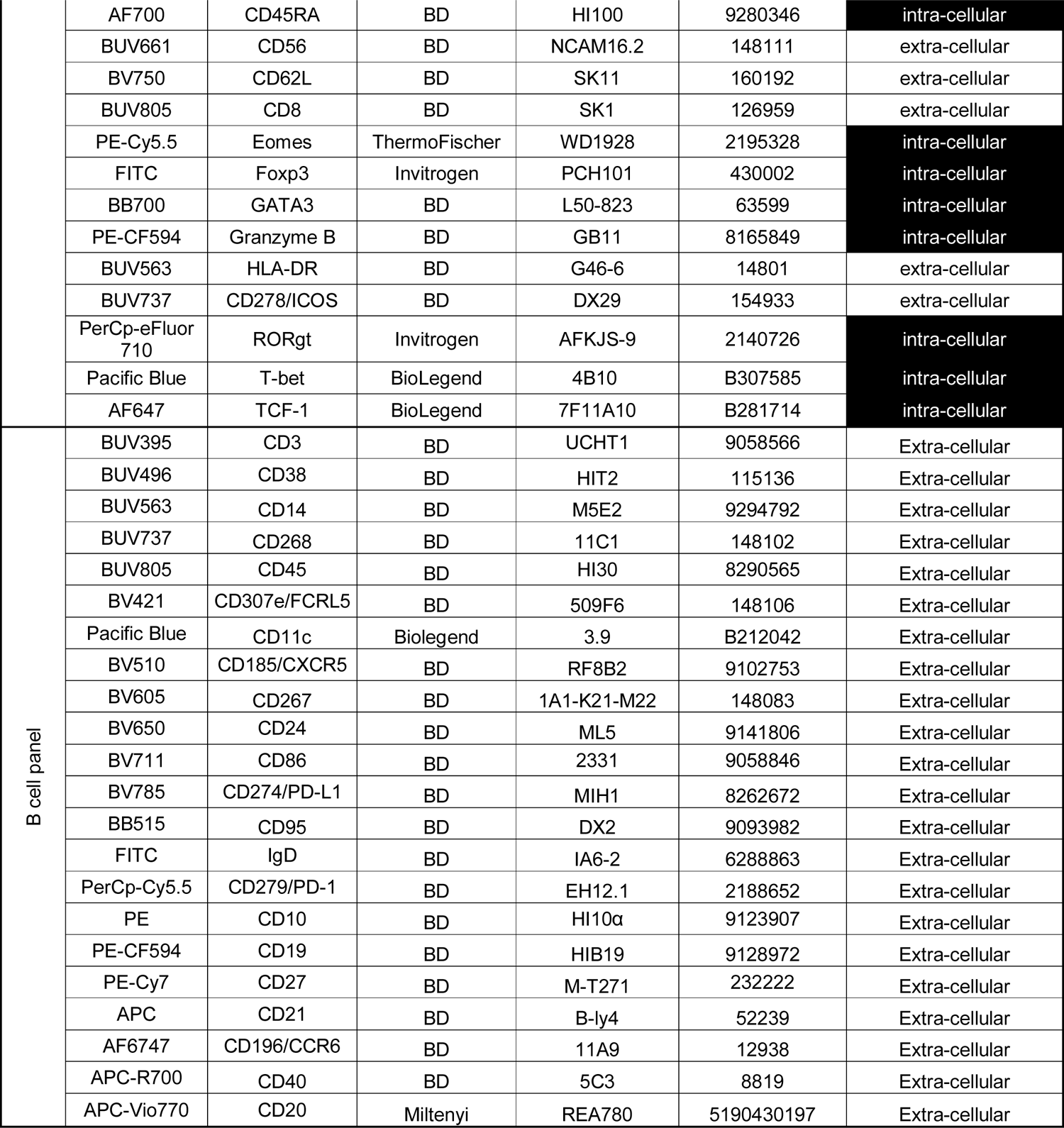
List of monoclonal antibodies used for the stainings.

| Panel | Fluorochrome | Marker | Manufacturer | Clone | lot number | Target |
| --- | --- | --- | --- | --- | --- | --- |
| Myeloid cells/global panel | PE-Cy5 | CD10 | eBioscience | eBioCB-CALLA | 4278756 | extra-cellular |
|  | AF6747 | CD101 | BioLegend | BB27 | B221821 | extra-cellular |
|  | BV480 | CD117 | BD | YB5.B8 | 9186378 | extra-cellular |
|  | BV786 | CD11b | BD | ICRF44 | 9294787 | extra-cellular |
|  | BV711 | CD11c | BD | B-ly6 | 9169751 | extra-cellular |
|  | BV421 | CD14 | BioLegend | M5E2 | B240487 | extra-cellular |
|  | BV650 | CD141 | BD | 1A4 | 9294829 | extra-cellular |
|  | Pacific Blue | CD15 | BioLegend | W6D3 | B210001 | extra-cellular |
|  | BUV496 | CD16 | BD | 3G8 | 7306671 | extra-cellular |
|  | PE | CD169 | BioLegend | 7-239 | B279791 | extra-cellular |
|  | BUV563 | CD19 | BD | SJ25C1 | 9191109 | extra-cellular |
|  | BB700 | CD3/CD56 | BD | SK7/B159 | 9108776/8319951 | extra-cellular |
|  | APC | CD36 | BioLegend | 5-271 | B257710 | extra-cellular |
|  | FITC | CD37 | BioLegend | B280601 | B280601 | extra-cellular |
|  | BV605 | CD38 | BD | HB7 | 9072931 | extra-cellular |
|  | BUV805 | CD4 | BD | SK3 | 9136550 | extra-cellular |
|  | BUV395 | CD45 | BD | HI30 | 9004778 | extra-cellular |
|  | PE-Cy7 | CD63 | BioLegend | H5C6 | B257710 | extra-cellular |
|  | AF700 | CD64 | R&D | 10.1 | 1522423 | extra-cellular |
|  | BV510 | CD8 | BD | SK1 | 9210674 | extra-cellular |
|  | PE-Dazzle594 | CXCR4 | BioLegend | 12G5 | B257889 | extra-cellular |
|  | BUV737 | FceRI | BD | AER-37 | 1043538 | extra-cellular |
|  | APC-H7 | HLA-DR | BD | G46-6 | 8318843 | extra-cellular |
| T cell panel | BV650 | CD95 | BD | DX2 | 8155876 | extra-cellular |
|  | APC-Cy7 | Bcl-6 | BD | K112-91 | 50675 | intra-cellular |
|  | PE | Blimp1 | R&D system | PRDM1 | ACTT0219051 | intra-cellular |
|  | BB515 | CD127 | BD | HIL-7R-M21 | 6320843 | extra-cellular |
|  | BV711 | CD16 | BioLegend | 3G8 | B256275 | extra-cellular |
|  | APC | CD178/CD95-L | BD | NOK-1 | 9274343 | extra-cellular |
|  | BV510 | CD185/CXCR5 | BD | RF8B2 | 9102753 | extra-cellular |
|  | BUV395 | CD19 | BD | SJ25C1 | 7167875 | extra-cellular |
|  | PerCp | CD197/CCR7 | BioLegend | G043H7 | B279387 | extra-cellular |
|  | BV785 | CD25 | BioLegend | BC96 | B254092 | extra-cellular |
|  | PE-C7 | CD27 | BD | M-T271 | 232222 | extra-cellular |
|  | BV421 | CD279/PD-1 | BD | EH12.1 | 8345993 | extra-cellular |
|  | BV480 | CD3 | BD | UCHT1 | 7136608 | extra-cellular |
|  | BV605 | CD38 | BD | HIT2 | 9072931 | extra-cellular |
|  | BUV496 | CD4 | BD | SK3 | 9080989 | extra-cellular |
|  | Krome Orange | CD45 | Beckman Coulter | J33 | 3 | extra-cellular |
|  | AF700 | CD45RA | BD | HI100 | 9280346 | intra-cellular |
|  | BUV661 | CD56 | BD | NCAM16.2 | 148111 | extra-cellular |
|  | BV750 | CD62L | BD | SK11 | 160192 | extra-cellular |
|  | BUV805 | CD8 | BD | SK1 | 126959 | extra-cellular |
|  | PE-Cy5.5 | Eomes | ThermoFischer | WD1928 | 2195328 | intra-cellular |
|  | FITC | Foxp3 | Invitrogen | PCH101 | 430002 | intra-cellular |
|  | BB700 | GATA3 | BD | L50-823 | 63599 | intra-cellular |
|  | PE-CF594 | Granzyme B | BD | GB11 | 8165849 | intra-cellular |
|  | BUV563 | HLA-DR | BD | G46-6 | 14801 | extra-cellular |
|  | BUV737 | CD278/ICOS | BD | DX29 | 154933 | extra-cellular |
|  | PerCp-eFluor 710 | RORgt | Invitrogen | AFKJS-9 | 2140726 | intra-cellular |
|  | Pacific Blue | T-bet | BioLegend | 4B10 | B307585 | intra-cellular |
|  | AF647 | TCF-1 | BioLegend | 7F11A10 | B281714 | intra-cellular |
| B cell panel | BUV395 | CD3 | BD | UCHT1 | 9058566 | Extra-cellular |
|  | BUV496 | CD38 | BD | HIT2 | 115136 | Extra-cellular |
|  | BUV563 | CD14 | BD | M5E2 | 9294792 | Extra-cellular |
|  | BUV737 | CD268 | BD | 11C1 | 148102 | Extra-cellular |
|  | BUV805 | CD45 | BD | HI30 | 8290565 | Extra-cellular |
|  | BV421 | CD307e/FCRL5 | BD | 509F6 | 148106 | Extra-cellular |
|  | Pacific Blue | CD11c | Biolegend | 3.9 | B212042 | Extra-cellular |
|  | BV510 | CD185/CXCR5 | BD | RF8B2 | 9102753 | Extra-cellular |
|  | BV605 | CD267 | BD | 1A1-K21-M22 | 148083 | Extra-cellular |
|  | BV650 | CD24 | BD | ML5 | 9141806 | Extra-cellular |
|  | BV711 | CD86 | BD | 2331 | 9058846 | Extra-cellular |
|  | BV785 | CD274/PD-L1 | BD | MIH1 | 8262672 | Extra-cellular |
|  | BB515 | CD95 | BD | DX2 | 9093982 | Extra-cellular |
|  | FITC | IgD | BD | IA6-2 | 6288863 | Extra-cellular |
|  | PerCp-Cy5.5 | CD279/PD-1 | BD | EH12.1 | 2188652 | Extra-cellular |
|  | PE | CD10 | BD | HI10α | 9123907 | Extra-cellular |
|  | PE-CF594 | CD19 | BD | HIB19 | 9128972 | Extra-cellular |
|  | PE-Cy7 | CD27 | BD | M-T271 | 232222 | Extra-cellular |
|  | APC | CD21 | BD | B-ly4 | 52239 | Extra-cellular |
|  | AF6747 | CD196/CCR6 | BD | 11A9 | 12938 | Extra-cellular |
|  | APC-R700 | CD40 | BD | 5C3 | 8819 | Extra-cellular |
|  | APC-Vio770 | CD20 | Miltenyi | REA780 | 5190430197 | Extra-cellular |

**Figure 6**
| Discovery cohort |  |  |  |  |  |  |  |
| --- | --- | --- | --- | --- | --- | --- | --- |
| Cancer patients' characteristics |  | Cancer_FR1_TR+<br>Cancer_FR1_CR+<br>Cancer_FR3<br>(n=110) | Ct>25 &<br>ALC>800<br>(n=36) | Ct>25 &<br>ALC<800<br>(n=22) | Ct<25 &<br>ALC>800<br>(n=29) | Ct<25 &<br>ALC<800<br>(n=23) | P |
| Age (year) | Median (range) | 62 (13-95) | 62 (13-82) | 63 (20-83) | 59 (38-95) | 60 (21-84) | 0.76<sup>#</sup> |
| Gender–no(%) | Male | 46 (42) | 18 (50) | 9 (41) | 13 (45) | 6 (26) | 0.33 |
|  | Female | 64 (58) | 18 (50) | 13 (59) | 16 (55) | 17 (74) |  |
| Number of comorbidities<br>–no(%)<sup>o</sup> | 0 | 38 (45) | 10 (45) | 5 (34) | 10 (38) | 13 (62) | 0.26 |
|  | 1 | 25 (30) | 5 (23) | 6 (40) | 10 (38) | 4 (19) |  |
|  | 2 | 16 (19) | 4 (18) | 2 (13) | 6 (24) | 4 (19) |  |
|  | 3 | 5 (6) | 3 (14) | 2 (13) | 0 (0) | 0 (0) |  |
| Comorbidities<br>–no(%)<sup>o</sup> | COPD | 6 (7) | 2 (9) | 1 (7) | 1 (4) | 2 (10) | 0.98 |
|  | BMI ≥ 30 | 12 (14) | 2 (9) | 3 (20) | 4 (15) | 3 (14) |  |
|  | Hypertension | 32 (38) | 11 (50) | 7 (47) | 8 (31) | 6 (29) |  |
|  | Congestive heart failure | 3 (6) | 1 (5) | 1 (7) | 1 (4) | 0 (0) |  |
|  | Diabetes mellitus | 10 (12) | 3 (14) | 1 (7) | 4 (15) | 2 (10) |  |
| Type of malignancy<br>–no(%) | S | 92 (84) | 33 (92) | 15 (68) | 26 (90) | 18 (78) | 0.08 |
|  | H | 18 (16) | 3 (8) | 7 (32) | 3 (10) | 5 (22) |  |
| Cancer spread<br>–no(%) | Localized | 19 (17) | 7 (19) | 1 (5) | 7 (24) | 4 (17) | 0.46 |
|  | Locally advanced | 24 (22) | 9 (25) | 6 (27) | 3 (10) | 6 (26) |  |
|  | Metastatic | 67 (61) | 20 (56) | 15 (68) | 19 (66) | 13 (57) |  |
| Cancer status<br>–no(%) | Remission or NED | 29 (26) | 12 (30) | 3 (14) | 10 (34) | 4 (17) | 0.21 |
|  | SD/PD | 47 (43) | 17 (47) | 11 (50) | 11 (38) | 8 (35) |  |
|  | Present or PD | 34 (31) | 7 (19) | 8 (36) | 8 (28) | 11 (48) |  |
| ECOG PS– no(%) | 0 | 28 (25) | 13 (36) | 5 (23) | 5 (18) | 5 (22) | 0.01 |
|  | 1 | 46 (42) | 18 (50) | 4 (18) | 12 (41) | 12 (52) |  |
|  | 2 or more | 36 (33) | 5 (14) | 13 (59) | 12 (41) | 6 (26) |  |
| Type of anticancer therapy–<br>no(%) | None* | 53 (48) | 20 (56) | 8 (36) | 14 (48) | 10 (43) | 0.53 |
|  | Chemotherapy | 47 (43) | 4 (11) | 12 (55) | 11 (38) | 14 (61) | 0.19 |
|  | Radiotherapy | 8 (7) | 2 (6) | 3 (14) | 1 (3) | 2 (9) |  |
|  | Surgery | 8 (7) | 3 (8) | 2 (9) | 3 (10) | 0 (0) |  |
|  | Hormonal therapy | 11 (10) | 4 (11) | 0 | 4 (14) | 3 (13) |  |
|  | Immunotherapy | 12 (11) | 4 (11) | 1 (5) | 4 (14) | 3 (13) |  |
|  | Others | 11 (10) | 2 (6) | 2 (9) | 0 (0) | 5 (22) |  |
| Delay of treatment–no(%)<sup>o</sup> | Yes (range: 16-170 days) | 12 (32) | 2 (33) | 2 (22) | 8 (67) | 0 (0) | <0.01 |
|  | No | 26 (68) | 4 (67) | 7 (78) | 4 (33) | 11 (100) |  |
| Clinical course<br>–no(%)<sup>o</sup> | Day hospital | 27 (32) | 10 (45) | 4 (27) | 8 (31) | 5 (24) | 0.63 |
|  | Hospitalization | 53 (63) | 12 (55) | 10 (67) | 17 (65) | 14 (67) |  |
|  | Admission to ICU | 4 (5) | 0 | 1 (6) | 1 (4) | 2 (9) |  |
| Death–no(%) | Yes | 31 (28) | 4 (11) | 7 (32) | 9 (31) | 11 (48) | 0.02 |
| Validation cohort |  |  |  |  |  |  |  |
| Cancer patients' characteristics |  | Cancer_FR1_CR+<br>Cancer_FR3<br>(n=116) | Ct>25 &<br>ALC>800<br>(n=22) | Ct>25 &<br>ALC<800<br>(n=27) | Ct<25 &<br>ALC>800<br>(n=29) | Ct<25 &<br>ALC<800<br>(n=38) | P |
| Age (year) | Median (range) | 65 (13-91) | 55 (13-86) | 64 (46-77) | 68 (41-84) | 66 (18-91) | 0.09<sup>#</sup> |
| Gender–no(%) | Male | 71 (61) | 9 (41) | 14 (52) | 17 (59) | 23 (61) | 0.48 |
|  | Female | 45 (39) | 13 (59) | 13 (48) | 12 (41) | 15 (39) |  |
| Type of malignancy<br>–no(%) | S | 85 (73) | 19 (86) | 19 (70) | 22 (76) | 25 (66) | 0.36 |
|  | H | 31 (27) | 3 (14) | 8 (30) | 7 (24) | 13 (34) |  |
| Cancer spread<br>–no(%) | Localized | 9 (8) | 3 (14) | 1 (4) | 3 (10) | 2 (5.3) | 0.40 |
|  | Locally advanced | 15 (13) | 4 (18) | 6 (22) | 3 (10) | 2 (5.3) |  |
|  | Metastatic | 82 (70) | 15 (68) | 17 (63) | 21 (72) | 29 (76.4) |  |
|  | Unknown | 10 (9) | 0 (0) | 3 (11) | 2 (8) | 5 (13) |  |
| Type of anticancer therapy–<br>no(%)<sup>o*</sup> | None* | 33 (36) | 14 (74) | 8 (36) | 5 (23) | 6 (21) | 0.001 |
|  | Chemotherapy | 28 (31) | 1 (5) | 11 (50) | 5 (23) | 11 (39) | 0.17 |
|  | Radiotherapy | 5 (5) | 0 (0) | 2 (9) | 1 (5) | 2 (7) |  |
|  | Surgery | 1 (1) | 0 (0) | 0 (0) | 1 (5) | 0 (0) |  |
|  | Hormonal therapy | 4 (4) | 2 (11) | 1 (5) | 0 (0) | 1 (4) |  |
|  | Immunotherapy | 20 (22) | 2 (11) | 2 (9) | 6 (27) | 10 (36) |  |
|  | Others | 17 (19) | 1 (5) | 4 (18) | 5 (23) | 7 (25) |  |
| Death–no(%) | Yes | 27 (23) | 1 (4) | 5 (18) | 6 (21) | 15 (39) | 0.016 |

## Abbreviations in alphabetic order

ACE2: Angiotensin-Converting Enzyme 2

CD: cluster of differentiation

cMo: classical monocytes

COVID-19: severe coronavirus infectious disease 2019

Ct: Cycle threshold

CR: Clinical Routine

CXCR5: C-X-C chemokine receptor type 5

ECOG: Eastern Cooperative Oncology Group performance status

EM: effector memory cells

Eomes: eomesodermin

FC: fold change

FasL: Fas ligand

GzB: granzyme B

HCW: health care worker

ICOS: Inducible T-cell co-stimulator

ICU: intensive care unit

IFN: Interferon

Ig: Immunoglobulin

IL: interleukin

LVS: long term viral RNA shedding

NK: Natural killer

non-cMo: non classical monocytes

NSM: Non-switch Memory

Orf1b: replicase open reading frame 1b

PD: progressive disease

PD-1: programmed cell death 1

PMN: polymorphonuclear cells

RBD: receptor-binding domain

RdRP: RNA-dependent RNA polymerase

RNA: ribonucleic acid

rIL-7: recombinant interleukin 7

RT-qPCR: quantitative reverse transcription-polymerase chain reaction

SARS-CoV-2: severe acute respiratory syndrome coronavirus 2

SD: stable disease

SVS: short term viral RNA shedding

Tc1: cytotoxic T lymphocytes

TCF-1: Transcription factor 1

TFH: T follicular helper cells

TR: Translational Research.

## Notes

### Competing Interest Statement

The authors have declared no competing interest.

### Clinical Trial

NCT04341207

### Author Declarations

All patients provided written informed consent. Protocol approval was obtained from an independent ethics committee (ethics protocol number EudraCT No: 2020-001250-21). Gustave Roussy Data Protection Officer (DPO) has evaluated this project and sent to the principal investigator a formalized-operational action plan about data protection compliance: patient's information, security measures, good practices about pseudonymization, etc. All of the DPO's recommendations has been applied by the research team.In agreement with MR004 in France, we reported the series to the national information science and liberties commission. Ethic committees: The Agence Nationale de securite du medicament et des produits de sante (FRANCE) gave the approval for the clinical trial No. EudraCT 2020-001250-21 (reference: MEDAECPP-2020-03-0012). The Committee of the Convention for the Protection of Individuals CPP Ile de France XI (FRANCE) approved the clinical trail (reference: 20025-75647). Decision made: CPP Ile de France XI and Agence Nationale de securite du medicament et des produits de sante authorized Gustave Roussy (No. CSET 2020/3078) for the clinical trial ONCOVID.

## References

1. Albiges, L., Foulon, S., Bayle, A., Gachot, B., Pommeret, F., Willekens, C., Stoclin, A., Merad, M., Griscelli, F., Lacroix, L., Netzer, F., Hueso, T., Balleyguier, C., Ammari, S., Colomba, E., Baciarello, G., Perret, A., Hollebecque, A., Hadoux, J., Michot, J.-M., Chaput, N., Saada, V., Hauchecorne, M., Micol, J.-B., Sun, R., Valteau-Couanet, D., André, F., Scotte, F., Besse, B., Soria, J.-C., Barlesi, F., 2020. Determinants of the outcomes of patients with cancer infected with SARS-CoV-2: results from the Gustave Roussy cohort. Nature Cancer 1, 965–975. https://doi.org/10.1038/s43018-020-00120-5

2. Alsaleh, G., Panse, I., Swadling, L., Zhang, H., Richter, F.C., Meyer, A., Lord, J., Barnes, E., Klenerman, P., Green, C., Simon, A.K., 2020. Autophagy in T cells from aged donors is maintained by spermidine and correlates with function and vaccine responses. Elife 9. https://doi.org/10.7554/eLife.57950

3. Amrane, S., Tissot-Dupont, H., Doudier, B., Eldin, C., Hocquart, M., Mailhe, M., Dudouet, P., Ormières, E., Ailhaud, L., Parola, P., Lagier, J.-C., Brouqui, P., Zandotti, C., Ninove, L., Luciani, L., Boschi, C., La Scola, B., Raoult, D., Million, M., Colson, P., Gautret, P., 2020. Rapid viral diagnosis and ambulatory management of suspected COVID-19 cases presenting at the infectious diseases referral hospital in Marseille, France, - January 31st to March 1st, 2020: A respiratory virus snapshot. Travel Med Infect Dis 36, 101632. https://doi.org/10.1016/j.tmaid.2020.101632

4. Anhê, F.F., Jensen, B.A.H., Varin, T.V., Servant, F., Van Blerk, S., Richard, D., Marceau, S., Surette, M., Biertho, L., Lelouvier, B., Schertzer, J.D., Tchernof, A., Marette, A., 2020. Type 2 diabetes influences bacterial tissue compartmentalisation in human obesity. Nature Metabolism 2, 233–242. https://doi.org/10.1038/s42255-020-0178-9

5. Arunachalam, P.S., Wimmers, F., Mok, C.K.P., Perera, R.A.P.M., Scott, M., Hagan, T., Sigal, N., Feng, Y., Bristow, L., Tsang, O.T.-Y., Wagh, D., Coller, J., Pellegrini, K.L., Kazmin, D., Alaaeddine, G., Leung, W.S., Chan, J.M.C., Chik, T.S.H., Choi, C.Y.C., Huerta, C., McCullough, M.P., Lv, H., Anderson, E., Edupuganti, S., Upadhyay, A.A., Bosinger, S.E., Maecker, H.T., Khatri, P., Rouphael, N., Peiris, M., Pulendran, B., 2020. Systems biological assessment of immunity to mild versus severe COVID-19 infection in humans. Science 369, 1210–1220. https://doi.org/10.1126/science.abc6261

6. Assaad, S., Avrillon, V., Fournier, M.-L., Mastroianni, B., Russias, B., Swalduz, A., Cassier, P., Eberst, L., Steineur, M.-P., Kazes, M., Perol, M., Michallet, A.-S., Rey, P., Erena-Penet, A.-S., Morel, A., Brahmi, M., Dufresne, A., Tredan, O., Chvetzoff, G., Fayette, J., de la Fouchardiere, C., Ray-Coquard, I., Bachelot, T., Saintigny, P., Tabutin, M., Dupré, A., Nicolas-Virelizier, E., Belhabri, A., Roux, P.-E., Fuhrmann, C., Pilleul, F., Basle, A., Bouhamama, A., Galvez, C., Herr, A.-L., Gautier, J., Chabaud, S., Zrounba, P., Perol, D., Blay, J.-Y., 2020. High mortality rate in cancer patients with symptoms of COVID-19 with or without detectable SARS-COV-2 on RT-PCR. European Journal of Cancer 135, 251–259. https://doi.org/10.1016/j.ejca.2020.05.028

7. Avanzato, V.A., Matson, M.J., Seifert, S.N., Pryce, R., Williamson, B.N., Anzick, S.L., Barbian, K., Judson, S.D., Fischer, E.R., Martens, C., Bowden, T.A., de Wit, E., Riedo, F.X., Munster, V.J., 2020. Case Study: Prolonged Infectious SARS-CoV-2 Shedding from an Asymptomatic Immunocompromised Individual with Cancer. Cell 183, 1901–1912.e9. https://doi.org/10.1016/j.cell.2020.10.049

8. Aydillo, T., Gonzalez-Reiche, A.S., Aslam, S., van de Guchte, A., Khan, Z., Obla, A., Dutta, J., van Bakel, H., Aberg, J., García-Sastre, A., Shah, G., Hohl, T., Papanicolaou, G., Perales, M.-A., Sepkowitz, K., Babady, N.E., Kamboj, M., 2020. Shedding of Viable SARS-CoV-2 after Immunosuppressive Therapy for Cancer. N Engl J Med 383, 2586–2588. https://doi.org/10.1056/NEJMc2031670

9. Becht, E., McInnes, L., Healy, J., Dutertre, C.-A., Kwok, I.W.H., Ng, L.G., Ginhoux, F., Newell, E.W., 2019. Dimensionality reduction for visualizing single-cell data using UMAP. Nature Biotechnology 37, 38–44. https://doi.org/10.1038/nbt.4314

10. Bitterman, R., Eliakim Raz, N., Vinograd, I., Trestioreanu, A.Z., Leibovici, L., Paul, M., 2018. Influenza vaccines in immunosuppressed adults with cancer. Cochrane Database of Systematic Reviews. https://doi.org/10.1002/14651858.CD008983.pub3

11. Boutin, C.-A., Grandjean-Lapierre, S., Gagnon, S., Labbé, A.-C., Charest, H., Roger, M., Coutlée, F., 2020. Comparison of SARS-CoV-2 detection from combined nasopharyngeal/oropharyngeal swab samples by a laboratory-developed real-time RT-PCR test and the Roche SARS-CoV-2 assay on a cobas 8800 instrument. J Clin Virol 132, 104615. https://doi.org/10.1016/j.jcv.2020.104615

12. Buja, L.M., Wolf, D.A., Zhao, B., Akkanti, B., McDonald, M., Lelenwa, L., Reilly, N., Ottaviani, G., Elghetany, M.T., Trujillo, D.O., Aisenberg, G.M., Madjid, M., Kar, B., 2020. The emerging spectrum of cardiopulmonary pathology of the coronavirus disease 2019 (COVID-19): Report of 3 autopsies from Houston, Texas, and review of autopsy findings from other United States cities. Cardiovasc Pathol 48, 107233. https://doi.org/10.1016/j.carpath.2020.107233

13. Campbell, C., McKenney, P.T., Konstantinovsky, D., Isaeva, O.I., Schizas, M., Verter, J., Mai, C., Jin, W.-B., Guo, C.-J., Violante, S., Ramos, R.J., Cross, J.R., Kadaveru, K., Hambor, J., Rudensky, A.Y., 2020. Bacterial metabolism of bile acids promotes generation of peripheral regulatory T cells. Nature 581, 475–479. https://doi.org/10.1038/s41586-020-2193-0

14. Carvelli, J., Demaria, O., Vély, F., Batista, L., Benmansour, N.C., Fares, J., Carpentier, S., Thibult, M.-L., Morel, A., Remark, R., André, P., Represa, A., Piperoglou, C., Cordier, P.Y., Dault, E.L., Guervilly, C., Simeone, P., Gainnier, M., Morel, Y., Ebbo, M., Schleinitz, N., Vivier, E., 2020. Association of COVID-19 inflammation with activation of the C5a–C5aR1 axis. Nature 1–5. https://doi.org/10.1038/s41586-020-2600-6

15. Channappanavar, R., Perlman, S., 2017. Pathogenic human coronavirus infections: causes and consequences of cytokine storm and immunopathology. Semin Immunopathol 39, 529–539. https://doi.org/10.1007/s00281-017-0629-x

16. Cheng, L.-L., Guan, W.-J., Duan, C.-Y., Zhang, N.-F., Lei, C.-L., Hu, Y., Chen, A.-L., Li, S.-Y., Zhuo, C., Deng, X.-L., Cheng, F.-J., Gao, Y., Zhang, J.-H., Xie, J.-X., Peng, Hong, Li, Y.-X., Wu, X.-X., Liu, W., Peng, Hui, Wang, J., Xiao, G.-M., Chen, P.-Y., Wang, C.-Y., Yang, Z.-F., Zhao, J.-C., Zhong, N.-S., 2020. Effect of Recombinant Human Granulocyte Colony-Stimulating Factor for Patients With Coronavirus Disease 2019 (COVID-19) and Lymphopenia: A Randomized Clinical Trial. JAMA Intern Med. https://doi.org/10.1001/jamainternmed.2020.5503

17. Choi, B., Choudhary, M.C., Regan, J., Sparks, J.A., Padera, R.F., Qiu, X., Solomon, I.H., Kuo, H.-H., Boucau, J., Bowman, K., Adhikari, U.D., Winkler, M.L., Mueller, A.A., Hsu, T.Y.-T., Desjardins, M., Baden, L.R., Chan, B.T., Walker, B.D., Lichterfeld, M., Brigl, M., Kwon, D.S., Kanjilal, S., Richardson, E.T., Jonsson, A.H., Alter, G., Barczak, A.K., Hanage, W.P., Yu, X.G., Gaiha, G.D., Seaman, M.S., Cernadas, M., Li, J.Z., 2020. Persistence and Evolution of SARS-CoV-2 in an Immunocompromised Host. N Engl J Med. https://doi.org/10.1056/NEJMc2031364

18. Chua, R.L., Lukassen, S., Trump, S., Hennig, B.P., Wendisch, D., Pott, F., Debnath, O., Thürmann, L., Kurth, F., Völker, M.T., Kazmierski, J., Timmermann, B., Twardziok, S., Schneider, S., Machleidt, F., Müller-Redetzky, H., Maier, M., Krannich, A., Schmidt, S., Balzer, F., Liebig, J., Loske, J., Suttorp, N., Eils, J., Ishaque, N., Liebert, U.G., von Kalle, C., Hocke, A., Witzenrath, M., Goffinet, C., Drosten, C., Laudi, S., Lehmann, I., Conrad, C., Sander, L.-E., Eils, R., 2020. COVID-19 severity correlates with airway epithelium–immune cell interactions identified by single-cell analysis. Nature Biotechnology 38, 970–979. https://doi.org/10.1038/s41587-020-0602-4

19. Corman, V.M., Landt, O., Kaiser, M., Molenkamp, R., Meijer, A., Chu, D.K., Bleicker, T., Brünink, S., Schneider, J., Schmidt, M.L., Mulders, D.G., Haagmans, B.L., van der Veer, B., van den Brink, S., Wijsman, L., Goderski, G., Romette, J.-L., Ellis, J., Zambon, M., Peiris, M., Goossens, H., Reusken, C., Koopmans, M.P., Drosten, C., 2020. Detection of 2019 novel coronavirus (2019-nCoV) by real-time RT-PCR. Euro Surveill 25. https://doi.org/10.2807/1560-7917.ES.2020.25.3.2000045

20. Dai, M., Liu, D., Liu, M., Zhou, F., Li, G., Chen, Z., Zhang, Z., You, H., Wu, M., Zheng, Q., Xiong, Y., Xiong, H., Wang, C., Chen, C., Xiong, F., Zhang, Y., Peng, Y., Ge, S., Zhen, B., Yu, T., Wang, L., Wang, H., Liu, Y., Chen, Y., Mei, J., Gao, X., Li, Zhuyan, Gan, L., He, C., Li, Zhen, Shi, Y., Qi, Y., Yang, J., Tenen, D.G., Chai, L., Mucci, L.A., Santillana, M., Cai, H., 2020. Patients with Cancer Appear More Vulnerable to SARS-CoV-2: A Multicenter Study during the COVID-19 Outbreak. Cancer Discov 10, 783–791. https://doi.org/10.1158/2159-8290.CD-20-0422

21. Danlos, F.-X., Grajeda-Iglesias, C., Durand, S., Sauvat, A., Roumier, M., Cantin, D., Colomba, E., Rohmer, J., Pommeret, F., Baciarello, G., et al., In Press. Metabolomic analyses of COVID-19 patients unravel stage-dependent and prognostic biomarkers. Cell Death & Disease.

22. Derosa, L., Melenotte, C., Griscelli, F., Gachot, B., Marabelle, A., Kroemer, G., Zitvogel, L., 2020. The immuno-oncological challenge of COVID-19. Nature Cancer 1, 946–964. https://doi.org/10.1038/s43018-020-00122-3

23. El Ramahi, R., Freifeld, A., 2019. Epidemiology, Diagnosis, Treatment, and Prevention of Influenza Infection in Oncology Patients. JOP 15, 177–184. https://doi.org/10.1200/JOP.18.00567

24. Elkrief, A., Desilets, A., Papneja, N., Cvetkovic, L., Groleau, C., Lakehal, Y.A., Shbat, L., Richard, C., Malo, J., Belkaid, W., Cook, E., Doucet, S., Tran, T.H., Jao, K., Daaboul, N., Bhang, E., Loree, J.M., Miller, W.H., Vinh, D.C., Bouganim, N., Batist, G., Letendre, C., Routy, B., 2020. High mortality among hospital-acquired COVID-19 infection in patients with cancer: A multicentre observational cohort study. European Journal of Cancer 139, 181–187. https://doi.org/10.1016/j.ejca.2020.08.017

25. Escudié, F., Auer, L., Bernard, M., Mariadassou, M., Cauquil, L., Vidal, K., Maman, S., Hernandez-Raquet, G., Combes, S., Pascal, G., 2018. FROGS: Find, Rapidly, OTUs with Galaxy Solution. Bioinformatics 34, 1287–1294. https://doi.org/10.1093/bioinformatics/btx791

26. Fischer, K., Hoffmann, P., Voelkl, S., Meidenbauer, N., Ammer, J., Edinger, M., Gottfried, E., Schwarz, S., Rothe, G., Hoves, S., Renner, K., Timischl, B., Mackensen, A., Kunz-Schughart, L., Andreesen, R., Krause, S.W., Kreutz, M., 2007. Inhibitory effect of tumor cell-derived lactic acid on human T cells. Blood 109, 3812–3819. https://doi.org/10.1182/blood-2006-07-035972

27. Francois, B., Jeannet, R., Daix, T., Walton, A.H., Shotwell, M.S., Unsinger, J., Monneret, G., Rimmelé, T., Blood, T., Morre, M., Gregoire, A., Mayo, G.A., Blood, J., Durum, S.K., Sherwood, E.R., Hotchkiss, R.S., 2018. Interleukin-7 restores lymphocytes in septic shock: the IRIS-7 randomized clinical trial. JCI Insight 3. https://doi.org/10.1172/jci.insight.98960

28. Garassino, M.C., Whisenant, J.G., Huang, L.-C., Trama, A., Torri, V., Agustoni, F., Baena, J., Banna, G., Berardi, R., Bettini, A.C., Bria, E., Brighenti, M., Cadranel, J., De Toma, A., Chini, C., Cortellini, A., Felip, E., Finocchiaro, G., Garrido, P., Genova, C., Giusti, R., Gregorc, V., Grossi, F., Grosso, F., Intagliata, S., La Verde, N., Liu, S.V., Mazieres, J., Mercadante, E., Michielin, O., Minuti, G., Moro-Sibilot, D., Pasello, G., Passaro, A., Scotti, V., Solli, P., Stroppa, E., Tiseo, M., Viscardi, G., Voltolini, L., Wu, Y.-L., Zai, S., Pancaldi, V., Dingemans, A.-M., Van Meerbeeck, J., Barlesi, F., Wakelee, H., Peters, S., Horn, L., TERAVOLT investigators, 2020. COVID-19 in patients with thoracic malignancies (TERAVOLT): first results of an international, registry-based, cohort study. Lancet Oncol 21, 914–922. https://doi.org/10.1016/S1470-2045(20)30314-4

29. Ge, H., Wang, X., Yuan, X., Xiao, G., Wang, C., Deng, T., Yuan, Q., Xiao, X., 2020. The epidemiology and clinical information about COVID-19. Eur J Clin Microbiol Infect Dis 1–9. https://doi.org/10.1007/s10096-020-03874-z

30. Geis, S., Prifert, C., Weissbrich, B., Lehners, N., Egerer, G., Eisenbach, C., Buchholz, U., Aichinger, E., Dreger, P., Neben, K., Burkhardt, U., Ho, A.D., Kräusslich, H.-G., Heeg, K., Schnitzler, P., 2013. Molecular characterization of a respiratory syncytial virus outbreak in a hematology unit in Heidelberg, Germany. J Clin Microbiol 51, 155–162. https://doi.org/10.1128/JCM.02151-12

31. Helleberg, M., Niemann, C.U., Moestrup, K.S., Kirk, O., Lebech, A.-M., Lane, C., Lundgren, J., 2020. Persistent COVID-19 in an Immunocompromised Patient Temporarily Responsive to Two Courses of Remdesivir Therapy. J Infect Dis 222, 1103–1107. https://doi.org/10.1093/infdis/jiaa446

32. Jaafar, R., Aherfi, S., Wurtz, N., Grimaldier, C., Hoang, V.T., Colson, P., Raoult, D., La Scola, B., 2020. Correlation between 3790 qPCR positives samples and positive cell cultures including 1941 SARS-CoV-2 isolates. Clin Infect Dis. https://doi.org/10.1093/cid/ciaa1491

33. Kampen, J.J.A. van Vijver, D.A.M.C. van de Fraaij, P.L.A., Haagmans, B.L., Lamers, M.M., Okba, N., Akker J.P.C. van den, Endeman, H., Gommers, D.A.M.P.J., Cornelissen, J.J., Hoek, R.A.S., Eerden M.M. van der, Hesselink, D.A., Metselaar, H.J., Verbon, A., Steenwinkel, J.E.M. de Aron, G.I., Gorp, E.C.M. van Boheemen, S. van Voermans, J.C., Boucher, C.A.B., Molenkamp, R., Koopmans, M.P.G., Geurtsvankessel, C., Eijk A.A. van der, 2020. Shedding of infectious virus in hospitalized patients with coronavirus disease-2019 (COVID-19): duration and key determinants. medRxiv 2020.06.08.20125310. https://doi.org/10.1101/2020.06.08.20125310

34. Kaneko, N., Kuo, H.-H., Boucau, J., Farmer, J.R., Allard-Chamard, H., Mahajan, V.S., Piechocka-Trocha, A., Lefteri, K., Osborn, M., Bals, J., Bartsch, Y.C., Bonheur, N., Caradonna, T.M., Chevalier, J., Chowdhury, F., Diefenbach, T.J., Einkauf, K., Fallon, J., Feldman, J., Finn, K.K., Garcia-Broncano, P., Hartana, C.A., Hauser, B.M., Jiang, C., Kaplonek, P., Karpell, M., Koscher, E.C., Lian, X., Liu, H., Liu, J., Ly, N.L., Michell, A.R., Rassadkina, Y., Seiger, K., Sessa, L., Shin, S., Singh, N., Sun, W., Sun, X., Ticheli, H.J., Waring, M.T., Zhu, A.L., Alter, G., Li, J.Z., Lingwood, D., Schmidt, A.G., Lichterfeld, M., Walker, B.D., Yu, X.G., Padera, R.F., Pillai, S., 2020. Loss of Bcl-6-Expressing T Follicular Helper Cells and Germinal Centers in COVID-19. Cell 183, 143–157.e13. https://doi.org/10.1016/j.cell.2020.08.025

35. Lagier, J.-C., Million, M., Gautret, P., Colson, P., Cortaredona, S., Giraud-Gatineau, A., Honoré, S., Gaubert, J.-Y., Fournier, P.-E., Tissot-Dupont, H., Chabrière, E., Stein, A., Deharo, J.-C., Fenollar, F., Rolain, J.-M., Obadia, Y., Jacquier, A., La Scola, B., Brouqui, P., Drancourt, M., Parola, P., Raoult, D., 2020. Outcomes of 3,737 COVID-19 patients treated with hydroxychloroquine/azithromycin and other regimens in Marseille, France: A retrospective analysis. Travel Med Infect Dis 36, 101791. https://doi.org/10.1016/j.tmaid.2020.101791

36. Laing, A.G., Lorenc, A., del Molino del Barrio, I., Das, A., Fish, M., Monin, L., Muñoz-Ruiz, M., McKenzie, D.R., Hayday, T.S., Francos-Quijorna, I., Kamdar, S., Joseph, M., Davies, D., Davis, R., Jennings, A., Zlatareva, I., Vantourout, P., Wu, Y., Sofra, V., Cano, F., Greco, M., Theodoridis, E., Freedman, J., Gee, S., Chan, J.N.E., Ryan, S., Bugallo-Blanco, E., Peterson, P., Kisand, K., Haljasmägi, L., Chadli, L., Moingeon, P., Martinez, L., Merrick, B., Bisnauthsing, K., Brooks, K., Ibrahim, M.A.A., Mason, J., Lopez Gomez, F., Babalola, K., Abdul-Jawad, S., Cason, J., Mant, C., Seow, J., Graham, C., Doores, K.J., Di Rosa, F., Edgeworth, J., Shankar-Hari, M., Hayday, A.C., 2020. A dynamic COVID-19 immune signature includes associations with poor prognosis. Nature Medicine 26, 1623–1635. https://doi.org/10.1038/s41591-020-1038-6

37. Laterre, P.F., François, B., Collienne, C., Hantson, P., Jeannet, R., Remy, K.E., Hotchkiss, R.S., 2020. Association of Interleukin 7 Immunotherapy With Lymphocyte Counts Among Patients With Severe Coronavirus Disease 2019 (COVID-19). JAMA Netw Open 3. https://doi.org/10.1001/jamanetworkopen.2020.16485

38. Lax, S.F., Skok, K., Zechner, P., Kessler, H.H., Kaufmann, N., Koelblinger, C., Vander, K., Bargfrieder, U., Trauner, M., 2020. Pulmonary Arterial Thrombosis in COVID-19 With Fatal Outcome[: Results From a Prospective, Single-Center, Clinicopathologic Case Series. Ann Intern Med 173, 350–361. https://doi.org/10.7326/M20-2566

39. Lehners, N., Schnitzler, P., Geis, S., Puthenparambil, J., Benz, M.A., Alber, B., Luft, T., Dreger, P., Eisenbach, C., Kunz, C., Benner, A., Buchholz, U., Aichinger, E., Frank, U., Heeg, K., Ho, A.D., Egerer, G., 2013. Risk factors and containment of respiratory syncytial virus outbreak in a hematology and transplant unit. Bone Marrow Transplantation 48, 1548–1553. https://doi.org/10.1038/bmt.2013.94

40. Lehners, N., Tabatabai, J., Prifert, C., Wedde, M., Puthenparambil, J., Weissbrich, B., Biere, B., Schweiger, B., Egerer, G., Schnitzler, P., 2016. Long-Term Shedding of Influenza Virus, Parainfluenza Virus, Respiratory Syncytial Virus and Nosocomial Epidemiology in Patients with Hematological Disorders. PLOS ONE 11, e0148258. https://doi.org/10.1371/journal.pone.0148258

41. Li, Q., Guan, X., Wu, P., Wang, X., Zhou, L., Tong, Y., Ren, R., Leung, K.S.M., Lau, E.H.Y., Wong, J.Y., Xing, X., Xiang, N., Wu, Y., Li, C., Chen, Q., Li, D., Liu, T., Zhao, J., Liu, M., Tu, W., Chen, C., Jin, L., Yang, R., Wang, Q., Zhou, S., Wang, R., Liu, H., Luo, Y., Liu, Y., Shao, G., Li, H., Tao, Z., Yang, Y., Deng, Z., Liu, B., Ma, Z., Zhang, Y., Shi, G., Lam, T.T.Y., Wu, J.T., Gao, G.F., Cowling, B.J., Yang, B., Leung, G.M., Feng, Z., 2020. Early Transmission Dynamics in Wuhan, China, of Novel Coronavirus–Infected Pneumonia. New England Journal of Medicine 382, 1199–1207. https://doi.org/10.1056/NEJMoa2001316

42. Lluch, J., Servant, F., Païssé, S., Valle, C., Valière, S., Kuchly, C., Vilchez, G., Donnadieu, C., Courtney, M., Burcelin, R., Amar, J., Bouchez, O., Lelouvier, B., 2015. The Characterization of Novel Tissue Microbiota Using an Optimized 16S Metagenomic Sequencing Pipeline. PLOS ONE 10, e0142334. https://doi.org/10.1371/journal.pone.0142334

43. Luo, J., Rizvi, H., Preeshagul, I.R., Egger, J.V., Hoyos, D., Bandlamudi, C., McCarthy, C.G., Falcon, C.J., Schoenfeld, A.J., Arbour, K.C., Chaft, J.E., Daly, R.M., Drilon, A., Eng, J., Iqbal, A., Lai, W.V., Li, B.T., Lito, P., Namakydoust, A., Ng, K., Offin, M., Paik, P.K., Riely, G.J., Rudin, C.M., Yu, H.A., Zauderer, M.G., Donoghue, M.T.A., Łuksza, M., Greenbaum, B.D., Kris, M.G., Hellmann, M.D., 2020. COVID-19 in patients with Martín lung cancer. Annals of Oncology 31, 1386–1396. https://doi.org/10.1016/j.annonc.2020.06.007

44. Moro, F., Marquet, J., Piris, M., Michael, B.M., Sáez, A.J., Corona, M., Jiménez, C., Astibia, B., García, I., Rodríguez, E., García Hoz, C., Fortún Abete, J., Herrera, P., López Jiménez, J., 2020. Survival study of hospitalised patients with concurrent COVID-19 and haematological malignancies. British Journal of Haematology 190, e16–e20. https://doi.org/10.1111/bjh.16801

45. Mathew, D., Giles, J.R., Baxter, A.E., Oldridge, D.A., Greenplate, A.R., Wu, J.E., Alanio, C., Kuri-Cervantes, L., Pampena, M.B., D’Andrea, K., Manne, S., Chen, Z., Huang, Y.J., Reilly, J.P., Weisman, A.R., Ittner, C.A.G., Kuthuru, O., Dougherty, J., Nzingha, K., Han, N., Kim, J., Pattekar, A., Goodwin, E.C., Anderson, E.M., Weirick, M.E., Gouma, S., Arevalo, C.P., Bolton, M.J., Chen, F., Lacey, S.F., Ramage, H., Cherry, S., Hensley, S.E., Apostolidis, S.A., Huang, A.C., Vella, L.A., Unit†, T.Up.C.P., Betts, M.R., Meyer, N.J., Wherry, E.J., 2020. Deep immune profiling of COVID-19 patients reveals distinct immunotypes with therapeutic implications. Science 369. https://doi.org/10.1126/science.abc8511

46. Milano, F., Campbell, A.P., Guthrie, K.A., Kuypers, J., Englund, J.A., Corey, L., Boeckh, M., 2010. Human rhinovirus and coronavirus detection among allogeneic hematopoietic stem cell transplantation recipients. Blood 115, 2088–2094. https://doi.org/10.1182/blood-2009-09-244152

47. Moole, P.K.R., Papireddypalli, J.M.R., 2020. Effect of Deoxycholic Acid on Immune Cells - An Immunophenotyping Analysis of Peripheral Blood and Splenic Lymphocytes in CD57 Female Mice. International Journal of Pharmaceutical Investigation 10, 548– 552. https://doi.org/10.5530/ijpi.2020.4.95

48. NCT04407689, n.d. A Multicenter, Randomized, Double-blinded Placebo-controlled Study of Recombinant Interleukin-7 (CYT107) for Immune Restoration of Hospitalized Lymphopenic Patients With Coronavirus COVID-19 Infection in France and Belgium (Clinical trial registration No. NCT04407689). clinicaltrials.gov.

49. NCT04426201, n.d. A Multicenter, Randomized, Double-blinded Placebo-controlled Study of Recombinant Interleukin-7 (CYT107) for Immune Restoration of Hospitalized Lymphopenic Patients With Coronavirus COVID-19 Infection. US Oncology Cohort (Clinical trial registration No. NCT04426201). clinicaltrials.gov.

50. Païssé, S., Valle, C., Servant, F., Courtney, M., Burcelin, R., Amar, J., Lelouvier, B., 2016. Comprehensive description of blood microbiome from healthy donors assessed by 16S targeted metagenomic sequencing. Transfusion 56, 1138–1147. https://doi.org/10.1111/trf.13477

51. Park, M.D., 2020. Macrophages: a Trojan horse in COVID-19? Nature Reviews Immunology 20, 351–351. https://doi.org/10.1038/s41577-020-0317-2

52. Passamonti, F., Cattaneo, C., Arcaini, L., Bruna, R., Cavo, M., Merli, F., Angelucci, E., Krampera, M., Cairoli, R., Della Porta, M.G., Fracchiolla, N., Ladetto, M., Gambacorti Passerini, C., Salvini, M., Marchetti, M., Lemoli, R., Molteni, A., Busca, A., Cuneo, A., Romano, A., Giuliani, N., Galimberti, S., Corso, A., Morotti, A., Falini, B., Billio, A., Gherlinzoni, F., Visani, G., Tisi, M.C., Tafuri, A., Tosi, P., Lanza, F., Massaia, M., Turrini, M., Ferrara, F., Gurrieri, C., Vallisa, D., Martelli, M., Derenzini, E., Guarini, A., Conconi, A., Cuccaro, A., Cudillo, L., Russo, D., Ciambelli, F., Scattolin, A.M., Luppi, M., Selleri, C., Ortu La Barbera, E., Ferrandina, C., Di Renzo, N., Olivieri, A., Bocchia, M., Gentile, M., Marchesi, F., Musto, P., Federici, A.B., Candoni, A., Venditti, A., Fava, C., Pinto, A., Galieni, P., Rigacci, L., Armiento, D., Pane, F., Oberti, M., Zappasodi, P., Visco, C., Franchi, M., Grossi, P.A., Bertù, L., Corrao, G., Pagano, L., Corradini, P., 2020. Clinical characteristics and risk factors associated with COVID-19 severity in patients with haematological malignancies in Italy: a retrospective, multicentre, cohort study. The Lancet Haematology 7, e737–e745. https://doi.org/10.1016/S2352-3026(20)30251-9

53. Péron, J., Cropet, C., Tredan, O., Bachelot, T., Ray-Coquard, I., Clapisson, G., Chabaud, S., Philip, I., Borg, C., Cassier, P., Labidi Galy, I., Sebban, C., Perol, D., Biron, P., Caux, C., Menetrier-Caux, C., Blay, J.-Y., 2013. CD4 lymphopenia to identify end-of-life metastatic cancer patients. European Journal of Cancer 49, 1080–1089. https://doi.org/10.1016/j.ejca.2012.11.003

54. Pontelli, M.C., Castro, I.A., Martins, R.B., Veras, F.P., Serra, L.L., Nascimento, D.C., Cardoso, R.S., Rosales, R., Lima, T.M., Souza, J.P., Caetité, D.B., Lima, M.H.F. de Kawahisa, J.T., Giannini, M.C., Bonjorno, L.P., Lopes, M.I.F., Batah, S.S., Siyuan, L., Assad, R.L., Almeida, S.C.L., Oliveira, F.R., Benatti, M.N., Pontes, L.L.F., Santana, R.C., Vilar, F.C., Martins, M.A., Cunha, T.M., Calado, R.T., Alves-Filho, J.C., Zamboni, D.S., Fabro, A., Louzada-Junior, P., Oliveira, R.D.R., Cunha, F.Q., Arruda, E., 2020. Infection of human lymphomononuclear cells by SARS-CoV-2. bioRxiv 2020.07.28.225912. https://doi.org/10.1101/2020.07.28.225912

55. Puleston, D.J., Zhang, H., Powell, T.J., Lipina, E., Sims, S., Panse, I., Watson, A.S., Cerundolo, V., Townsend, A.R., Klenerman, P., Simon, A.K., 2014. Autophagy is a critical regulator of memory CD8(+) T cell formation. Elife 3. https://doi.org/10.7554/eLife.03706

56. Q, W., Q, C., H, Z., B, Y., X, H., Y, Z., X, Y., Mlk, C., C, X., 2020. Clinical outcomes of coronavirus disease 2019 (COVID-19) in cancer patients with prior exposure to immune checkpoint inhibitors. Cancer Commun (Lond) 40, 374–379. https://doi.org/10.1002/cac2.12077

57. Rd, S., Lj, O., Mj, S., 2011. Cancer immunoediting: integrating immunity’s roles in cancer suppression and promotion [WWW Document]. Science (New York, N.Y.). https://doi.org/10.1126/science.1203486

58. Robilotti, E.V., Babady, N.E., Mead, P.A., Rolling, T., Perez-Johnston, R., Bernardes, M., Bogler, Y., Caldararo, M., Figueroa, C.J., Glickman, M.S., Joanow, A., Kaltsas, A., Lee, Y.J., Lucca, A., Mariano, A., Morjaria, S., Nawar, T., Papanicolaou, G.A., Predmore, J., Redelman-Sidi, G., Schmidt, E., Seo, S.K., Sepkowitz, K., Shah, M.K., Wolchok, J.D., Hohl, T.M., Taur, Y., Kamboj, M., 2020. Determinants of COVID-19 disease severity in patients with cancer. Nat Med 26, 1218–1223. https://doi.org/10.1038/s41591-020-0979-0

59. Rugge, M., Zorzi, M., Guzzinati, S., 2020. SARS-CoV-2 infection in the Italian Veneto region: adverse outcomes in patients with cancer. Nature Cancer 1, 784–788. https://doi.org/10.1038/s43018-020-0104-9

60. Sanmamed, M.F., Perez-Gracia, J.L., Schalper, K.A., Fusco, J.P., Gonzalez, A., Rodriguez-Ruiz, M.E., Oñate, C., Perez, G., Alfaro, C., Martín-Algarra, S., Andueza, M.P., Gurpide, A., Morgado, M., Wang, J., Bacchiocchi, A., Halaban, R., Kluger, H., Chen, L., Sznol, M., Melero, I., 2017. Changes in serum interleukin-8 (IL-8) levels reflect and predict response to anti-PD-1 treatment in melanoma and non-small-cell lung cancer patients. Annals of Oncology 28, 1988–1995. https://doi.org/10.1093/annonc/mdx190

61. Schierwagen, R., Alvarez-Silva, C., Servant, F., Trebicka, J., Lelouvier, B., Arumugam, M., 2020. Trust is good, control is better: technical considerations in blood microbiome analysis. Gut 69, 1362–1363. https://doi.org/10.1136/gutjnl-2019-319123

62. Sheng, L., Jena, P.K., Hu, Y., Liu, H.-X., Nagar, N., Kalanetra, K.M., French, Samuel William, French, Samuel Wheeler, Mills, D.A., Wan, Y.-J.Y., 2017. Hepatic inflammation caused by dysregulated bile acid synthesis is reversible by butyrate supplementation. J Pathol 243, 431–441. https://doi.org/10.1002/path.4983

63. Shlomai, A., Ben-Zvi, H., Glusman Bendersky, A., Shafran, N., Goldberg, E., Sklan, E.H., 2020. Nasopharyngeal viral load predicts hypoxemia and disease outcome in admitted COVID-19 patients. Critical Care 24, 539. https://doi.org/10.1186/s13054-020-03244-3

64. Silvin, A., Chapuis, N., Dunsmore, G., Goubet, A.-G., Dubuisson, A., Derosa, L., Almire, C., Hénon, C., Kosmider, O., Droin, N., Rameau, P., Catelain, C., Alfaro, A., Dussiau, C., Friedrich, C., Sourdeau, E., Marin, N., Szwebel, T.-A., Cantin, D., Mouthon, L., Borderie, D., Deloger, M., Bredel, D., Mouraud, S., Drubay, D., Andrieu, M., Lhonneur, A.-S., Saada, V., Stoclin, A., Willekens, C., Pommeret, F., Griscelli, F., Ng, L.G., Zhang, Z., Bost, P., Amit, I., Barlesi, F., Marabelle, A., Pène, F., Gachot, B., André, F., Zitvogel, L., Ginhoux, F., Fontenay, M., Solary, E., 2020. Elevated Calprotectin and Abnormal Myeloid Cell Subsets Discriminate Severe from Mild COVID-19. Cell 182, 1401–1418.e18. https://doi.org/10.1016/j.cell.2020.08.002

65. Song, X., Sun, X., Oh, S.F., Wu, M., Zhang, Y., Zheng, W., Geva-Zatorsky, N., Jupp, R., Mathis, D., Benoist, C., Kasper, D.L., 2020. Microbial bile acid metabolites modulate gut ROR + regulatory T cell homeostasis. Nature 577, 410–415. https://doi.org/10.1038/s41586-019-1865-0

66. Sterlin, D., Mathian, A., Miyara, M., Mohr, A., Anna, F., Claer, L., Quentric, P., Fadlallah, J., Ghillani, P., Gunn, C., Hockett, R., Mudumba, S., Guihot, A., Luyt, C.-E., Mayaux, J., Beurton, A., Fourati, S., Lacorte, J.-M., Yssel, H., Parizot, C., Dorgham, K., Charneau, P., Amoura, Z., Gorochov, G., 2020. IgA dominates the early neutralizing antibody response to SARS-CoV-2. medRxiv 2020.06.10.20126532. https://doi.org/10.1101/2020.06.10.20126532

67. Takahashi, T., Ellingson, M.K., Wong, P., Israelow, B., Lucas, C., Klein, J., Silva, J., Mao, T., Oh, J.E., Tokuyama, M., Lu, P., Venkataraman, A., Park, A., Liu, F., Meir, A., Sun, J., Wang, E.Y., Casanovas-Massana, A., Wyllie, A.L., Vogels, C.B.F., Earnest, R., Lapidus, S., Ott, I.M., Moore, A.J., Shaw, A., Fournier, J.B., Odio, C.D., Farhadian, S., Cruz, C.D., Grubaugh, N.D., Schulz, W.L., Ring, A.M., Ko, A.I., Omer, S.B., Iwasaki, A., 2020. Sex differences in immune responses that underlie COVID-19 disease outcomes. Nature 1–6. https://doi.org/10.1038/s41586-020-2700-3

68. Westblade, L.F., Brar, G., Pinheiro, L.C., Paidoussis, D., Rajan, M., Martin, P., Goyal, P., Sepulveda, J.L., Zhang, L., George, G., Liu, D., Whittier, S., Plate, M., Small, C.B., Rand, J.H., Cushing, M.M., Walsh, T.J., Cooke, J., Safford, M.M., Loda, M., Satlin, M.J., 2020. SARS-CoV-2 Viral Load Predicts Mortality in Patients with and without Cancer Who Are Hospitalized with COVID-19. Cancer Cell. https://doi.org/10.1016/j.ccell.2020.09.007

69. Westmeier, J., Paniskaki, K., Karaköse, Z., Werner, T., Sutter, K., Dolff, S., Overbeck, M., Limmer, A., Liu, J., Zheng, X., Brenner, T., Berger, M.M., Witzke, O., Trilling, M., Lu, M., Yang, D., Babel, N., Westhoff, T., Dittmer, U., Zelinskyy, G., 2020. Impaired Cytotoxic CD8+ T Cell Response in Elderly COVID-19 Patients. mBio 11. https://doi.org/10.1128/mBio.02243-20

70. Wölfel, R., Corman, V.M., Guggemos, W., Seilmaier, M., Zange, S., Müller, M.A., Niemeyer, D., Jones, T.C., Vollmar, P., Rothe, C., Hoelscher, M., Bleicker, T., Brünink, S., Schneider, J., Ehmann, R., Zwirglmaier, K., Drosten, C., Wendtner, C., 2020. Virological assessment of hospitalized patients with COVID-2019. Nature 581, 465–469. https://doi.org/10.1038/s41586-020-2196-x

71. Xu, X., Chang, X.N., Pan, H.X., Su, H., Huang, B., Yang, M., Luo, D.J., Weng, M.X., Ma, L., Nie, X., 2020. [Pathological changes of the spleen in ten patients with coronavirus disease 2019(COVID-19) by postmortem needle autopsy]. Zhonghua Bing Li Xue Za Zhi 49, 576–582. https://doi.org/10.3760/cma.j.cn112151-20200401-00278

72. Yeoh, Y.K., Zuo, T., Lui, G.C.-Y., Zhang, F., Liu, Q., Li, A.Y., Chung, A.C., Cheung, C.P., Tso, E.Y., Fung, K.S., Chan, V., Ling, L., Joynt, G., Hui, D.S.-C., Chow, K.M., Ng, S.S.S., Li, T.C.-M., Ng, R.W., Yip, T.C., Wong, G.L.-H., Chan, F.K., Wong, C.K., Chan, P.K., Ng, S.C., 2021. Gut microbiota composition reflects disease severity and dysfunctional immune responses in patients with COVID-19. Gut. https://doi.org/10.1136/gutjnl-2020-323020

73. Zhang, H., Alsaleh, G., Feltham, J., Sun, Y., Napolitano, G., Riffelmacher, T., Charles, P., Frau, L., Hublitz, P., Yu, Z., Mohammed, S., Ballabio, A., Balabanov, S., Mellor, J., Simon, A.K., 2019. Polyamines Control eIF5A Hypusination, TFEB Translation, and Autophagy to Reverse B Cell Senescence. Mol Cell 76, 110–125.e9. https://doi.org/10.1016/j.molcel.2019.08.005

74. Ziegler, C.G.K., Allon, S.J., Nyquist, S.K., Mbano, I.M., Miao, V.N., Tzouanas, C.N., Cao, Y., Yousif, A.S., Bals, J., Hauser, B.M., Feldman, J., Muus, C., Wadsworth, M.H., Kazer, S.W., Hughes, T.K., Doran, B., Gatter, G.J., Vukovic, M., Taliaferro, F., Mead, B.E., Guo, Z., Wang, J.P., Gras, D., Plaisant, M., Ansari, M., Angelidis, I., Adler, H., Sucre, J.M.S., Taylor, C.J., Lin, B., Waghray, A., Mitsialis, V., Dwyer, D.F., Buchheit, K.M., Boyce, J.A., Barrett, N.A., Laidlaw, T.M., Carroll, S.L., Colonna, L., Tkachev, V., Peterson, C.W., Yu, A., Zheng, H.B., Gideon, H.P., Winchell, C.G., Lin, P.L., Bingle, C.D., Snapper, S.B., Kropski, J.A., Theis, F.J., Schiller, H.B., Zaragosi, L.-E., Barbry, P., Leslie, A., Kiem, H.-P., Flynn, J.L., Fortune, S.M., Berger, B., Finberg, R.W., Kean, L.S., Garber, M., Schmidt, A.G., Lingwood, D., Shalek, A.K., Ordovas-Montanes, J., HCA Lung Biological Network. Electronic address, HCA Lung Biological Network, 2020. SARS-CoV-2 Receptor ACE2 Is an Interferon-Stimulated Gene in Human Airway Epithelial Cells and Is Detected in Specific Cell Subsets across Tissues. Cell 181, 1016-1035.e19. https://doi.org/10.1016/j.cell.2020.04.035

