## Supplementary figures and images for "Prolonged SARS-CoV-2 RNA virus shedding and lymphopenia are hallmarks of COVID-19 in cancer patients with poor prognosis"

### Supplemental figures

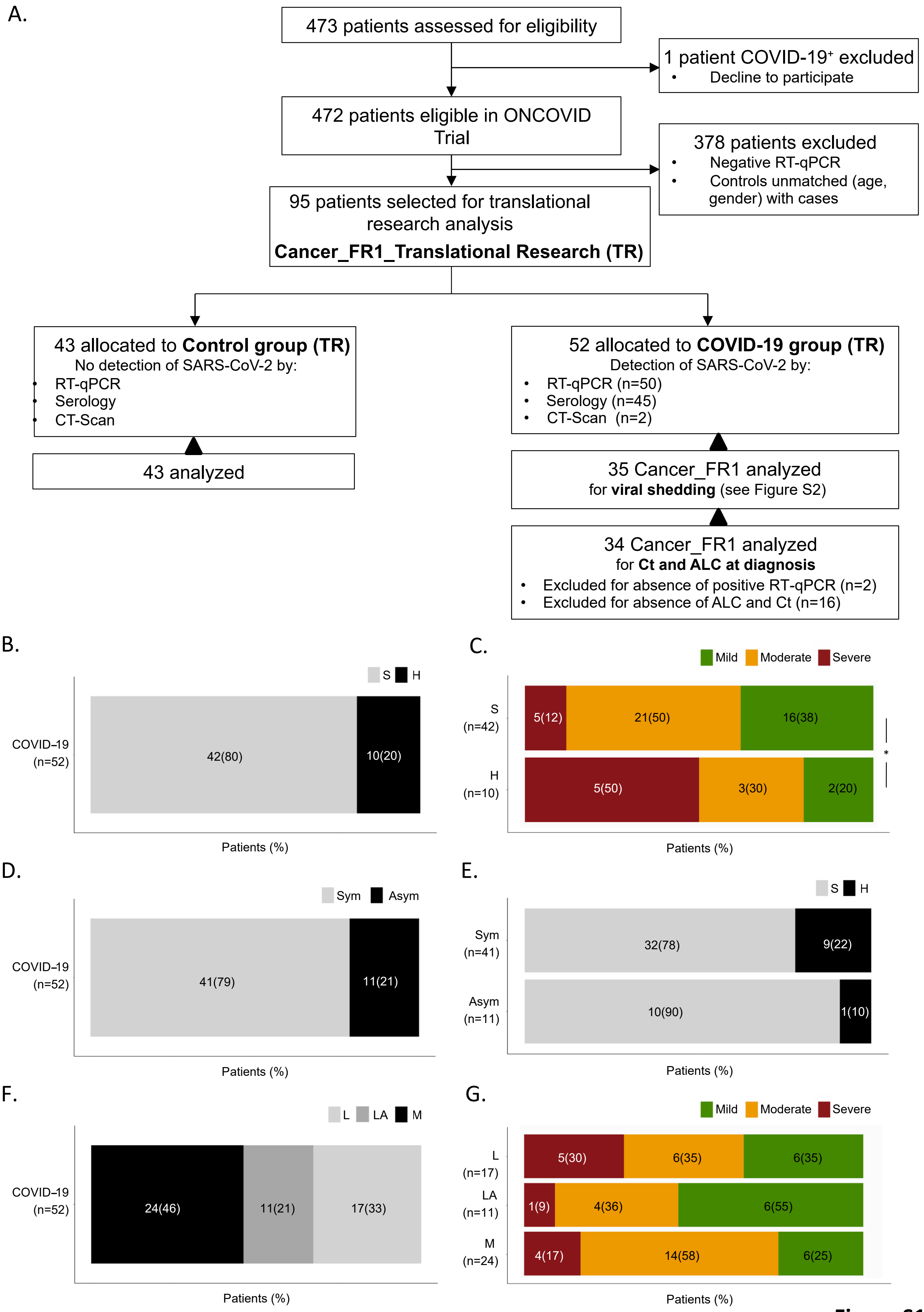

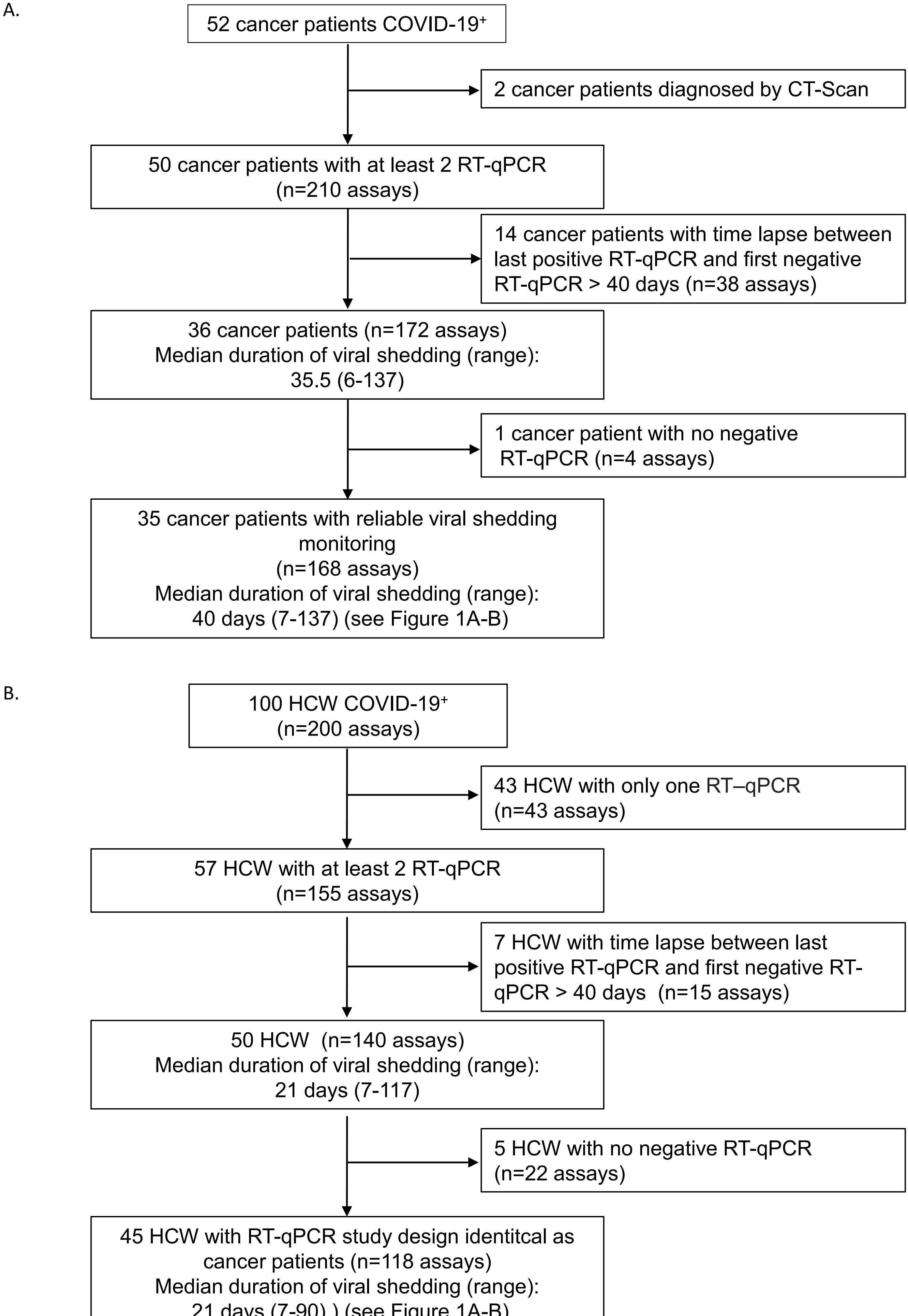

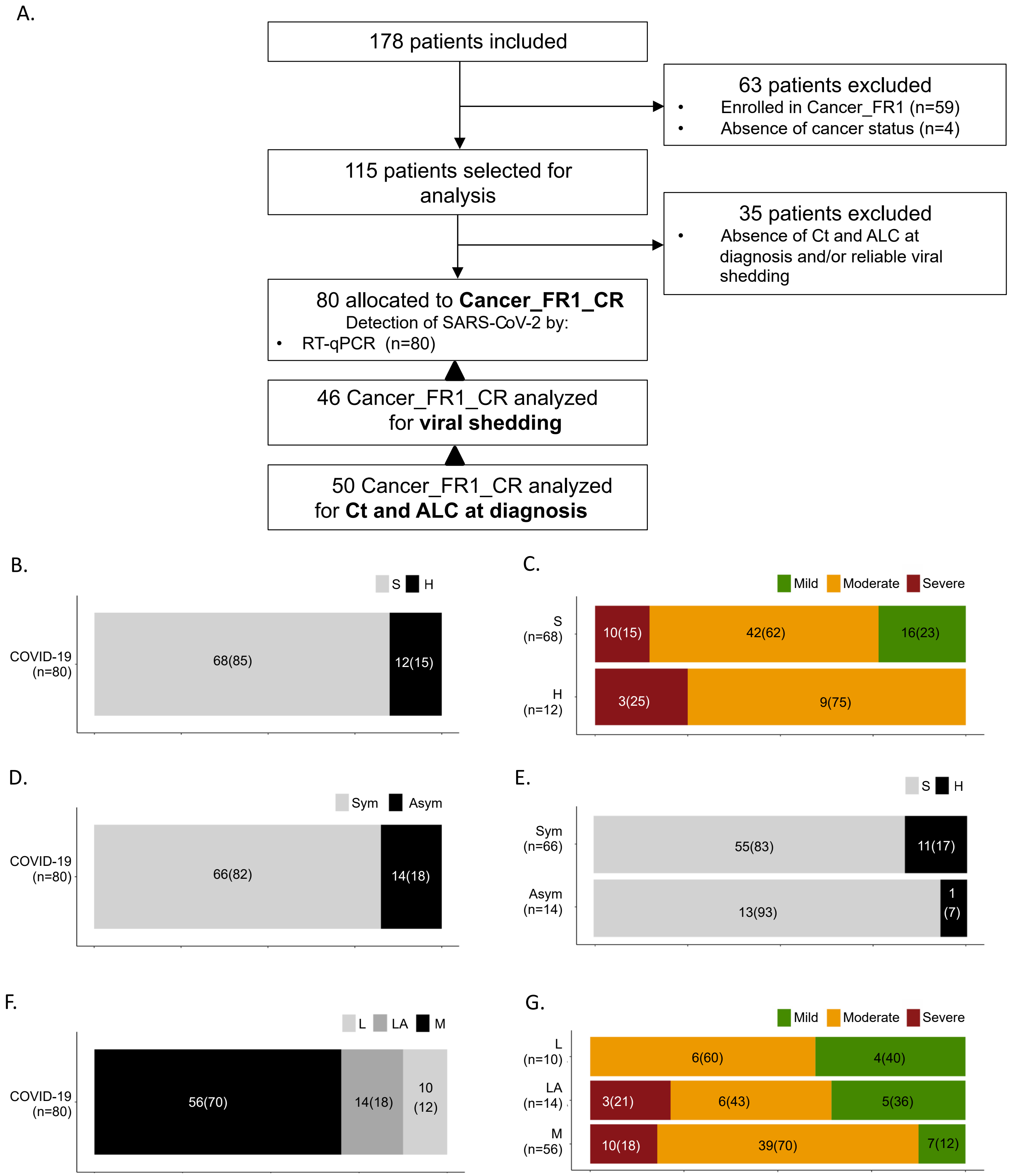

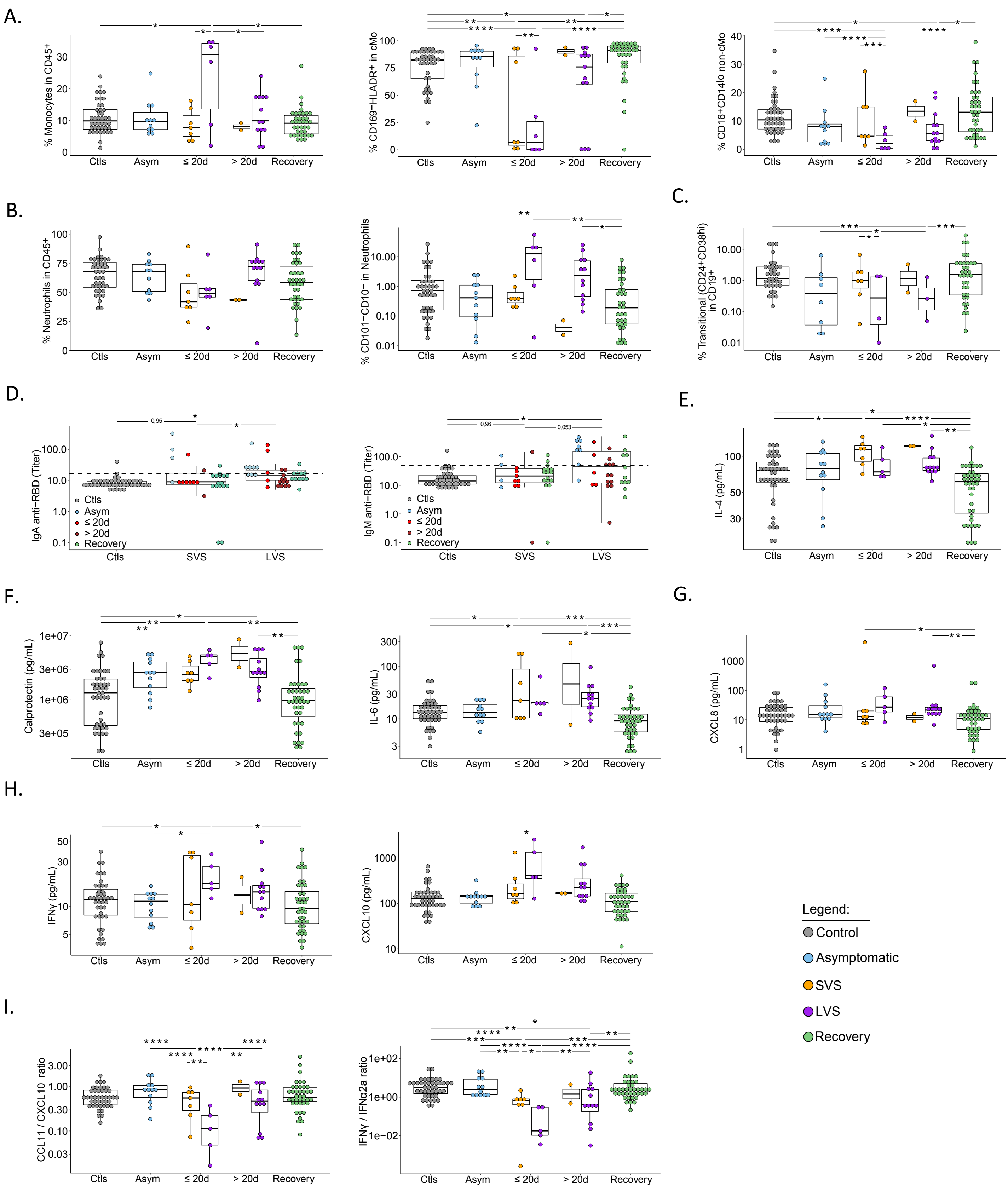

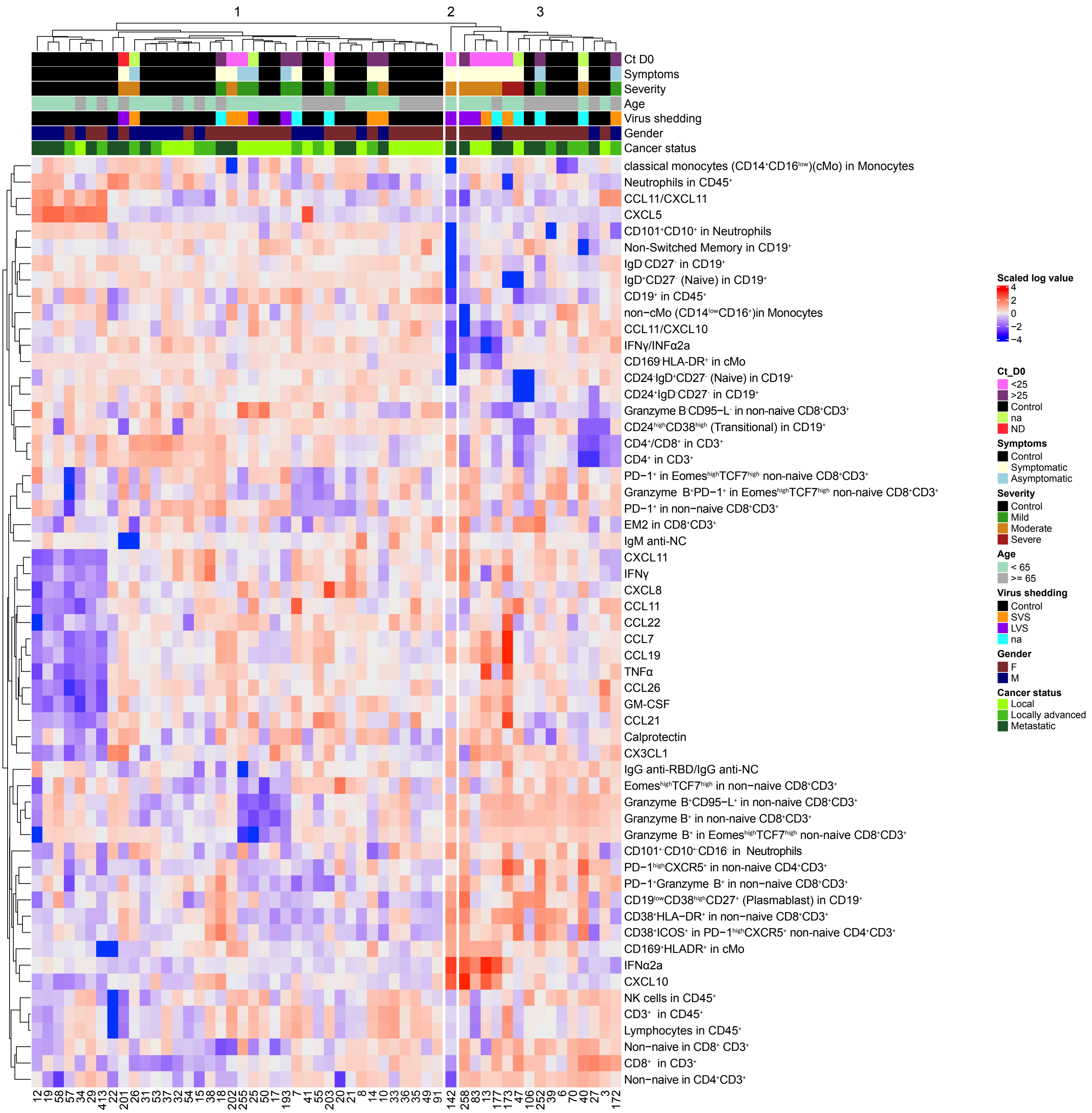

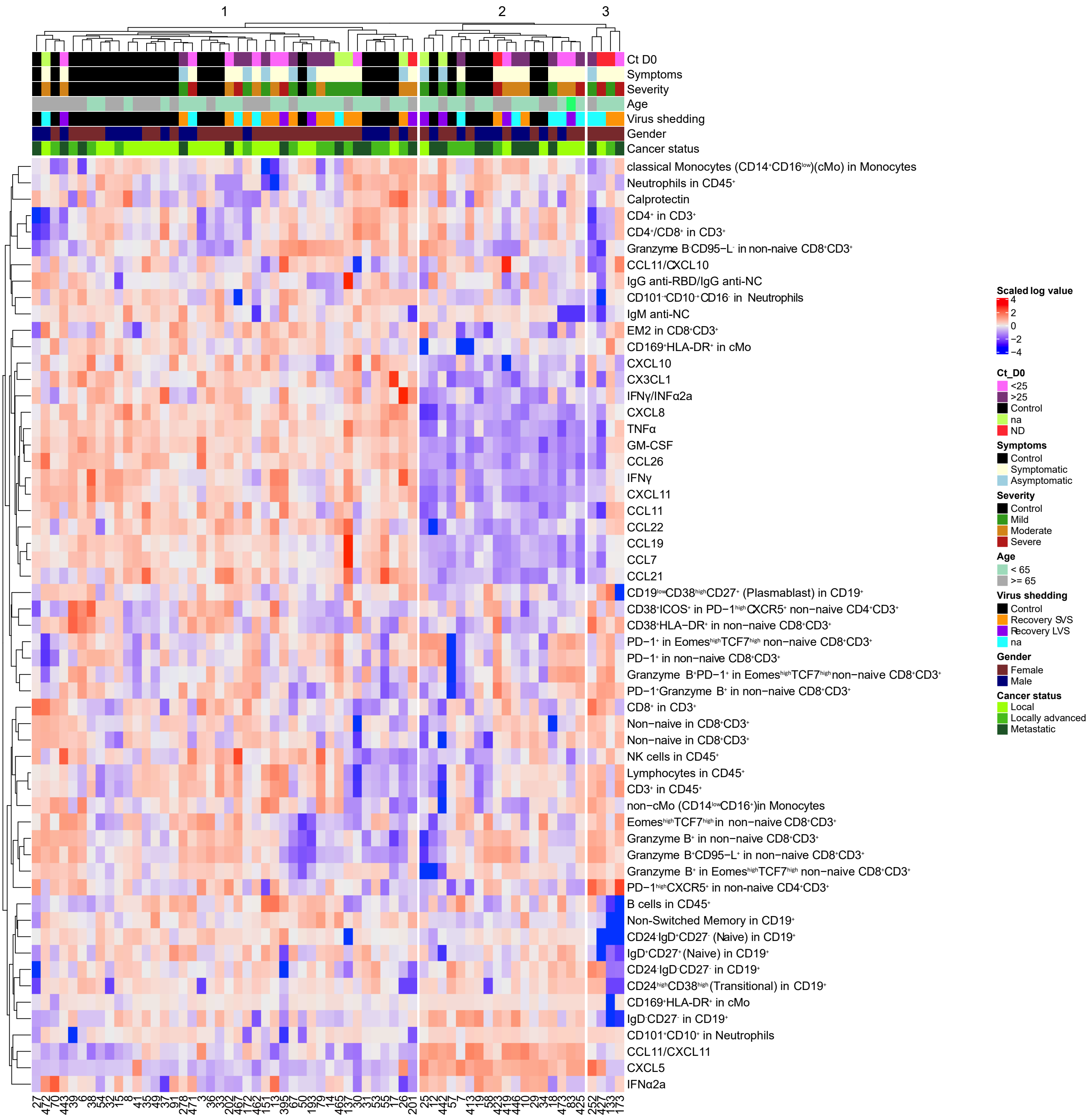

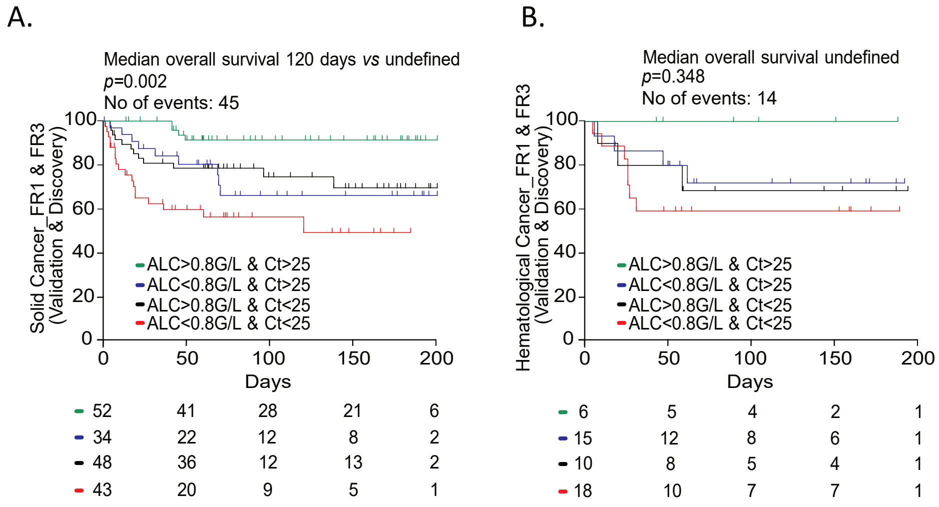

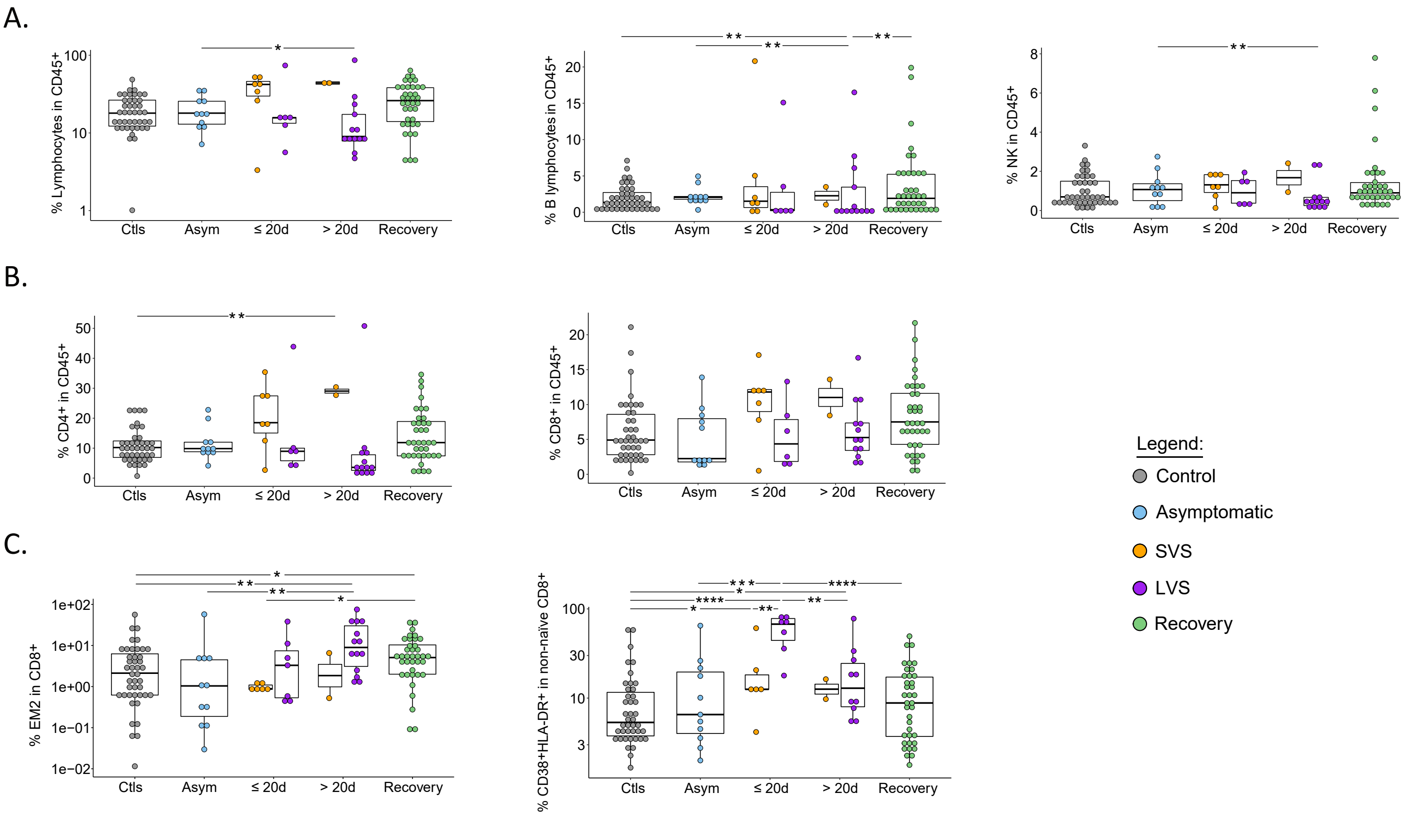

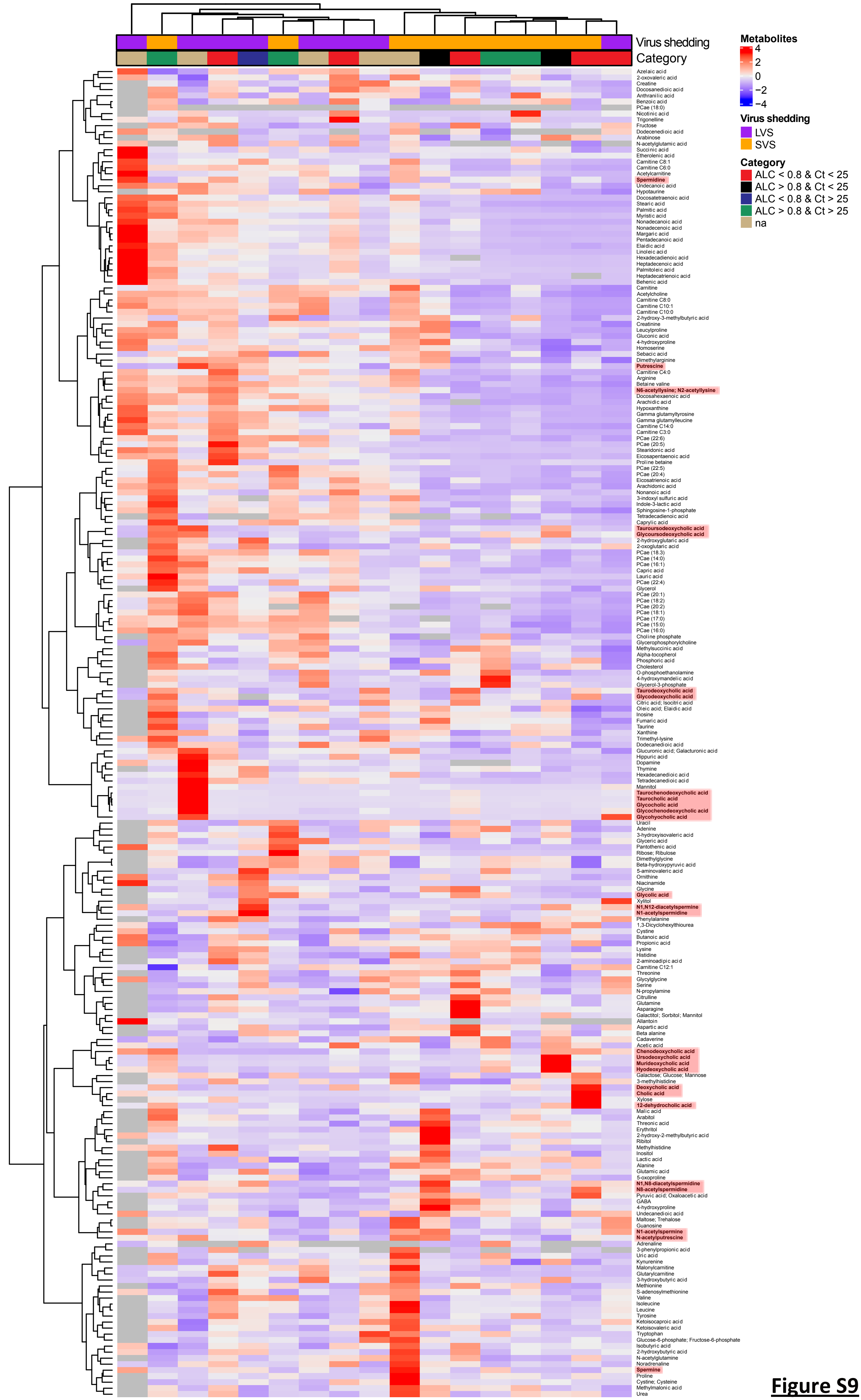

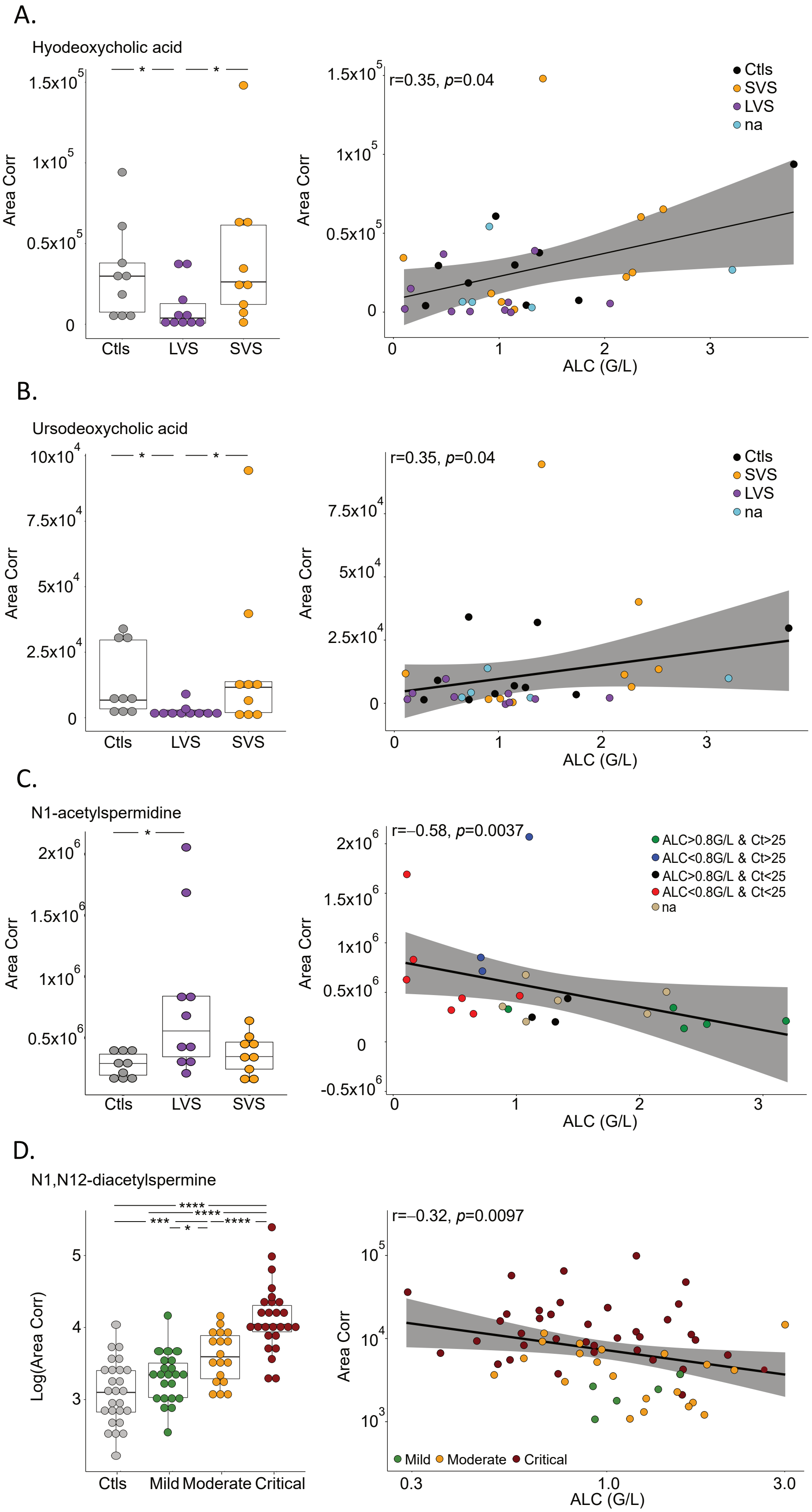
