## Supplemental tables for "Prolonged SARS-CoV-2 RNA virus shedding and lymphopenia are hallmarks of COVID-19 in cancer patients with poor prognosis"

**Table S1.** Clinical and viral characteristics of patients COVID-19$^+$ enrolling in ONCOVID. Clinical and viral characteristics of patients COVID-19$^+$ enrolling in three French COVID-19$^+$ and one Canadian cohort according to the duration of viral shedding (SVS *vs* LVS).

| | | | Cancer_FR1_TR | | | | Cancer_FR2 | | | Cancer_FR1_CR | | | Cancer_CA | | |
|---|---|---|---|---|---|---|---|---|---|---|---|---|---|---|---|
| | **Patient's characteristics** | | **COVID-19$^+$ (n=52)** | **SVS (n=17)** | **LVS (n=18)** | ***P*** | **SVS (n=90)** | **LVS (n=85)** | ***P*** | **SVS (n=37)** | **LVS (n=9)** | ***P*** | **SVS (n=49)** | **LVS (n=17)** | ***P*** |
| **Clinical characteristics** | **Age(year)** | **Median (range)** | 63 (31-89) | 56 (31-81) | 66 (39-89) | *0.08#* | 69 (23-94) | 72 (26-98) | *0.43#* | 61 (29-90) | 52 (21-67) | *0.10#* | 72 (20-94) | 68 (52-90) | *0.85#* |
| | **Gender –no(%)** | **Male** | 19 (37) | 6 (35) | 6 (33) | *0.90* | 42 (47) | 43 (51) | *0.6* | 20 (54) | 4 (44) | *0.60* | 28 (57) | 6 (35) | *0.12* |
| | | **Female** | 33 (63) | 11 (65) | 12 (67) | | 48 (53) | 42 (49) | | 17 (46) | 5 (56) | | 21 (43) | 11 (65) | |
| | **No of comorbidities –no(%)** | **0** | 21 (40) | 7 (36) | 6 (35) | *0.98* | 22 (24.5) | 21 (25) | *0.94* | 16 (43) | 7 (78) | *0.31* | - | - | |
| | | **1** | 15 (29) | 6 (33) | 6 (35) | | 22 (24.5) | 24 (28) | | 13 (35) | 1 (11) | | - | - | |
| | | **2** | 11 (21) | 3 (17) | 3 (18) | | 27 (30) | 24 (28) | | 7 (19) | 1 (11) | | - | - | |
| | | **3 or more** | 5 (10) | 2 (11) | 2 (12) | | 19 (21) | 16 (19) | | 1 (3) | 0 | | - | - | |
| | **Comorbidities –no(%)** | **COPD** | 3 (6) | 1 (8) | 2 (13) | | 14 (16) | 20 (24) | | 3 (8) | 0 | | 18 (38) | 5 (29) | |
| | | **BMI ≥ 30** | 11 (21) | 3 (23) | 3 (20) | | 13 (14) | 15 (18) | | 6 (16) | 0 | | 8 (16) | 2 (12) | |
| | | **Hypertension** | 31 (58) | 9 (69) | 7 (47) | *0.63* | 47 (52) | 48 (56) | *0.31* | 14 (38) | 1 (11) | *0.67* | - | - | 0.19 |
| | | **CHF** | 3 (6) | 0 | 1 (7) | | 36 (40) | 23 (27) | | 0 | 0 | | 19 (39) | 7 (41) | |
| | | **DM** | 5 (9) | 0 | 2 (13) | | 25 (28) | 18 (21) | | 8 (22) | 2 (22) | | 9 (18) | 9 (53) | |
| | **Type of cancer–no(%)** | **S** | 42 (80) | 16 (94) | 12 (67) | ***0.04*** | 71 (79) | 62 (73) | *0.40* | 30 (81) | 7 (78) | *0.83* | 39 (80) | 11 (65) | *0.22* |
| | | **H** | 10 (20) | 1 (6) | 6 (33) | | 12 (13) | 15 (18) | | 7 (19) | 2 (22) | | 10 (20) | 6 (35) | |
| | | **Unknown** | - | - | - | | 7 (8) | 8 (9) | | - | - | | - | - | |
| | **Cancer spread–no(%)** | **Localized** | 17 (33) | 4 (24) | 4 (22) | | - | - | | 5 (13) | 0 | | 15 (31) | 7 (41) | |
| | | **Locally advanced** | 11 (21) | 8 (47) | 1 (6) | ***0.01*** | - | - | | 4 (11) | 1 (11) | *0.81* | 3 (6) | 2 (12) | *0.80* |
| | | **Metastatic** | 24 (46) | 5 (29) | 13 (72) | | 11 (12) | 8 (9) | *0.55* | 28 (76) | 8 (88) | | 17 (35) | 6 (35) | |
| | | **Unknown** | - | - | - | | 79 (88) | 77 (91) | | - | - | | 14 (29) | 2 (12) | |
| | **Cancer status–no(%)** | **Remission or NED** | 22 (42) | 7 (41) | 5 (28) | *0.67* | - | - | | 6 (16) | 1 (11) | *0.92* | 17 (35) | 5 (29) | *0.76* |
| | | **SD/PR** | 14 (27) | 6 (35) | 7 (39) | | - | - | | 7 (19) | 2 (22) | | 6 (12) | 3 (18) | |
| | | **PD** | 16 (31) | 4 (24) | 6 (33) | | - | - | | 24 (65) | 6 (67) | | 6 (12) | 3 (18) | |
| | | **Unknown** | - | - | - | | - | - | | - | - | | 20 (41) | 6 (35) | |
| | **ECOG PS–no(%)** | **0** | 17 (33) | 8 (47) | 5 (28) | *0.17* | - | - | - | 16 (43) | 2 (22) | *0.45* | 11 (22) | 5 (29) | *0.13* |
| | | **1** | 23 (44) | 7 (41) | 6 (33) | | - | - | | 13 (35) | 5 (56) | | 19 (39) | 3 (18) | |
| | | **2 or more** | 12 (23) | 2 (12) | 7 (39) | | - | - | | 8 (22) | 2 (22) | | 13 (27) | 9 (53) | |
| | | **Unknown** | - | - | - | | - | - | | - | - | | 6 (12) | 0 | |
| | **Anticancer therapy–no(%)** | **None*** | 32 (61) | 14 (82) | 12 (67) | *0.29* | - | - | - | 14 (38) | 3 (33) | *0.80* | 26 (53) | 8 (47) | *0.67* |
| | | **Chemotherapy** | 8 (15) | 2 (12) | 4 (22) | | - | - | | 14 (38) | 4 (44) | | 12 (24) | 4 (24) | |
| | | **Radiotherapy** | 1 (2) | 0 | 0 | | - | - | | 1 (3) | 1 (11) | | 0 | 0 | |
| | | **Surgery** | 2 (4) | 1 (6) | 1 (6) | | - | - | | 2 (6) | 0 | | 1 (2) | 0 | |
| | | **HT** | 3 (6) | 0 | 2 (11) | | - | - | | 0 | 0 | | 2 (4) | 1 (6) | |
| | | **Immunotherapy** | 0 () | 0 | 0 | | - | - | | 2 (6) | 2 (22) | | 2 (4) | 3 (18) | |
| | | **Others** | 4 (8) | 1 56) | 2 (11) | | - | - | | 4 (12) | 1 (11) | | 6 (12) | 1 (6) | |
| | **Clinical course–no(%)** | **Day hospital** | 14 (27) | 10 (59) | 2 (11) | | - | - | | 14 (38) | 3 (33) | | 4 (8) | 2 (12) | |
| | | **Hospitalization** | 32 (61) | 4 (23) | 15 (83) | **<0.01** | - | - | **0.02** | 23 (63) | 5 (56) | *0.29* | 44 (90) | 15 (88) | *0.77* |
| | | **ICU** | 6 (11) | 3 (18) | 1 (6) | | 9 (10) | 19 (22) | | 0 | 1 (11) | | 1 (2) | 0 | |
| | **Death-–no(%)** | **Yes** | 4 (7) | 0 | 3 (18) | *0.22* | 5 (6) | 4 (5) | *0.80* | 7 (19) | 3 (33) | *0.38* | 3 (6) | 1 (6) | *0.97* |
| **Viral characteristics** | **Evaluable Ct–no(%)** | **Day 0** | 37 (71) | 13 (76) | 12 (67) | | 68 (76) | 66 (78) | | 15 (41) | 5 (56) | | - | - | - |
| | **Median Ct (range)** | | 25 (11-36) | 27 (20-36) | 23 (19-30) | ***0.06#*** | 29 (16-34) | 24 (12-34) | **<0.01** | 29 (19-33) | 19 (13-31) | ***0.03#*** | | | |
| | **Treatment –no(%)** | **HCQ** | - | - | - | | 54 (60) | 63 (74) | | - | - | | - | - | - |
| | | **AZI** | - | - | - | | 85 (94) | 82 (96) | *0.61* | - | - | | - | - | - |
| | | **HCQ+AZI** | 19 (37) | 3 (18) | 7 (39) | *0.16* | 52 (58) | 62 (73) | | 5 (13) | 1 (11) | *0.84* | - | - | - |

AZI: Azithromycin; BMI: Body Mass Index; CHF: Congestive heart failure; COPD: Chronic Obstructive Pulmonary Disease; Ct: Cycle threshold; CR: Clinical Research; H: Hematological malignancies; DM: Diabetes mellitus; HCQ: Hydroxychloroquine; HT: Hormonal therapy; ICU: Intensive Care Unit; LVS: Long Viral Shedding; na: not applicable; NED: No Evidence of Disease; no: number; PD: Progressive Disease; PS: Performance Status; S: Solid tumors; SD/PR: Stable Disease/Partial Response; SVS: Short Viral Shedding; TR: Translational Research; * in the 4 weeks before inclusion.

Statistical analyses: Mann-Whitney#, Chi-Square or Fisher's exact tests.

**Table S2.** Clinical characteristics of Cancer_FR1 available for translational research

| Patient's characteristics | | | Controls (n=43) | COVID-19$^{+}$ | | | | Recovery (n=40) |
|---|---|---|---|---|---|---|---|---|
| | | | | SVS+LVS (n=25) | SVS (n=10) | LVS (n=15) | *P* | |
| Clinical characteristics | **Age(year)** | **Median (range)** | 64 (33-82) | 59 (31-89) | 52 (31-62) | 64 (39-89) | *0.03* | 62,5 (31-89) |
| | **Gender –no(%)** | **Male** | 18 (42) | 9 (36) | 4 (40) | 5 (33) | *0.73* | 15 (38) |
| | | **Female** | 25 (58) | 16 (64) | 6 (60) | 10 (67) | | 25 (62) |
| | **N of comorbidities–no(%)** | **0** | 13 (30) | 10 (40) | 5 (50) | 5 (33) | *0.43* | 16 (40) |
| | | **1** | 17 (39) | 6 (24) | 1 (10) | 5 (33) | | 13 (33) |
| | | **2** | 7 (16) | 6 (24) | 2 (20) | 4 (27) | | 7 (17) |
| | | **3 or more** | 6 (14) | 3 (12) | 2 (20) | 1 (7) | | 4 (10) |
| | **Comorbidities–no(%)** | **COPD** | 4 (9) | 2 (8) | 1 (10) | 1 (7) | *0.90* | 3 (8) |
| | | **BMI ≥ 30** | 7 (16) | 6 (24) | 3 (30) | 3 (20) | | 10 (25) |
| | | **Hypertension** | 20 (46) | 10 (40) | 4 (40) | 6 (40) | | 17 (42) |
| | | **CHF** | 2 (5) | 1 (4) | 0 (0) | 1 (7) | | 1 (2) |
| | | **DM** | 5 (12) | 3 (12) | 1 (10) | 2 (13) | | 3 (8) |
| | **Type of cancer–no(%)** | **S** | 40 (93) | 18 (72) | 9 (90) | 9 (60) | *0.10* | 34 (85) |
| | | **H** | 3 (7) | 7 (28) | 1 (10) | 6 (40) | | 6 (15) |
| | **Cancer spread–no(%)** | **Localized** | 15 (35) | 6 (24) | 3 (30) | 3 (20) | *0.18* | 14 (35) |
| | | **Locally advanced** | 12 (28) | 4 (16) | 3 (30) | 1 (7) | | 9 (23) |
| | | **Metastatic** | 16 (37) | 15 (60) | 4 (40) | 11 (73) | | 17 (42) |
| | **Cancer status–no(%)** | **Remission or NED** | 10 (23) | 10 (40) | 5 (50) | 5 (33.3) | *0.66* | 21 (53) |
| | | **SD/PR** | 9 (21) | 8 (32) | 3 (30) | 5 (33.3) | | 6 (15) |
| | | **PD** | 24 (56) | 7 (28) | 2 (20) | 5 (33.3) | | 13 (33) |
| | **ECOG PS–no(%)** | **0** | 26 (60) | 8 (32) | 3 (30) | 5 (33.3) | *0.66* | 16 (40) |
| | | **1** | 14 (33) | 10 (40) | 5 (50) | 5 (33.3) | | 13 (33) |
| | | **2 or more** | 3 (7) | 7 (28) | 2 (20) | 5 (33.3) | | 10 (25) |
| | **Anticancer therapy–no(%)** | **None*** | 28 (65) | 18 (72) | 9 (90) | 9 (60) | *0.10* | 34 (85) |
| | | **Chemotherapy** | 8 (19) | 5 (20) | 1 (10) | 4 (27) | | 4 (10) |
| | | **Radiotherapy** | 6 (14) | 0 (0) | 0 (0) | 0 (0) | | 0 (0) |
| | | **Surgery** | 0 (0) | 1 (4) | 0 (0) | 1 (6) | | 0 (0) |
| | | **Hormonal therapy** | 1 (2) | 2 (8) | 0 (0) | 2 (13) | | 1 (2) |
| | | **Immunotherapy** | 3 (7) | 0 (0) | 0 (0) | 0 (0) | | 0 (0) |
| | | **Others** | 1 (2) | 3 (12) | 1 (10) | 2 (13) | | 4 (10) |
| | **Clinical course–no(%)** | **Day hospital** | 33 (77) | 7 (28) | 5 (50) | 2 (13) | ***0.04*** | 13 (33) |
| | | **Hospitalization** | 10 (23) | 15 (60) | 3 (30) | 12 (80) | | 22 (55) |
| | | **ICU** | - | 3 (12) | 2 (20) | 1 (7) | | 5 (12) |
| | **Death-–no(%)** | **Yes** | 5 (12) | 2 (8) | 0 (0) | 2 (13) | *0.5* | 1 (2) |

BMI: Body Mass Index; CHF: Congestive heart failure; COPD: Chronic Obstructive Pulmonary Disease; H: Hematological malignancies; DM: Diabetes mellitus; ICU: Intensive Care Unit; LVS: Long Viral Shedding; NED: No Evidence of Disease; no: number; PD: Progressive Disease; PS: Performance Status; S: Solid tumors; SD/PR: Stable Disease/Partial Response; SVS: Short Viral Shedding; *in the 4 weeks before inclusion. Statistical analyses: Mann-Whitney#, Chi-Square or Fisher's exact tests.
